# Genome-wide association study of 1,391 plasma metabolites in 6,136 Finnish men identifies 303 novel signals and provides biological insights into human diseases

**DOI:** 10.1101/2021.10.19.21265094

**Authors:** Xianyong Yin, Lap Sum Chan, Debraj Bose, Anne U. Jackson, Peter VandeHaar, Adam E. Locke, Christian Fuchsberger, Heather M. Stringham, Ketian Yu, Lilian Fernandes Silva, Susan K. Service, Daiwei Zhang, Emily C. Hector, Erica Young, Liron Ganel, Indraniel Das, Haley Abel, Michael R. Erdos, Lori L. Bonnycastle, Johanna Kuusisto, Nathan O. Stitziel, Ira Hall, Gregory R. Wagner, FinnGen, Jian Kang, Jean Morrison, Charles F. Burant, Francis S. Collins, Samuli Ripatti, Aarno Palotie, Nelson B. Freimer, Karen L. Mohlke, Laura J. Scott, Xiaoquan Wen, Eric B. Fauman, Markku Laakso, Michael Boehnke

## Abstract

Few studies have explored the impact of rare variants (minor allele frequency, MAF<1%) on highly heritable plasma metabolites identified in metabolomic screens. The Finnish population provides an ideal opportunity for such explorations, given the multiple bottlenecks and expansions that have shaped its history, and the enrichment for many otherwise rare alleles that has resulted. Here, we report genetic associations for 1,391 plasma metabolites in 6,136 men from the late-settlement region of Finland. We identify 303 novel association signals, more than one third at variants rare or enriched in Finns. Many of these signals identify genes not previously implicated in metabolite genome-wide association studies and suggest mechanisms for diseases and disease-related traits.

## Main text

The Finns are a geographically and linguistically isolated population who have experienced multiple population bottlenecks and expansions. This population history has resulted in large allele-frequency differences between Finns and non-Finnish Europeans (NFE), which are most pronounced in northern and eastern Finland, regions first settled in the 15th-16th centuries (“late settlement Finland”)^1^. In a previous study of 64 cardiometabolic traits in ∼20,000 individuals from these regions, we took advantage of the enrichment of otherwise rare alleles to identify 26 novel trait-associated rare deleterious alleles, 19 of which were >20-fold more frequent in late settlement Finns than in NFE^2^. These results suggested this allele-frequency enrichment could be leveraged to identify novel rare-variant associations for additional quantitative traits. Here, we do so, reporting genome-wide association study (GWAS) results for 1,391 plasma metabolites (Metabolon platform) in 6,136 participants in METSIM, a study of middle-aged and older men recruited from a single site in late-settlement northeast Finland^3^, who were part of our previous study^2^.

Metabolites are small molecules that play a pivotal role in cellular and physiological processes and their observed levels in biofluids can reflect those processes^4^. Most metabolomics studies are performed in blood (plasma or serum) which reflects the aggregate production and consumption of metabolites by tissues^4^. Abnormal metabolite levels are commonly associated with human diseases and disease-related traits, making them useful aids to understand disease mechanisms and to identify biomarkers for disease diagnosis, prognosis, and treatment monitoring^4^. Many metabolites are highly heritable, and previous metabolite GWAS have identified common variants^5-15^; the impact of rare variants on metabolites is less well studied^16,17^.

We identify 2,030 independent association signals (metabolite-index variant pairs) for 803 metabolites and demonstrate 946 genetic colocalizations of 248 metabolites with 105 diseases and disease-related traits. Many of these associations identify genes not previously implicated in metabolite GWAS and suggest mechanisms for these diseases and traits. Of the 2,030 association signals, 303 are novel; of these 303 signals, 111 are at 70 variants rare or >10-fold more frequent (“enriched”) in Finns compared to NFE, 78 are for 44 metabolites identified since 2015 on the Metabolon platform, and 17 are at variants on the X chromosome, which has often been ignored in previous metabolite GWAS. This study highlights the advantages of the Finnish population for rare-variant genetic association studies and the utility of integrating metabolite and disease genetic associations in disentangling disease mechanisms.

## Results

### GWAS on 1,391 metabolites

We assayed 1,544 plasma metabolites using the Metabolon DiscoveryHD4 mass spectrometry platform (Supplementary Tables 1-2) in 6,136 randomly-selected METSIM participants who were non-diabetic at baseline and passed quality control (QC) (Supplementary Table 3; Supplementary Fig. 1). 1,391 metabolites were successfully quantified in ≥500 of these 6,136 participants. We created a METSIM imputation reference panel of >26M genetic variants by integrating genome and exome sequence and array genotypes in 2,922 METSIM participants (Methods; Supplementary Table 4). We used this reference panel to impute genotypes in all METSIM participants.

We carried out GWAS across >16M variants with imputation r^2^≥0.3 and minor allele count (MAC)≥5 in the 6,136 METSIM participants for the 1,391 (correlated) metabolites (Methods; Supplementary Table 5; Supplementary Fig. 1). Single-variant association tests identified 305,555 associations at 109,368 variants for 803 metabolites at *P*<7.2×10^−11^=5.0×10^−8^/692 (Bonferroni correction for 692 principal components that together explained 95%^15^ of phenotypic variance for the 1,391 correlated metabolites; Methods). The GWAS *p-*values for each metabolite were well calibrated (genomic control inflation factor median=1.00, range=0.92-1.07; Supplementary Fig. 2). We built a multi-phenotype GWAS browser (PheWeb) (https://pheweb.org/metsim-metab/) to visualize and make publicly available our results for all 1,391 GWAS (Fig. 1; see Discussion).

**Figure 1:**
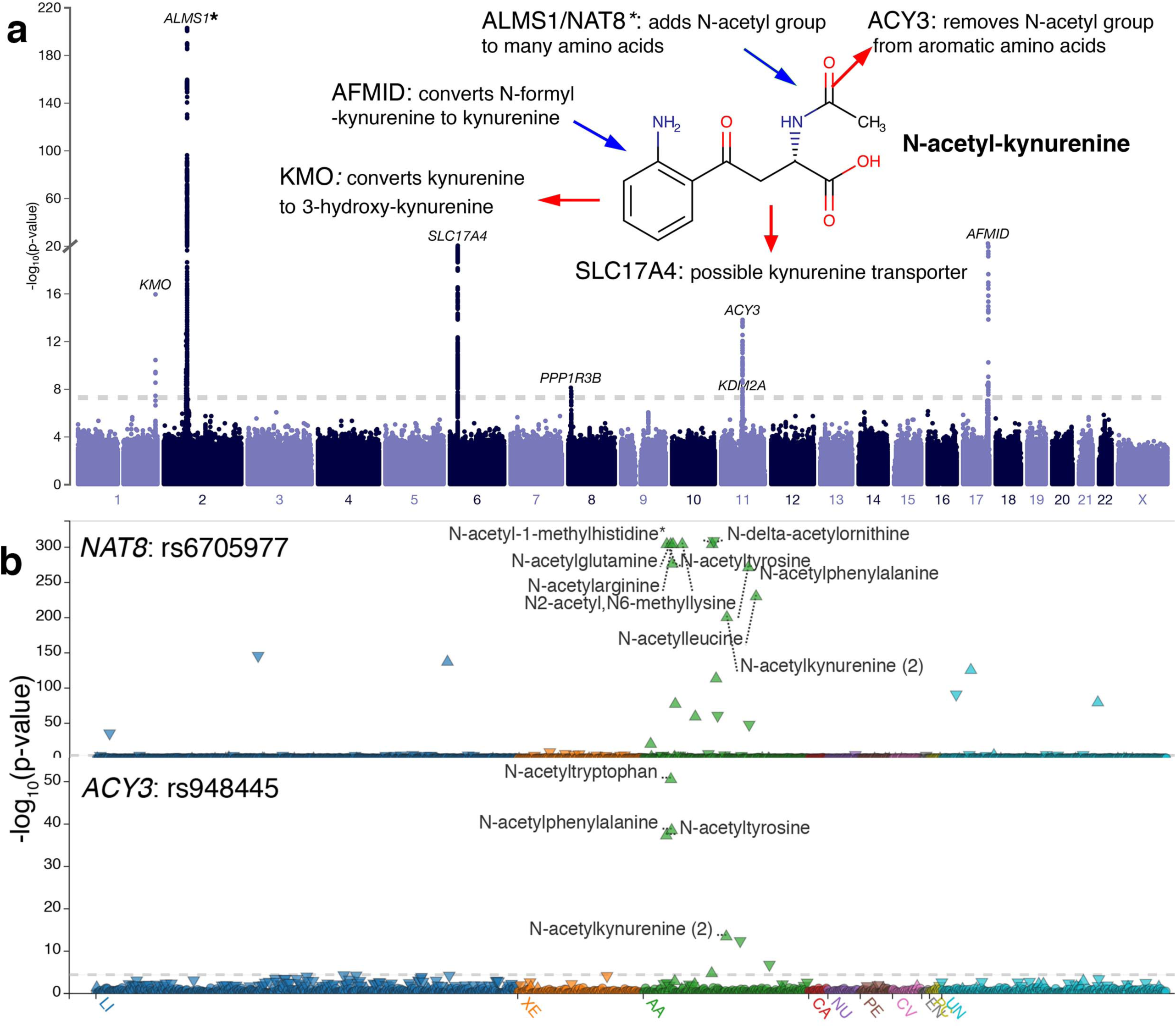
Our METSIM Metabolomics PheWeb facilitates the characterization of genetic associations and gene activities. a) Manhattan plot for N-acetylkynurenine highlighting the roles of the associated genes (https://pheweb.org/metsim-metab/pheno/C100006378). Chemical structure for N-acetylkynurenine and activities for the associated genes are added manually on top of the Manhattan plot. b) Stacked PheWeb plots show significant associations between rs6705977 (*NAT8*, https://pheweb.org/metsim-metab/variant/2:73622043-C-G) and fifteen N-acetylated molecules, and the more restricted set of associations between rs948445 (*ACY3*, https://pheweb.org/metsim-metab/variant/11:67647021-C-T) and four N-acetylated aromatic amino acids. LI: lipid; XE: xenobiotics; AA: amino acid; CA: carbohydrate; NU: nucleotide; PE: peptide; CV: cofactor and vitamin; EN: energy; PC: partially characterized; UN: unnamed.

Since body mass index (BMI) influences levels of many metabolites^18^, we repeated all 1,391 GWAS with BMI as an additional covariate. Results with and without BMI adjustment were generally very similar, with Pearson correlation coefficient r=0.999 for effect size estimates and -log_10_*p*-values for variant-metabolite pairs with *P*<7.2×10^−11^ in either of the two analyses (Supplementary Fig. 3). Supplementary Table 6 lists the 83 associations with substantially different effect sizes (ratio≥1.20) with and without BMI adjustment. In what follows, we present results for analyses without BMI adjustment.

### Detecting independent association signals

To identify (nearly) independent association signals, we carried out chromosome-wide stepwise conditional analysis for each chromosome-metabolite pair with ≥1 association at *P*<5.0×10^−8^. Conditional analysis identified 2,030 association signals at 1,143 index variants for 803 metabolites at *P*<7.2×10^−11^ (Table 1; Supplementary Table 7; Supplementary Figs. 4-5). The 1,143 index variants were of high imputation quality (r^2^ median=0.99, range=0.63-1.00). 311 (27.2%) of the 1,143 index variants were associated (*P*<7.2×10^−11^) with ≥2 metabolites, suggesting widespread pleiotropy (Supplementary Fig. 6). Among the 1,143 index variants, 121 (for 125 metabolites) are rare in METSIM and 99 (for 148 metabolites) have MAF>10-fold greater in METSIM than in NFE (gnomAD v3.1); 58 of these variants are both rare and enriched in Finns (Fig. 2a; Supplementary Table 7).

**Table 1:** Summary of the 2,030 genetic association signals by metabolite biochemical class

| **Biochemical class**<br>**and abbreviation** | **Metabolites** |  | **Association signals** |  |
| --- | --- | --- | --- | --- |
|  | **Total** | **Significant** | **Total** | **Novel** |
| Lipid (LI) | 548 | 357 | 903 | 74 |
| Amino acid (AA) | 215 | 154 | 441 | 73 |
| Xenobiotics (XE) | 163 | 52 | 91 | 16 |
| Nucleotide (NU) | 42 | 26 | 65 | 29 |
| Peptide (PE) | 42 | 18 | 28 | 7 |
| Cofactors and vitamins (CV) | 38 | 25 | 69 | 27 |
| Carbohydrate (CA) | 25 | 20 | 38 | 7 |
| Energy (EN) | 10 | 4 | 11 | 3 |
| Partially characterized (PC) | 16 | 8 | 20 | 1 |
| Unnamed (UN) | 292 | 139 | 364 | 66 |
| **Total** | **1,391** | **803** | **2,030** | **303** |

Significant: number of metabolites with at least one association signal with $P < 7.2 \times 10^{-11}$ .

**Figure 2:**
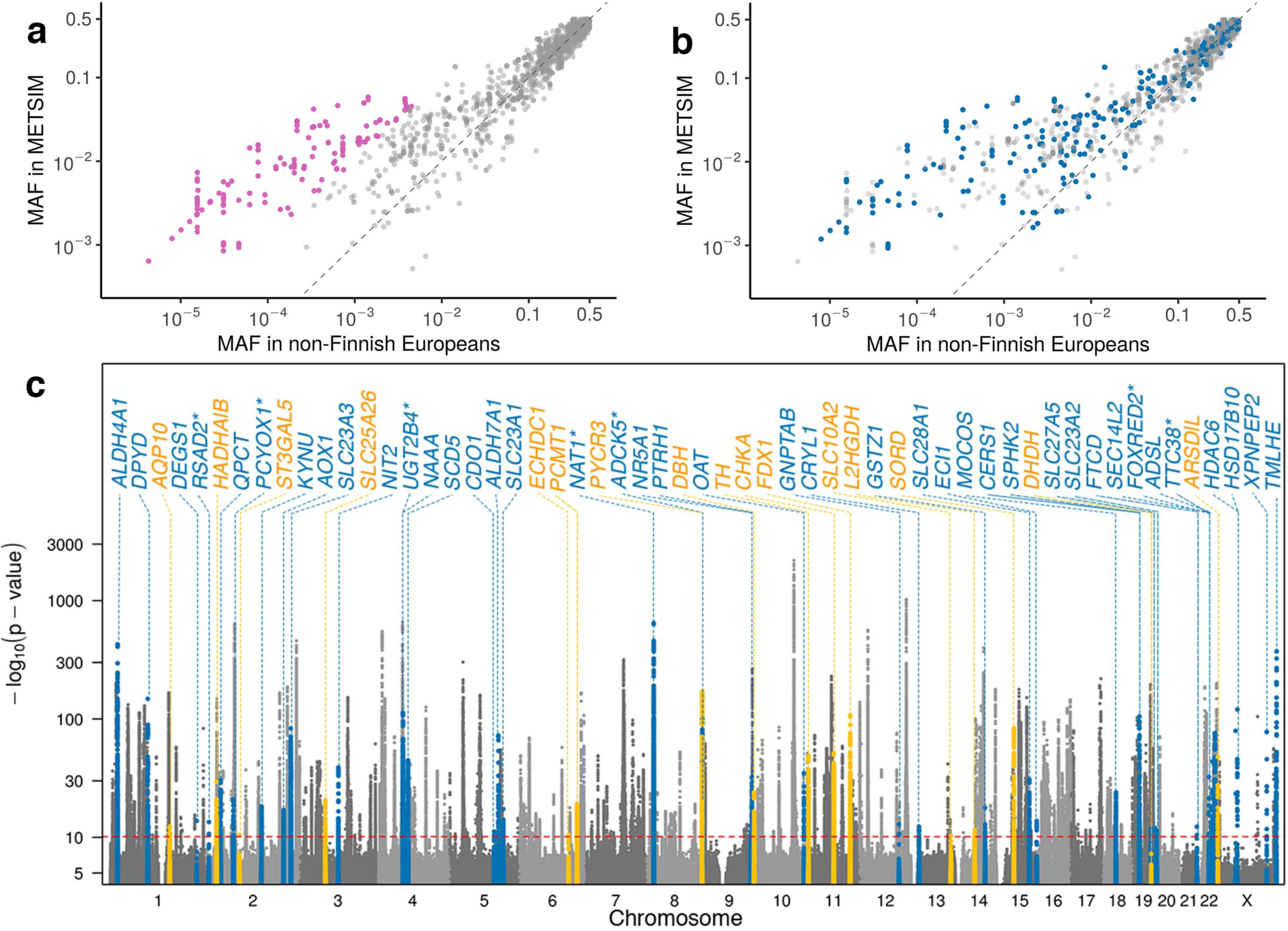
Characterization of the 2,030 significant metabolite genetic association signals in the 6,136 METSIM participants with Metabolon metabolomics data. Comparison of MAFs for the 1,143 index variants between METSIM and non-Finnish Europeans in gnomAD v3.1; index variants are colored a) purple if MAF>10-fold greater in METSIM than in non-Finnish Europeans; or b) blue they represent novel association signals. The dashed line is of slope one through the origin. c) Overlaid Manhattan plots of the 1,391 metabolite GWAS. The red dashed line depicts *P*=7.2×10^−11^. The associations at 40 novel putative causal genes within novel regions (blue) and 18 novel putative causal genes within previously reported regions (maize) are highlighted. The seven novel putative causal genes implicated only by fine-mapping analysis are starred. *HADHA/B* represents the *HADHA* and *HADHB* genes and *ARSD/L* the *ARSD* and *ARSL* genes.

Index variants explained from 0.7% to 62.0% (median=1.4%; Supplementary Fig. 7) of the phenotypic variance of the corresponding metabolite; 99 index variants explained ≥10% of the variance (Supplementary Table 8), including three missense variants with >10-fold greater frequency in METSIM than in NFE. For example, the putatively-deleterious *AFMID* missense variant p.Ala41Pro (rs77585764; MAF=5.4% in METSIM vs. 0.38% in NFE) explained 15.5% of the variance in N-formylanthranilic acid. *AFMID* encodes arylformamidase, an enzyme which catalyzes N-formylanthranilate to produce anthranilate and formate^19^.

### Fine mapping

To fine map the causal variants for the 2,030 association signals, we created 2 Mb regions centered on each index variant and merged overlapping regions associated with the same metabolite, resulting in 1,501 regions. We used Bayesian fine-mapping^20^ with a uniform prior to calculate the variant posterior inclusion probability (VPIP) that each variant is causal and the signal posterior inclusion probability (SPIP), the sum of the VPIPs for the variants in a region (Methods). This method can identify multiple independent signals in a region. In the 1,501 regions, we identified 2,435 signals with SPIP≥0.95, 1,952 of which are among the 2,030 association signals identified in conditional analysis. We built 95% credible sets^21^ of potential causal variants for the 1,952 signals (Fig. 3a; Supplementary Table 9). These credible sets included 1 to 544 variants (median=6); notably, 334 credible sets included only one variant (168 distinct variants).

**Figure 3:**
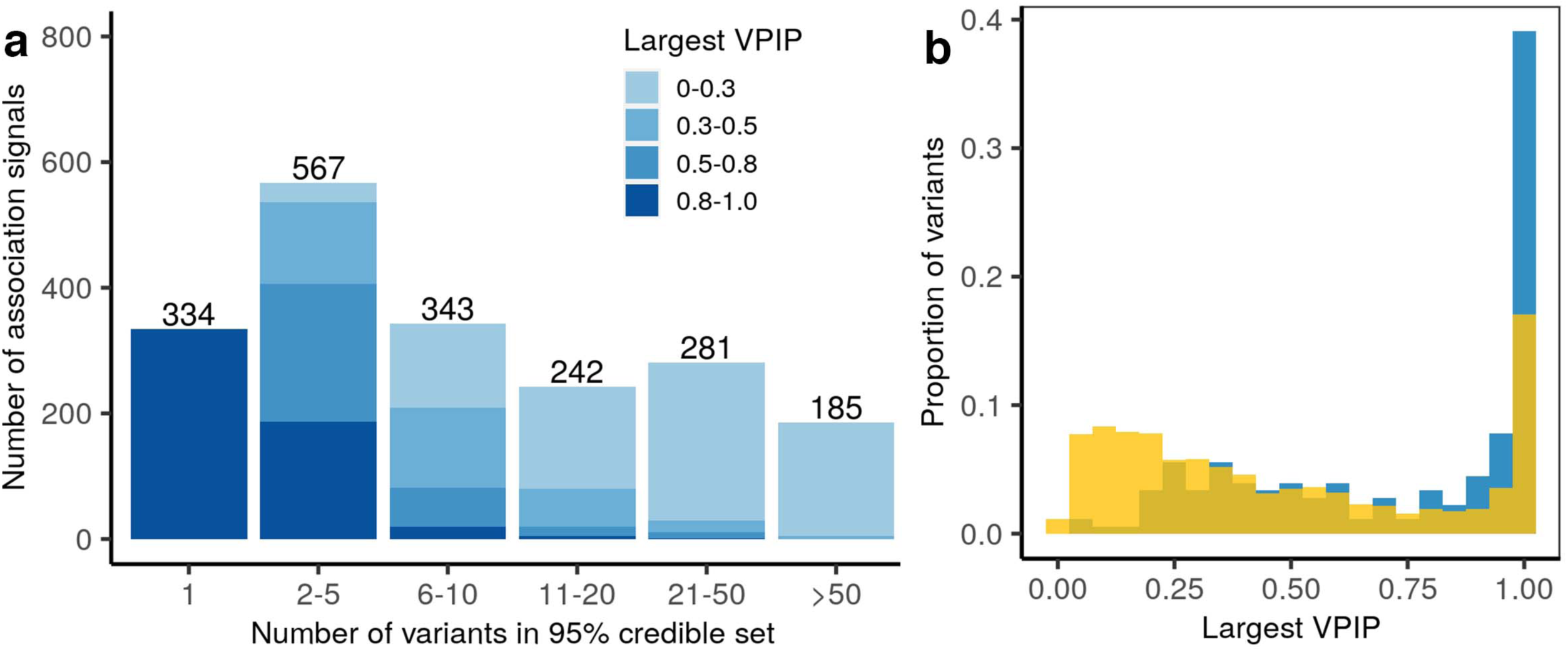
The 1,952 of the 2,030 metabolite genetic association signals identified in stepwise conditional tests with SPIP_≥_0.95 in DAP-g Bayesian fine mapping. a) Numbers of variants in the 95% credible sets and distribution of variant posterior inclusion probabilities (VPIPs) for the most likely causal variants within the 95% credible sets. b) Density plot of largest VPIPs highlights the variants with >10-fold greater frequency in METSIM than non-Finnish Europeans (gnomAD v3.1; blue) have larger VPIPs than all other variants (maize).

In each of the 1,952 credible sets, we identified the variant with the largest VPIP. This list comprised 1,119 distinct variants, 100 with MAF>10-fold greater in METSIM than in NFE. VPIPs for these 100 variants were greater than those for the remaining 1,019 (VPIP mean=0.73 vs. 0.47; t-test *P*=1.7×10^−21^; Fig. 3b).

Of the 1,119 variants, 263 had VPIP≥0.8 in 547 credible sets. Among these 263 variants, 46 are rare in METSIM and 47 have MAF>10-fold greater in METSIM than in NFE; 28 of these variants are both rare and enriched in Finns (Supplementary Table 9). The 263 variants include 11 protein-truncating (PTV) and 69 missense variants across 66 genes, and 183 other (mostly non-coding) variants (Supplementary Table 9). Given their likely impact on gene function, we focused on the 80=11+69 PTV and missense variants, which suggested causal roles for the corresponding 66 genes. These 80 variants had VPIP≥0.8 in credible sets for 208 signals with 173 metabolites. Among the 80 variants, 26 (5 PTV and 21 missense) are rare and 30 (6 PTV and 24 missense) have MAF>10-fold greater in METSIM than in NFE; 16 of these variants are both rare and enriched in Finns.

### Identifying novel association signals at rare and Finnish enriched variants

To determine which of the 2,030 association signals are distinct from previous metabolite GWAS findings, we repeated metabolite association analysis conditioning on all variants that were (a) ≤1 Mb of the index variant and (b) previously reported as associated with any metabolite in a curated list of 381 publications (Methods; Supplementary Table 10). 303 association signals at 229 index variants remained significant for 201 metabolites (*P*_*condition*_<7.2×10^−11^; Fig. 2b; Supplementary Table 7). The 303 novel signals included 64 signals (for 58 metabolites) at 51 rare variants and 79 signals (for 71 metabolites) at 47 variants >10-fold more frequent in METSIM than in NFE (Table 2); 33 of these signals are at variants both rare and enriched in Finns (Supplementary Table 7). In addition, 17 signals for 16 metabolites are on the X chromosome, and 78 signals are for 44 metabolites identified since 2015 on the Metabolon DiscoveryHD4 platform.

**Table 2:**
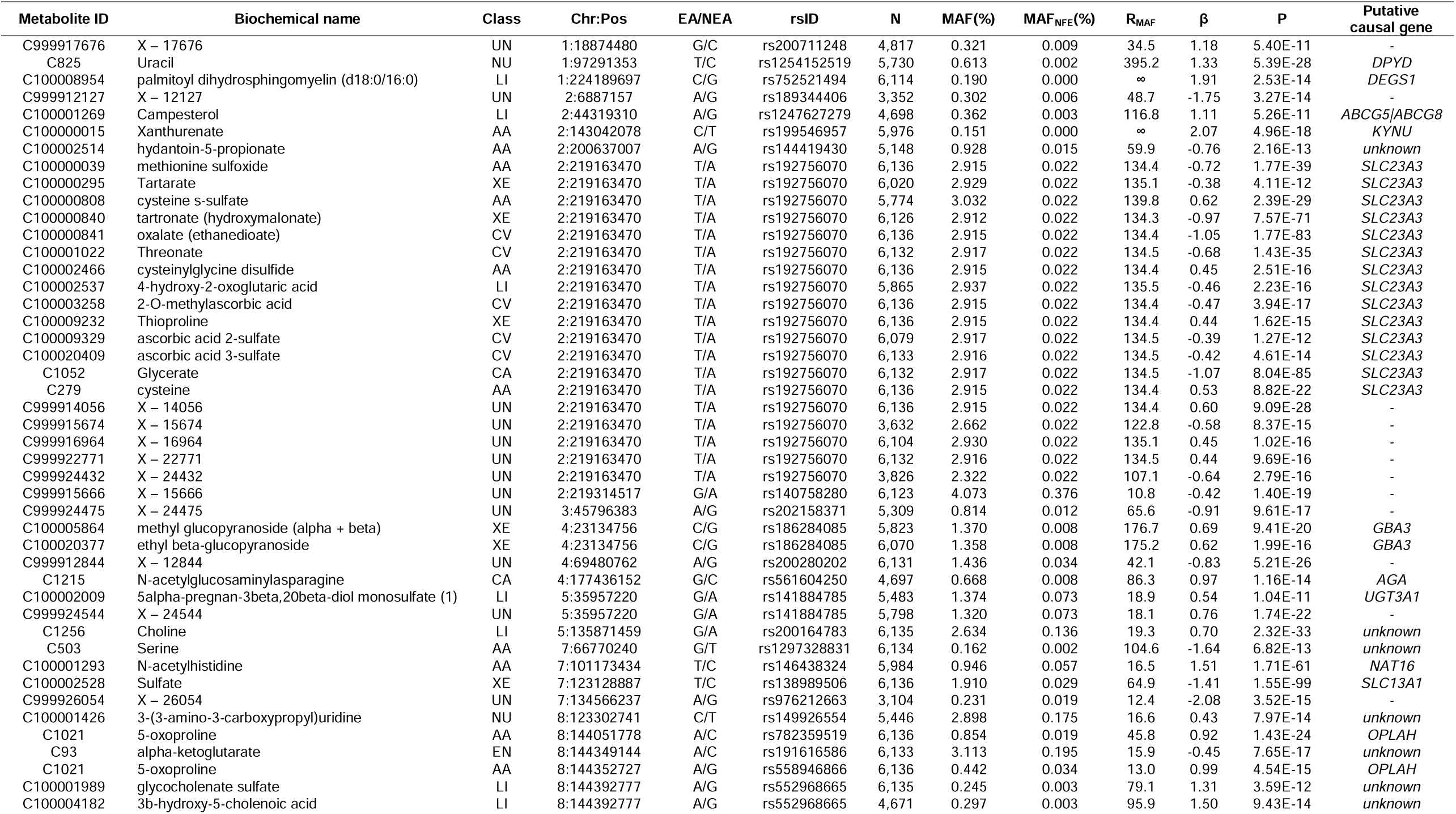
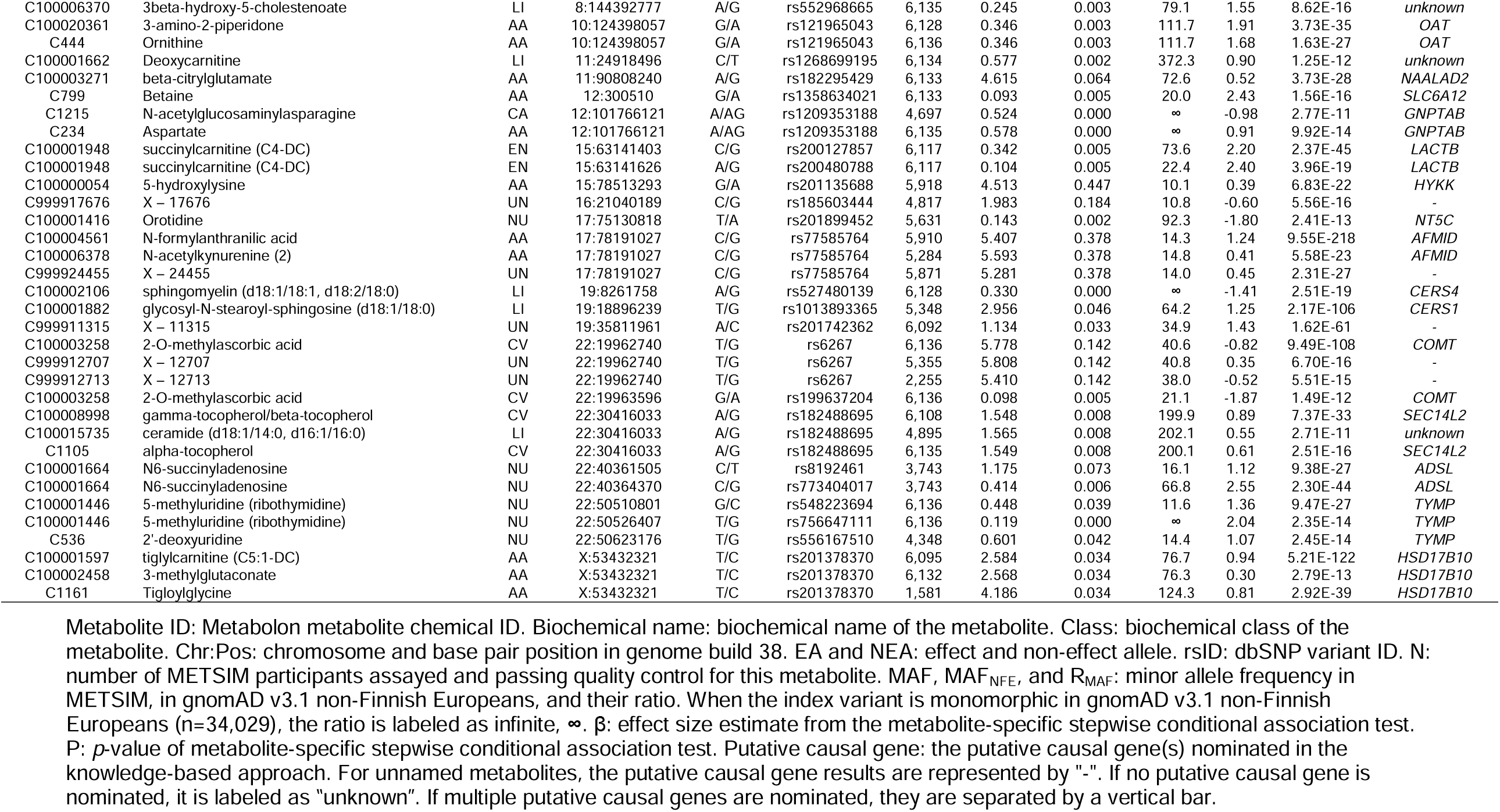
79 significant metabolite genetic association signals at 47 novel variants with MAF>10-fold greater in METSIM than in gnomAD v3.1 non-Finnish Europeans

Multiple novel associations arose at the same index variants. For example, we identified novel association signals with 19 metabolites at the putatively-deleterious *SLC23A3* missense variant p.Asn336Lys (rs192756070; Supplementary Fig. 8). p.Asn336Lys has 107-fold greater frequency in METSIM than in NFE (MAF=2.3% vs. 0.022%) and is likely the causal variant for most or all 19 metabolite associations (VPIP median=0.98, range=0.57-1.00). *SLC23A3* encodes an SLC23 ascorbic acid transporter without demonstrated nucleobase transport^22^. These novel associations suggest a wide range of transport functions for SLC23A3.

Among the novel association signals at index variants enriched in Finns, we identified an association with 3-amino-2-piperidone at the putatively-deleterious *OAT* missense variant p.Leu402Pro (rs121965043, β=1.91, *P*=3.7×10^−35^). p.Leu402Pro has 100-fold greater frequency in METSIM than in NFE (MAF=0.35% vs. 0.0031%) and is the likely causal variant for this association (VPIP=0.997). *OAT* encodes the key mitochondrial enzyme ornithine aminotransferase which converts arginine and ornithine into glutamate and gamma aminobutyric acid^23^. *OAT* has not previously been implicated in metabolite GWAS, but inactivation of OAT is responsible for the Finnish heritage disease gyrate atrophy characterized by hyperornithinemia^24^. Previous studies have found increased 3-amino-2-piperidone levels in the urine of individuals with gyrate atrophy^25^.

Among the novel association signals on the X chromosome, we identified an association for tiglylcarnitine at the putatively-deleterious *HSD17B10* missense variant p.Ala95Thr (rs201378370, β=0.94, *P*=5.2×10^−122^). p.Ala95Thr has 76-fold greater frequency in METSIM than in NFE (MAF=2.6% vs. 0.034%) and is the likely causal variant for this association (VPIP=0.997). *HSD17B10* encodes 17-β-hydroxysteroid dehydrogenase X, a mitochondrial enzyme which catalyzes oxidation of neuroactive steroids and degradation of isoleucine^26^. Mutations in *HSD17B10* that abolish enzyme activity lead to HSD10 deficiency, an infantile neurodegenerative disorder in which tiglylcarnitine level is elevated^27^. In contrast, *HSD17B10* is overexpressed in brains of individuals with Alzheimer’s disease^28^, in which tiglylcarnitine level is decreased^29^.

A previous study reported an association between the *GNPTAB* intronic variant rs7964859 and aspartate^15^. We identified an independent aspartate association signal with the *GNPTAB* frameshift variant p.Cys528ValfsTer19 (rs1209353188) (LD r^2^=0.01; β=0.91, *P*_condition_=5.2×10^−15^ conditioning on rs7964859). p.Cys528ValfsTer19 is rare in METSIM (MAF=0.58%), absent in gnomAD NFE, and is the likely causal variant for the METSIM aspartate association (VPIP=0.996). *GNPTAB* encodes the alpha- and beta-subunits of N-acetylglucosamine-1-phosphotransferase which catalyzes the N-linked glycosylation of asparagine residues with mannose-6-phosphate^30^.

### Nominating putative causal genes

To nominate putative causal genes for metabolite association signals, we applied two approaches. First, we nominated 66 putative causal genes for the 208 association signals for which fine-mapping analysis identified PTV or missense variants at VPIP≥0.8 (see Fine mapping). Second, we implemented a knowledge-based approach to integrate biological information about the metabolite and the 20 protein-coding genes nearest the corresponding index variant for the 1,666 of the 2,030 association signals with named metabolites (Methods). The knowledge-based approach nominated 215 single genes for 1,033 association signals with 480 metabolites (Supplementary Fig. 9) and 19 sets of 2 to 7 genes with similar biochemical activity (62 additional genes) for 324 association signals with 208 metabolites (Supplementary Tables 7, 11).

We compared gene nominations for the 138 metabolite association signals for which both approaches nominated causal genes. Compared to the fine-mapping analysis, the knowledge-based approach nominated the same gene for 119 signals, multiple paralogs including the same gene for 18, and a different gene for 1, for an overall consistency >99% (Supplementary Table 12).

The 277=215+62 genes identified by the knowledge-based approach and the 66 identified by fine mapping together comprised 290 genes. 204 (70%) of the 290 genes are the closest genes to the index variants, including 188 (68%) of the 277 genes identified by the knowledge-based approach. 58 of the 290 genes have not previously been implicated in metabolite GWAS (Fig. 2c). Of the 58 novel genes, 51 were identified by the knowledge-based approach (Box 1), 21 by fine-mapping analysis (Supplementary Table 9), and 14 by both. Of the 58 novel genes, 40 represent novel loci and 18 are within loci in which metabolite associations have previously been identified but the genes we nominated have not previously been implicated.

Novel genes nominated based on association signals with amino acid levels provide insight into how the encoded enzymes or transporters contribute to modifications of amino acid derivatives. As a first example, we identified a novel association at the *HDAC6* missense variant p.Arg832His (rs61735967) with N6-acetyllysine (MAF=2.9%, β=0.71, *P*=3.6×10^−80^) and suggested p.Arg832His is the likely causal variant (VPIP=0.998). Both the fine-mapping and knowledge-based approaches nominated *HDAC6* as the putative causal gene. *HDAC6* encodes a lysine deacetylase that removes the acetyl group from acetyllysine in histones. Increased *HDAC6* expression has been found in brains of individuals with Alzheimer’s disease^31^. Elevated levels of N6-acetyllysine were recently found in an Alzheimer’s disease mouse model^32^.

As a second example, we identified a novel association between the *QPCT* intronic variant rs77684493 and pyroglutamylglutamine (MAF=6.2%, β=-0.55, *P*=9.0×10^−31^). rs77684493 is in near-perfect LD (r^2^=0.996) with the putatively-deleterious *QPCT* missense variant p.Arg54Trp (rs2255991), which was also associated with pyroglutamylglutamine (β=-0.54, *P*=1.6×10^−30^) and has >7-fold greater frequency in METSIM than in NFE (MAF=6.3% vs. 0.89%). Our knowledge-based approach nominated *QPCT* as the putative causal gene for this association. *QPCT* encodes the enzyme glutaminyl-peptide cyclotransferase, which performs cyclization of the N-terminal glutamine residues and results in the pyroglutamine residue^33^. *QPCT* has been implicated in a schizophrenia GWAS^34^ and suggested as a druggable target for Huntington’s disease^35^.

GWAS of metabolites recently identified on the Metabolon platform helped nominate novel putative causal genes with high biochemical relevance in known metabolite-associated regions. For example, a previous study in a Japanese sample identified associations for blood creatinine and uracil levels at the *LRIG1* missense variant p.Thr792Met (rs202007714)^36^, which is monomorphic in METSIM and gnomAD Finns. We identified an association at the nearby (13 kb) *SLC25A26* missense variant p.Thr208Met (rs13874) with 2,3-dihydroxy-5-methylthio-4-pentenoate (DMTPA) (MAF=48.3%, β=0.17, *P*=2.3×10^−21^). We suggest *SLC25A26* as the causal gene for the DMTPA association. DMTPA, an S-adenosylmethionine, was recently identified on the Metabolon DiscoveryHD4 platform. SLC25A26 is the only known mitochondrial S-adenosylmethionine transporter.

Seven of the 58 novel genes were identified only by fine mapping. Among them, we identified a novel association for glycocholenate sulfate at the rare *ADCK5* missense variant p.Ala508Thr (rs552968665; β=1.31, *P*=3.6×10^−12^), which is >79-fold more frequent in METSIM than in NFE (MAF=0.25% vs. 0.0031%). Fine-mapping analysis suggested p.Ala508Thr is the likely causal variant (VPIP=0.89), implicating a causal role for *ADCK5. ADCK5* encodes the aarF domain containing kinase 5. These results suggest *ADCK5* plays a role in human bile acid metabolism.

### Colocalization of metabolites with human diseases

Integrating metabolite and disease genetic associations can improve fine-mapping resolution^37^ and clarify the potentially causal variants and disease genes. We performed Bayesian colocalization analysis^37,38^ based on the probabilistic fine-mapping results of METSIM metabolites and of 980 disease and disease-related dichotomous traits (henceforth, disease traits) in 176,899 Finns in FinnGen release 4 (Methods; Supplementary Table 13). We calculated the regional colocalization probability (RCP) of a putative causal variant shared between a METSIM metabolite and a FinnGen disease trait (Methods). We identified 946 colocalizations involving 248 metabolites and 105 interrelated disease traits (RCP≥0.5; Supplementary Table 14).

Integrating metabolite associations substantially increased the fine-mapping confidence of FinnGen disease trait association signals (SPIP median=0.91 vs. 0.73; paired t-test *P*=2.5×10^−70^) and the probability assigned to the most likely disease variant (maximum VPIP median=0.54 vs. 0.11; paired t-test *P*=9.1×10^−128^; Supplementary Fig. 10). For example, the putatively-deleterious *SERPINA1* missense variant p.Glu366Lys (rs28929474) was associated with N-acetylglucosaminylasparagine level in METSIM (MAF=2.3%, β=0.56, *P*=4.9×10^−16^) and risk of cholestasis of pregnancy in FinnGen (odds ratio (OR)=6.23, *P*=8.1×10^−17^). Our fine-mapping analysis suggested a causal role of *SERPINA1* p.Glu366Lys for N-acetylglucosaminylasparagine (VPIP=0.81). We detected colocalization in this region between signals for N-acetylglucosaminylasparagine and cholestasis of pregnancy (RCP=0.99). Colocalizing these association signals increased the SPIP for cholestasis of pregnancy from 0.69 to 0.99, and the VPIP of *SERPINA1* p.Glu366Lys from 0.37 to 0.80. *SERPINA1* encodes a serine protease inhibitor produced mainly in the liver. *SERPINA1* mutations have been associated with familial intrahepatic cholestasis^39^, and *SERPINA1* p.Glu366Lys with liver diseases^40^ and circulating liver enzymes^41^.

### Campesterol and gallstones: potential causal link

Gallstones affect 10-20% of adults worldwide^42^. Aberrant cholesterol homeostasis, particularly the physical-chemical imbalance of cholesterol solubility in bile, induces gallstones^43^. Blood campesterol levels have been associated with gallstones^44^, but it is uncertain whether the relationship is causal. We identified associations at the *ABCG8* intronic variant rs6544713 with lower campesterol level in METSIM (MAF=20.1%, β=-0.33, *P*=2.7×10^−37^) and higher gallstone risk in FinnGen (OR=1.32, *P*=8.0×10^−65^). In 4,689 METSIM participants with observed campesterol levels, 199 with gallstones, plasma campesterol level was inversely associated with gallstone risk (β=-0.52, *P*=3.7×10^−5^).

Colocalization analysis suggested campesterol and gallstones share the same causal variant in this region (RCP=0.65; Fig. 4a) and nominated rs6544713 (SCP=0.45) as the most likely causal variant. rs6544713 resides in active regulatory units in intestinal tissue^45^ and its campesterol-decreasing allele is associated with higher *ABCG8* expression in colon tissue^46^ (β=0.37, *P*=7.2×10^−16^), but not in the liver^46^. *ABCG8* and its nearby paralog *ABCG5* have previously been suggested as candidate genes for gallstones^47^. Mutations in *ABCG5* and *ABCG8* cause sitosterolemia, characterized by elevated campesterol^48^.

**Figure 4:**
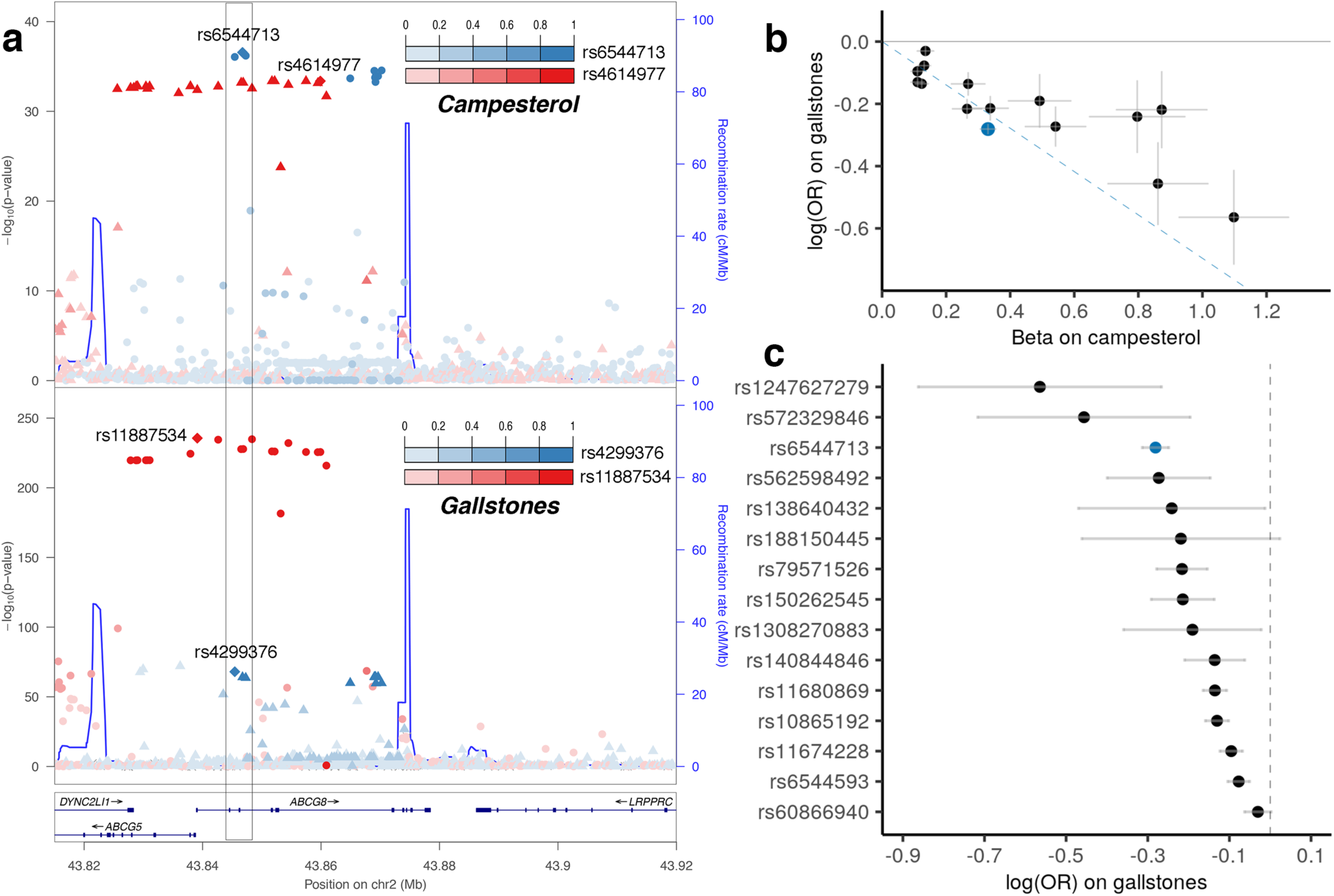
Colocalization and causal relationship between campesterol and gallstones. a) Stacked regional association plots for campesterol and gallstones (cholelithiasis, K11_CHOLELITH in FinnGen release 4) in the *ABCG5/ABCG8* region. The index variants identified in stepwise conditional analysis (campesterol) and approximate conditional analysis (gallstones) are labeled and variants colored by their linkage disequilibrium (LD) to the index variant with which they are in strongest LD in METSIM. The campesterol signal (index variant rs6544713) is colocalized with the gallstone signal (rs4299376, pairwise LD r^2^=0.993, RCP=0.65) shown in the gray box. In contrast, no colocalization was detected between the signals indexed by rs4614977 and rs11887534. No coding variants within 1 Mb have LD r^2^>0.2 with rs6544713 in METSIM. b) Comparison of effect sizes for the 15 instrumental variables genome-wide without significant heterogeneity (*P*>0.05) used in Mendelian randomization analysis between campesterol and gallstones. rs6544713 is in blue. The slope of the blue dashed line depicts the estimated causal effect size of campesterol on gallstones. The Egger regression intercept is deemed not significant (*P*=0.15). c) Negative relationship between instrumental variable and risk of gallstones. OR: odds ratio.

Mendelian randomization analysis using 15 independent variants for campesterol suggested a causal effect of lower plasma campesterol level on higher gallstone risk (Methods; β=-0.70, *P*=7.2×10^−8^; Fig. 4b-c). Interestingly, we also identified a causal effect of gallstones on lower plasma campesterol level (β=-0.49, *P*=1.3×10^−39^) in Mendelian randomization analysis using 151 independent variants. *ABCG5* and *ABCG8* together encode a heterodimeric ATP-binding cassette transporter that facilitates secretion of cholesterol and non-cholesterol sterols in the intestine and bile. High plasma campesterol levels might compete with cholesterol for ABCG5/ABCG8 transporters during biliary cholesterol secretion, resulting in decreased biliary cholesterol levels and reduced risk of gallstones^49,50^ (Supplementary Fig. 11).

### *DBH* influence on vanillylmandelate and hypertension: distinct pathways

Stepwise conditional analysis identified the putatively-deleterious *DBH* missense variant p.Arg79Trp (rs77273740) as associated with lower vanillylmandelate (β=-0.38, *P*=1.8×10^−15^); p.Arg79Trp is >10-fold more frequent in METSIM than in NFE (MAF=4.5% vs. 0.39%). Both fine-mapping and the knowledge-based approach suggested a causal role for *DBH*. The knowledge-based approach suggested DBH could exhibit an effect on vanillylmandelate in two ways (Supplementary Fig. 11): by converting dopamine to norepinephrine, a vanillylmandelate precursor^51^, or by transforming homovanillate acid to vanillylmandelate through hydroxylation^52^.

In FinnGen, the *DBH* p.Arg79Trp vanillylmandelate-decreasing allele was significantly associated with lower hypertension risk (OR=0.84, *P*=5.2×10^−13^), consistent with previous associations with systolic and diastolic blood pressures^53,54^. No other variants were associated with FinnGen hypertension or METSIM vanillylmandelate in this region (*P*>10^−6^; Supplementary Fig. 12). In 5,173 METSIM participants with observed vanillylmandelate levels, 1,233 with hypertension, plasma vanillylmandelate level was associated with hypertension (β=0.52, *P*=3.2×10^−9^).

Colocalization analysis suggested that hypertension colocalized with vanillylmandelate (RCP=0.996). However, Mendelian randomization found no evidence for a causal effect of vanillylmandelate on hypertension (*P*=0.15; 10 independent variants; Supplementary Fig. 12) or hypertension on vanillylmandelate (*P*=0.17; 157 independent variants), suggesting this signal conferred effects on hypertension risk and vanillylmandelate through different pathways, consistent with the two possible DBH effects identified by our knowledge-based approach (Supplementary Fig. 11).

## Discussion

We performed GWAS of 1,391 plasma metabolites in 6,136 men from the late-settlement region of Finland. We sought to identify putative causal variants and genes for the resulting genetic associations, and interrogated disease molecular mechanisms by integrating metabolite and disease genetic associations. We identified 2,030 association signals for 803 metabolites, including 157 signals for 125 metabolites at 121 rare variants. We identified 303 association signals for 201 metabolites as novel, including 64 signals for 58 metabolites at 51 rare variants.

Over half of these 303 novel association signals stem from the population history of Finland, the analysis of previously-unstudied metabolites, or the analysis of the X chromosome. The Finnish population history of alternating founding events and population expansions has resulted in a set of genetic variants rare elsewhere but more common in Finns, providing increased statistical power for genetic discovery for these variants^2^, as exemplified by the Finnish heritage diseases^55^. 79 of the 303 novel association signals we identified are at 47 variants with MAF>10-fold greater in METSIM than in NFE, with 37 novel signals at 14 variants with MAF>100-fold greater. These include the novel association of 3-amino-2-piperidone with the rare *OAT* missense variant p.Leu402Pro; mutations in *OAT* cause the Finnish heritage disease gyrate atrophy (see Results).

Metabolon continues to expand the set of metabolites identified on their platform. 78 of the 303 novel association signals were for 44 metabolites identified after 2015 on the Metabolon DiscoveryHD4 platform, and so studied only in the most recent Metabolon-based metabolomics GWAS^13^. For example, we identified a novel association at *SLC23A3* missense variant p.Asn336Lys for 2-O-methylascorbic acid, identified on the Metabolon platform in 2019.

Our study is one of the first Metabolon metabolomics GWAS to analyze the X chromosome, where 17 of the 303 novel association signals arose. For example, we identified a novel association for tiglylcarnitine at the *HSD17B10* missense variant p.Ala95Thr. *HSD17B10* mutations cause a rare inborn error of metabolism characterized by cognitive impairment and variable neurological abnormalities.

Biochemical analysis existed for decades prior to the advent of GWAS. Experiments linking a gene to a metabolite often already existed in the published literature. We identified 277 putative causal genes through existing links in the literature between tested metabolites and biochemical activities of genes near our association signals. Our results suggested most of these putative causal genes acted on the associated metabolites or closely-related metabolites. These putative causal genes characterized the genetic regulatory mechanisms for plasma metabolite levels. The associations of multiple metabolites with the same gene help improve the understanding of the gene function. For example, we nominated *SLC23A3* as a causal gene for 19 metabolites of various biochemical classes, suggesting a wide range of transport functions in addition to its known role as an ascorbic acid transporter.

Integrating metabolite and disease genetic associations helps disentangle disease biology. We identified 946 metabolite-disease trait pairs likely sharing the same causal variants, which helped pinpoint the likely causal variants and disease genes (Supplementary Table 14). For example, colocalization analysis of acetylglucosaminylasparagine and cholestasis suggested a shared causal role of *SERPINA1* p.Glu366Lys. Mendelian randomization analysis suggested for the first time a protective effect of high plasma campesterol on gallstones. Plasma campesterol is commonly used as a biomarker for gallstones^56^ and campesterol is used as a supplement to reduce low-density lipoprotein cholesterol^57^. Our finding provides supporting evidence for these applications of campesterol in the treatment of gallstones.

Data sharing increases the impact of genetic studies. To support data exploration of our metabolite GWAS results^58^, we have constructed a METSIM metabolite PheWeb site^59^ (Fig. 1). This site supports querying, visualizing, and downloading our METSIM Metabolon metabolite genetic association results, including Manhattan and quantile-quantile plots, and summary statistics for all 1,391 metabolites. In addition, we provide direct links to the Human Metabolome Database (HMDB)^60^, which presents the metabolites’ biochemical characteristics and enables interpretation of metabolite genetic association results.

In summary, we performed parallel GWAS for 1,391 plasma metabolites in 6,136 adult Finnish males from the METSIM study, colocalized metabolite and disease genetic associations, and made these GWAS results available using PheWeb. Our findings reveal genetic determinants for a wide range of plasma metabolites and demonstrate the utility of metabolite genetic associations for the investigation of disease biology.

## Methods

### METabolic Syndrome In Men (METSIM) study

METSIM is a study of 10,197 Finnish men from Kuopio in the late-settlement region of northeast Finland designed to investigate factors associated with type 2 diabetes and cardiovascular diseases^3^ (Supplementary Table 3). Participants were aged 45 to 74 (median=58) years during baseline visits from 2005-2010. Participants provided demographic, diet, exercise, disease, and medication history information, and underwent laboratory measurements, including oral glucose tolerance test, after ≥10-hour overnight fast. Morbidity, mortality, and drug treatment information was periodically updated for participants who consented to use of their hospital admission, drug reimbursement, and prescription records in Finnish national registries. Due to funding constraints, we randomly selected 6,490 of the 8,777 METSIM participants who at baseline were neither diagnosed with diabetes nor taking diabetes medications that might broadly impact metabolomics levels for the Metabolon metabolomics assay. After exclusion of participants who subsequently developed diabetes (n=264), lacked array genotypes (n=65) or body mass index (BMI) measurement (n=1), had sex mismatch (n=3), and/or were non-Finnish (n=21), our analysis set comprised 6,136 participants (Supplementary Table 3; Supplementary Fig. 1). This study was approved by the Ethics Committee at the University of Eastern Finland and the Institutional Review Board at the University of Michigan. All participants provided written informed consent.

### Metabolomics profiling and data processing

Non-targeted metabolomics profiling was performed at Metabolon, Inc. (Durham, North Carolina, USA)^61^ on EDTA-plasma samples obtained after ≥10-hour overnight fast during METSIM baseline visits. Briefly, methanol extraction of biochemicals followed by a non-targeted relative quantitative liquid chromatography–tandem mass spectrometry (LC-MS/MS) Metabolon DiscoveryHD4 platform was applied to assay named (n=1,240) and unnamed (n=304) metabolites (Supplementary Tables 1-2). Samples were randomized across batches.

Batches contained ∼144 METSIM samples and 20 well-characterized human-EDTA plasma samples for quality control (QC). All 6,490 samples were processed together for peak quantification and data scaling. We quantified raw mass spectrometry peaks for each metabolite using the area under the curve. We evaluated overall process variability by the median relative standard deviation for endogenous metabolites present in all 20 technical replicates in each batch. We adjusted for variation caused by day-to-day instrument tuning differences and columns used for biochemical extraction by scaling the raw peak quantification to the median for each metabolite by Metabolon batch.

### Array genotyping and exome sequencing

All METSIM participants were array genotyped on the Human OmniExpress-12v1_C BeadChip (OmniExpress) and Infinium HumanExome-12 v1.0 BeadChip (exome array) platforms^62^. We excluded individuals for sex or relationship mismatch, apparent sample duplication, or ancestry outliers based on genetic principal component analysis (PCA). We removed variants with genotype call rate <95% (OmniExpress) or <98% (exome array), or Hardy-Weinberg equilibrium (HWE) *P*<10^−6^ (either array)^62^.

We captured exomes for all METSIM participants by SeqCap EZ HGSC VCRome kit (Roche) and sequenced them by HiSeq2000 (Illumina) (average depth 45x)^2^. For exome sequences, we excluded samples with estimated contamination >3% or sample swaps compared to the array genotype data^62^ and required single-nucleotide variant (SNV) array genotype concordance >90% if array data were available. We filtered variants with genotype call rate <98%, HWE *P*<10^−6^, or overall low allele balance (alternate allele count/sum of total allele count <30%)^2^. The resulting array-genotype dataset consisted of n=10,066 METSIM participants with 679,866 SNVs. The exome-sequence dataset consisted of n=9,957 participants with 583,947 SNVs and 40,270 small insertions/deletions (indels).

### Genome sequencing

We whole genome sequenced METSIM participants in two waves. In wave 1, we genome sequenced 3,074 METSIM participants (average depth 23x)^63^. Genomic DNA was fragmented on a Covaris LE220 instrument and size-selected to ensure an average insert size of 350-375 base pairs (bp). Libraries were constructed with the Illumina TruSeq or KAPA Hyper PCR-free library prep kit. qPCR was used to determine concentration of each library. Libraries were subsequently pooled and sequenced with 2×150 bp paired-end reads using HiSeq X (Illumina). We filtered read alignments with mismatch rate ≥5%, inter-chromosomal rate ≥5%, discordance rate of paired reads ≥5%, or haploid coverage <19.5x. We generated QC statistics in Picard v2.4.1 (http://broadinstitute.github.io/picard/), Samtools v1.3.1^64^, and VerifyBamID v1.1.3^65^. We called SNVs and small indels and performed base quality score recalibration in GATK v3.5 (https://gatk.broadinstitute.org/). We excluded variants with missingness >2%, HWE *P*<10^−6^ in unrelated individuals, or allele imbalance <30%. The resulting genome sequence consisted of n=3,074 participants genotyped for 23,849,428 SNVs and 2,914,167 indels. We used wave 1 as part of our imputation reference panel (see METSIM integrative panel and genotype imputation).

In wave 2, we sequenced 2,875 additional METSIM participants using the same methods used for wave 1. We generated a combined wave 1+2 call set of n=5,949 using the same methods, resulting in calls for 55,648,111 SNVs and 12,850,837 indels. Wave 2 data became available only after the main analysis for this paper was complete; we used wave 1+2 combined data to determine linkage disequilibrium (LD) proxies for previously-identified metabolite associated variants that were missing in wave 1 but present in wave 1+2 combined data (see Identification of novel associations).

### METSIM integrative panel and genotype imputation

Using the 3,074 METSIM participants with wave 1 genome sequence data, we generated an integrated list of genetic variant sites by merging site lists from the genome and exome sequence data, and the OmniExpress and exome array data. Of the 3,074 participants, 3,055 had OmniExpress and exome array data, and 3,037 had exome sequence data. We calculated genotype likelihoods for each individual at each site as the product of genotype likelihoods assuming independent data across platforms^66^. For OmniExpress and exome array genotypes, we converted genotype calls to genotype likelihoods assuming a genotype error rate of 10^−6^. We then phased genotypes using integrated genotype likelihoods in Beagle v4.1 with 50,000 markers per chunk and 3,000 overlapping genetic markers between consecutive chunks^67^. We subsequently excluded 1 individual who self-identified as non-Finnish, 2 individuals identified as population outliers in genetic PCA, and 149 close relatives (estimated kinship ≥0.125 in KING v2.2.1^68^). The resulting integrative panel comprised 2,922 individuals genotyped for 23,294,337 SNVs and 2,851,848 indels (Supplementary Table 4). 2,670 (91.4%) of the 2,922 individuals had Metabolon metabolomics data.

We imputed genotypes for the 6,490 study participants on the framework of their OmniExpress genotypes using the METSIM integrative panel with Minimac v4^69^. We excluded imputed variants with imputation r^2^<0.3, leaving 19,182,997 SNVs and 2,404,717 indels for downstream analysis (Supplementary Table 5).

### Variant functional annotation

We annotated all variants using the Ensembl Variant Effect Predictor (VEP) version 99^70^. We used the “-pick_order” option to annotate each variant using a single transcript, with transcripts prioritized in the following order: transcript support level (i.e. well-supported and poorly-supported transcript models based on the type and quality of the alignments used to annotate the transcript), transcript biotype (protein coding preferred), APPRIS isoform annotation (i.e. annotation based on a range of computational methods to identify the most functionally important transcripts from cross-species conservation), deleteriousness of annotation as estimated by Ensembl, transcript CCDS status (i.e. amount and type of evidence that supports the existence of a variant), canonical status of transcript (https://m.ensembl.org/Help/Glossary), and transcript length^71^. We used the dbNSFP (version 4.0)^72^ plugin to generate additional predictions of variant deleteriousness from five *in silico* algorithms: Polyphen2 HDIV^73^, Polyphen2 HVAR^73^, SIFT4G^74^, MutationTaster^75^, and the Likelihood Ratio Test (LRT)^76^.

### Trait transformatio

For each metabolite, we inverse normalized the Metabolon-reported metabolite level, regressed on covariates (age at sampling, Metabolon batch, and lipid-lowering medication use status for lipid traits only), and inverse normalized the residuals. In the single-variant association analyses with BMI adjustment, we also included BMI among the covariates.

### Single-variant association analysis

We carried out single-variant association tests using a linear mixed model in EPACTS (v3.2.6) (https://github.com/statgen/EPACTS) on the normalized residual metabolite values. We limited analysis to the 1,391 metabolites successfully measured on ≥500 METSIM participants and to the genetic variants with minor allele count (MAC)≥5. This resulted in 10,914,098 to 16,172,108 variants (median=16,042,879) tested across the 1,391 metabolites, since the number of variants with MAC≥5 varied with the set of individuals successfully measured for each metabolite.

To choose a study-wise significance threshold for the 1,391 parallel metabolite GWAS, we carried out PCA across the metabolites to determine the number of principal components required to explain metabolite variation. To account for missing data (Supplementary Fig. 13), we first imputed missing metabolite values using the K-nearest neighbors approach^77^ with K=5. PCA of the imputed data showed that 692 principal components explained 95% of phenotypic variation for the 1,391 metabolites. We therefore used a study-wise significance threshold of *P*<5.0×10^−8^/692=7.2×10^−11^ for our single-variant analyses. A metabolite quantified in n=500 participants provided >80% power at *P*<7.2×10^−11^ to detect variants that explained phenotypic variance≥11% and had MAC≥5.

### PheWeb browser

We built a PheWeb browser^59^ of the 1,391 metabolite GWAS to support interactive visualization, exploration, and download of these results. This PheWeb (https://pheweb.org/metsim-metab/) includes Manhattan and quantile-quantile plots, summary statistics, and links to biochemical characteristics and functions in the Human Metabolome Database (HMDB)^60^ for all 1,391 metabolites.

### Stepwise conditional analysis

We carried out stepwise conditional analysis in EPACTS (v3.2.6) (https://github.com/statgen/EPACTS) to identify near-independent association signals. For each metabolite-chromosome pair with at least one single-trait genome-wide significant association (*P*<5.0×10^−8^), we first conditioned on the most significant associated variant and continued conditioning on the most significant remaining variant until no variant attained *P*<5.0×10^−8^.

### Fine mapping and credible sets

For each of the 2,030 nearly-independent association signals, we built genomic regions of 1 Mb on either side of the index variant, less near chromosome ends. We merged overlapping regions for the same metabolite, resulting in 1,501 genomic regions of 1.2 to 3.1 Mb. To identify potential causal variants within each region, we performed fine-mapping analysis using the Deterministic Approximation of Posteriors (DAP-g) method^20^ (https://github.com/xqwen/dap), assigning equal priors to all candidate variants. DAP-g uses individual-level metabolite, genotype, and covariate data to produce fine-mapping results. Since DAP-g does not allow for related participants, we corrected for relatedness approximately by including the first ten genetic principal components as covariates; repeating the DAP-g analysis with 0, 20, or 100 principal components yielded similar results.

DAP-g allows for multiple independent association signals within each region. For each identified signal, DAP-g computes (1) a signal posterior inclusion probability (SPIP) that there is at least one causal variant in the signal; and (2) a posterior inclusion probability for each variant (VPIP) that the variant is causal for the signal. For each of the 1,952 signals identified in stepwise conditional tests that had SPIP≥0.95 in DAP-g, we constructed a 95% credible set of potential causal variants by ranking the variants in descending VPIP and including variants until their summed VPIP was ≥0.95.

### Identification of novel associations

To assess which of our metabolite associations were novel, we compiled a list of 381 published metabolite GWAS papers (Supplementary Table 10): 354 from the NHGRI-EBI GWAS catalog^78^ (https://www.ebi.ac.uk/gwas/; release date December 1, 2020); and 27 others from the list curated by Kastenmüller et al.^11^ (accessed April 1, 2021). From these papers, we identified 8,502 variants with metabolite associations at *P*<5.0×10^−8^ or at the significance threshold used in the paper, whichever was more stringent (Supplementary Table 10). Among these 8,502 published variants, 7,807 were present in the METSIM imputed genotype data. For 194 of the published variants not present in the METSIM imputed genotype data, we identified proxies (LD r^2^≥0.8 and ≤500 kb) using the wave 1+2 genome sequence dataset of 5,949 METSIM participants. The 7,807 variants present in the METSIM imputed genotype data and the 194 LD proxies for missing variants together comprised 8,000 unique variants. To avoid problems with multicollinearity, we pruned these 8,000 variants at METSIM LD r^2^>0.99 and ≤1 Mb, yielding 6,501 LD-pruned variants. Then, for each of the 2,030 association signals, we repeated the conditional association analysis including the subset of these LD-pruned variants within ≤1 Mb of the corresponding index variant as covariates. We considered as novel signals those index variants with conditional *P*<7.2×10^−11^ and location >500 kb from any of the 8,502-7,807-194=501 published variants, which were neither present nor with proxies in the METSIM imputed genotype data. Among the 501 variants, 380 were monomorphic in gnomAD v3.1 Finns (n=5,316).

### Knowledge-based approach to gene nomination

To nominate putative causal genes for the 1,666 of 2,030 signals associated with named metabolites, we employed a two-stage knowledge-based approach^15^. In stage 1, for each variant, we identified the 20 closest protein-coding genes using the minimum distance from the index variant to the refSeq genes’ transcription start or end sites. We employed an algorithm to look for lexical overlaps between the associated metabolite and each of the 20 genes.

Specifically, we searched automatically for matching strings using customized scripts between: (1) the HMDB^60^ metabolite name and synonyms and Entrez gene names^79^; (2) the metabolite and Entrez gene names listed in HMDB as interacting with the metabolite; (3) the metabolite name and Uniprot protein names^75^ and their synonyms; (4) the metabolite and its parent classes as defined in HMDB and the Uniprot protein names and their synonyms; (5) the metabolite name and rare disease names linked to each gene in OMIM (Online Mendelian Inheritance in Man, https://omim.org/, accessed January 1, 2021) after removing the non-specific substrings uria, emia, deficiency, disease, transient, neonatal, hyper, hypo, defect, syndrome, familial, autosomal, dominant, recessive, benign, infantile, hereditary, congenital, early-onset, idiopathic; (6) the metabolite and its parent classes and Gene Ontology (GO) biological process names^80^ associated with each gene after removing the non-specific substrings metabolic process, metabolism, catabolic process, response to, positive regulation of, negative regulation of, regulation of (we only considered gene sets of <500 genes); and (7) Kyoto Encyclopedia of Genes and Genomes (KEGG)^81^ maps (https://www.kegg.jp/) containing the metabolite (as defined in HMDB) and KEGG maps containing each gene (as defined in KEGG) omitting the large “metabolic process map”. For each of these pairs of terms, we calculated a Pair Distance score ranging from 0 to 1 using the Ruby gem “fuzzy_match” (https://github.com/seamusabshere/fuzzy_match), and considered a score >0.5 as a match.

In stage 2, we manually reviewed the evidence collected at stage 1. We selected the biologically most plausible causal gene if we identified experimental evidence linking the gene to the metabolite. >1 putative causal genes could be nominated if >1 gene was suggested in stage 1 and/or 2; this happened most often when a locus contains multiple paralogs with similar biochemical activity. If no clear experimental evidence existed for any of the 20 genes, no causal gene was selected.

### Colocalization of FinnGen disease traits and METSIM metabolites

To identify shared causal variants between METSIM metabolites and FinnGen disease traits, we carried out Bayesian pairwise colocalization analysis using fastENLOC^37,38^ (https://github.com/xqwen/fastenloc). We downloaded FinnGen release 4 (https://www.finngen.fi/en/access_results) FINEMAP^82^-based fine-mapping results for 980 disease traits with at least one association at *P*<5.0×10^−8^. fastENLOC used these FinnGen fine-mapping results and our DAP-g-based fine-mapping results for METSIM metabolites to carry out colocalization analysis assuming a single causal variant. For each FinnGen disease trait, we estimated its degree of enrichment for genome-wide associations in metabolite GWAS using TORUS^83^ (https://github.com/xqwen/torus) and used this enrichment estimate as the prior for Bayesian analysis in fastENLOC. fastENLOC computes two probabilities. The regional colocalization posterior probability (RCP) is the probability of the same causal variant within a region for both the metabolite and the FinnGen disease trait. The variant colocalization posterior probability (SCP) is the probability a specific variant is causal for both traits. We limited colocalization analysis to the 1,952 metabolite stepwise association signals with SPIP≥0.95 for 792 metabolites and present colocalizations for metabolite-FinnGen disease trait pairs with RCP≥0.5 (Supplementary Table 14).

### Associations of campesterol with gallstones and vanillylmandelate with hypertension in METSIM

Among the 4,698 METSIM participants with measured plasma campesterol level at baseline, we identified 199 with gallstones in METSIM (December 2020). To test for association between plasma campesterol level and presence of gallstones, we used logistic regression with covariates baseline study age, Metabolon batch, and lipid medication use. Among the 5,173 METSIM participants with measured plasma vanillylmandelate level at baseline, we identified 1,233 individuals with hypertension, and used logistic regression with covariates baseline study age, Metabolon batch, and hypertension medication use to test for association between plasma vanillylmandelate level and hypertension status.

### Causal effects between metabolites and FinnGen disease traits

To infer the potential causal effects of plasma campesterol on FinnGen gallstones (phenocode: K11_CHOLELITH) and plasma vanillylmandelate on hypertension (phenocode: I9_HYPTENS), we applied four two-sample Mendelian randomization methods: inverse variance weighted^84^, weighted median^85^, MR-PRESSO^86^, and MR-Egger^87^. These methods make different assumptions and use different strategies to account for horizontal pleiotropy, which can result in false positive inference of causality. For each metabolite, we identified nearly-independent genetic instrument variables (LD r^2^<0.1, distance≥500 kb) with unconditional single-variant association *P*<10^−6^. We also ran Mendelian randomization analyses to infer the causal effect of FinnGen gallstones (phenocode: K11_CHOLELITH) on campesterol and FinnGen hypertension (phenocode: I9_HYPTENS) on vanillylmandelate in a similar way. We considered findings significant if they had the same effect direction and *P*<0.05 for all four Mendelian randomization methods. We present MR-PRESSO effect estimate and *p*-values in the main text.

## Data Availability

The exome sequencing and genotyping array data can be accessed through dbGaP (https://www.ncbi.nlm.nih.gov/gap/) with accession numbers phs000752 and phs000919, respectively. Complete summary statistics from the genome-wide association studies of the 1,391 plasma metabolites are available and downloadable at https://pheweb.org/metsim-metab/. FinnGen genome-wide summary statistics and Bayesian statistical fine-mapping results are available at https://r4.finngen.fi. Individual-level whole genome sequencing and mass spectrometry data from METSIM are not available to outside researchers due to privacy restrictions.

## Acknowledgements

We thank the participants in the METSIM and FinnGen studies and the FinnGen study investigators. We thank Hyun Min Kang and Matthew W. Mitchell for their expertise and consultation in the genotype integration and metabolomics data processing. This work was supported by the National Institutes of Health (NIH) under awards U01 DK062370 (M.B., A.E.L.), R35 GM138121 (X.Q.W.), R01 DK119380 (X.Q.W.), R01 GM124061 (J.Kang, E.C.H., D.W.Z.), R01 DA048993 (J.Kang), R01 MH105561 (J.Kang), P01 HL151328 (N.O.S), R01 HL131961 (N.O.S), UM1 HG008853 (I.H., N.O.S., L.G., A.E.L.), R01 DK093757 (K.L.M.), U01 DK105561 (K.L.M.), T32 HL007081 (E.Y.), and UL1 TR002345 (E.Y.). X.Y.Y. was supported by an American Diabetes Association Postdoctoral Fellowship (1-19-PDF-061) and a University of Michigan Precision Health Scholarship. M.L. was supported by the Academy of Finland (grant no. 321428) and the Sigrid Juselius Foundation. S.R. was supported by the Academy of Finland Center of Excellence in Complex Disease Genetics (grant no. 312062 and 336820), the Finnish Foundation for Cardiovascular Research, the Sigrid Juselius Foundation, University of Helsinki HiLIFE Fellow and Grand Challenge grants, and Horizon 2020 Research and Innovation Programme (grant no. 101016775 “INTERVENE”). A.P. was supported by the Academy of Finland Center of Excellence in Complex Disease Genetics (grant no. 312074 and 336824). F.S.C., M.R.E., and L.L.B were supported by the NIH Intramural Research Program of the National Human Genome Research Institute (ZIA HG000024).

## Author contributions

M.B., M.L., and E.B.F. supervised experiments and analyses. X.Y.Y., F.S.C., K.L.M., X.Q.W., L.J.S., E.B.F., M.L., and M.B. designed the study. A.U.J., A.E.L., C.F., H.M.S., K.T.Y., S.K.S., E.Y., L.G., I.H., I.D., M.R.E., L.L.B., N.O.S., and H.A. produced and quality-controlled the genotype and sequence data. M.L., L.F.S., and J.K. enrolled the study participants. M.L., X.Y.Y., L.F.S., J.Kang, C.F.B., and G.R.W collected, quality-controlled and/or prepared the metabolomics data for association analysis. X.Y.Y., A.U.J., H.M.S., E.B.F., L.S.C., D.B., D.W.Z., E.C.H., and J.M. analyzed data. P.V. designed and created the PheWeb site. S.R. and A.P. are principal investigators of the FinnGen study. M.L. is the principal investigator of the METSIM study. X.Y.Y., M.B., M.L., E.B.F., A.U.J., X.Q.W., and N.B.F. wrote the manuscript draft. All authors contributed to the interpretation of results and critically reviewed the manuscript.

## Box 1 Primary supportive biochemical evidence for the 51 novel putative causal genes nominated in the knowledge-based approach

***ADSL*** (N6-succinyladenosine, NU, rs773404017): *ADSL* encodes adenylosuccinate lyase which converts adenylosuccinate to adenosine monophosphate. N6-succinyladenosine is the dephosphorylated version of adenylosuccinate. Rare loss of function of this enzyme results in elevated circulating N6-succinyladenosine levels (PMID: 5432795).

***ALDH4A*1** (S-1-pyrroline-5-carboxylate, AA, rs61757683): *ALDH4A1* encodes delta-1-pyrroline-5-carboxylate dehydrogenase which converts pyrroline-5-carboxylate to glutamate in the breakdown of proline (PMID: 2211729).

***ALDH7A1*** (6-oxopiperidine-2-carboxylate, AA, rs79449010): *ALDH7A1* encodes an aldehyde dehydrogenase which produces alpha-aminoadipic acid in the pipecolic acid pathway of lysine catabolism. Alpha-aminoadipic acid can be cyclized to form 6-oxopiperidine-2-carboxylic acid though the mechanism in human is unknown (PMID: 16491085|9544928).

***AOX1*** (7-methylguanine, NU, rs77225800): *AOX1* encodes an aldehyde oxidase which can act on many substrates including purines. 7-methylguanine is a substrate (PMID: 5044040).

***AQP10*** (arabonate/xylonate, CA, rs6702754): *AQP10* encodes an aquaglyceroporin which transports water, glycerol and larger molecules including xylitol (PMID: 21733844).

***ARSD***|***ARSL*** (ascorbic acid 3-sulfate, CV, rs1637781): *ARSD*|*ARSL* encode a pair of aryl sulfatases. The homologs *ARSA* and *ARSB* act on ascorbic acid 2-sulfate. Ascorbic acid 3-sulfate is a plausible substrate for one or both proximal enzymes (PMID: 28257906|33715).

***CDO1*** (hypotaurine, AA, rs10038137): *CDO1* encodes cysteine dioxygenase which catalyzes the first step in the conversion of cysteine to hypotaurine (PMID: 2307).

***CERS1*** (glycosyl-N-stearoyl-sphingosine (d18:1/18:0), LI, rs1013893365): *CERS1* encodes a ceramide synthase with specificity for C18 fatty acids. Sphingomyelins can be synthesized from ceramides (PMID: 12105227).

***CHKA*** (choline phosphate, LI, rs7940113): *CHKA* encodes choline kinase. Choline phosphate is the product of the reaction (PMID: 4373031).

***CRYL1*** (mannonate, XE, rs9552189): *CRYL1* encodes an enzyme with gulonate dehydrogenase activity. Mannonate is a potential substrate (PMID: 15809331).

***DBH*** (vanillylmandelate (VMA), AA, rs77273740): *DBH* encodes dopamine beta-hydroxylase which converts dopamine to noradrenaline, a precursor for vanillylmandelate. DBH can also produce vanillylmandelate directly from homovanillate (PMID: 14253475|15702409).

***DEGS1*** (palmitoyl dihydrosphingomyelin (d18:0/16:0), LI, rs752521494): *DEGS1* encodes sphingolipid delta 4-desaturase which introduces a double bond at the 4 position in sphingomyelin. Palmitoyl dihydrosphingomyelin should be a substrate for this enzyme (PMID: 11937514).

***DHDH*** (arabinose, CA, rs35230038): *DHDH* encodes dihydrodiol dehydrogenase which can oxidize arabinose and generate the corresponding lactone (PMID: 11306093).

***DPYD*** (uracil, NU, rs3918290): *DPYD* encodes dihydropyrimidine dehydrogenase which catalyzes the first step in the breakdown of uracil (PMID: 8083224).

***ECHDC1*** (ethylmalonate, AA, rs79919786): *ECHDC1* encodes ethylmalonyl-CoA decarboxylase which converts ethylmalonyl-CoA to butyryl-CoA (PMID: 22016388).

***ECI1*** (dodecadienoate (12:2), LI, rs55650311): *ECI1* encodes an enoyl-CoA delta isomerase which exchanges a double bond at the 3 position for one at the 2 position in CoA bound fatty acids with demonstrated activity on C12 enoyl-CoA. Dodecadienoate is likely derived from a product of this reaction (PMID: 8486162).

***FDX1*** (3beta-hydroxy-5-cholestenoate, LI, rs2724417): *FDX1* encodes ferredoxin 1 which is involved in electron transport for multiple cytochrome P450 enzymes, including CYP27A1. CYP27A1 plays a key role in bile acid biosynthesis and leads to the metabolites observed (PMID: 2340092).

***FTCD*** (formiminoglutamate, AA, rs398124234): *FTCD* encodes a glutamate formimidoyltransferase which transfers one-carbon units between glutamate and folate. Forminoglutamate can be a substrate or a product for this enzyme (PMID: 13672973). ***GNPTAB*** (aspartate, AA, rs1209353188): *GNPTAB* encodes an N-acetylglucosamine-1-phosphotransferase which produces aspartylglucosamines. The degradation of aspartylglucosamine produces aspartate (PMID: 6457829).

***GSTZ1*** (maleate, LI, rs7972): *GSTZ1* encodes a maleylacetoacetate isomerase which converts maleylacetoacetate to fumarylacetoacetate. Hydrolysis of maleylacetoacetate produces maleate (PMID: 9925947).

***HADHA***|***HADHB*** (3-hydroxylaurate, LI, rs116654420): *HADHA*|*HADHB* together encode a mitochondrial heterodimer which is involved in fatty acid beta oxidation. The alpha subunit of the heterodimer encodes the 3-hydroxyacyl-CoA dehydrogenase activity (PMID:1550553).

***HDAC6*** (N6-acetyllysine, AA, rs61735967): *HDAC6* encodes a lysine deacetylase which can act on multiple proteins. Acetyllysine is the substrate (PMID: 12606581).

***HSD17B10*** (tiglylcarnitine (C5:1-DC), AA, rs201378370): *HSD17B10* encodes 2-methyl-3-hydroxybutyryl-CoA dehydrogenase in the isoleucine degradation pathway. Tiglyl-CoA is an upstream metabolite and HSD17B10 deficiency results in accumulation of tiglylcarnitine and tigloylglycine (PMID: 7639524|11102558).

***KYNU*** (xanthurenate, AA, rs199546957): *KYNU* encodes kynureninase, a vitamin B6 dependent enzyme which hydrolyzes kynurenine. Xanthurenate is an alternate metabolite of kynurenine. In animals deprived of vitamin B6, excretion of xanthurenate is increased (PMID: 13032082).

***L2HGDH*** (2-hydroxyglutarate, LI, rs12886516): *L2HGDH* encodes the mitochondrial L-2-hydroxyglutarate dehydrogenase which generates 2-oxoglutarate from L-2-hydroxyglutarate (PMID: 8241290).

***MOCOS*** (7-methylguanine, NU, rs16967566): *MOCOS* encodes molybdenum cofactor sulfurase which sulfurates the molybdenum cofactor used by aldehyde oxidase (encoded by *AOX1*).

Aldehyde oxidase acts on many substrates including 7-methylguanine. The *AOX1* locus harbors its own associations with this same metabolite (PMID: 11302742|5044040).

***NAAA*** (N-stearoyltaurine, LI, rs112197434): *NAAA* encodes N-acylethanolamine-hydrolyzing acid amidase. N-acyl taurines such as n-stearoyltaurine represent a potential substrate class for NAAA (PMID: 15655246).

***NIT2*** (alpha-ketoglutaramate, AA, rs3830303): *NIT2* encodes an omega-amidase which converts alpha-ketoglutaramate to alpha ketoglutarate (PMID: 19595734).

***NR5A1*** (pregnenediol sulfate (C21H34O5S), LI, rs4838190): *NR5A1* encodes the steroidogenic factor-1, a transcription factor that regulates many genes involved in steroidogenesis, including *SULT2A1*; *SULT2A1* is also associated with pregnenediol sulfate (PMID: 22427816).

***OAT*** (3-amino-2-piperidone, AA, rs121965043): *OAT* encodes ornithine aminotransferase which converts ornithine to glutamate 5-semialdehyde. 3-amino-2-piperidone, known as cyclo-ornithine, is a closely related metabolite (PMID: 4990629).

***PCMT1*** (S-adenosylhomocysteine, AA, rs9505982): *PCMT1* encodes a methyltransferase which produces S-adenosylhomocysteine. PCMT1 is important for reversing oxidative damage in neurons and erythrocytes. Pcmt1 knockout mice show reduced levels of S-adenosylhomocysteine (PMID: 12023972|11279164).

***PTRH1*** (N-formylmethionine, AA, rs504434): *PTRH1* encodes a mitochondrial-directed peptidyl-tRNA hydrolase which releases tRNA from peptidyl-tRNA chains. N-formylmethionine is likely a product of the activity of this enzyme (PMID: 4981785).

***PYCR3*** (S-1-pyrroline-5-carboxylate, AA, rs2242090): *PYCR3* encodes a pyrroline-5-carboxylate reductase which generates proline from S-1-pyrroline-5-carboxylate (PMID: 23024808).

***QPCT*** (pyroglutamylglutamine, PE, rs77684493): *QPCT* encodes a glutaminyl-peptide cyclotransferase which cyclizes an N-terminal glutamine residue. Pyroglutamylglutamine is a likely product of this reaction (PMID: 3473473).

***SCD5*** (lignoceroylcarnitine (C24), LI, rs141560958): *SCD5* encodes stearoyl-CoA desaturase which catalyzes the formation of monounsaturated fatty acids from saturated fatty acids. The metabolite observed here, the acylcarnitine of a long chain saturated fatty acid, is a potential substrate (PMID: 15907797).

***SEC14L2*** (gamma-tocopherol/beta-tocopherol, CV, rs182488695): *SEC14L2* encodes a tocopherol-binding protein (PMID: 10829015).

***SLC10A2*** (glycodeoxycholate 3-sulfate, LI, rs55971546): *SLC10A2* encodes a bile acid transporter. Glycodeoxycholate 3-sulfate is a sulfated bile acid derivative (PMID: 7860756). ***SLC23A1*** (ascorbic acid 3-sulfate*, CV, rs33972313): *SLC23A1* encodes an ascorbate transporter. Ascorbic acid 3-sulfate and its derivatives are key associations at this locus (PMID: 10471399).

***SLC23A2*** (ascorbic acid 3-sulfate, CV, rs141583725): *SLC23A2* encodes an ascorbic acid transporter. The observed metabolite is a sulfate derivative of ascorbic acid (PMID: 10556521). ***SLC23A3*** (glycerate, CA, rs192756070): *SLC23A3* encodes an SLC23 ascorbic acid transporter without detectable ascorbic acid transport activity. *SLC23A3* is highly expressed in the kidney, and is likely as a reuptake transporter. Glycerate is a possible substrate (PMID: 23506882).

***SLC25A26*** (2,3-dihydroxy-5-methylthio-4-pentenoate (DMTPA), AA, rs13874): *SLC25A26* encodes a transporter which transports S-adenosylmethionine (SAM) into the mitochondria. 2,3-dihydroxy-5-methylthio-4-pentenoate is derived from SAM (PMID: 14674884|29578721).

***SLC27A5*** (3b-hydroxy-5-cholenoic acid, LI, rs147464959): *SLC27A5* encodes a bile acid-CoA ligase which transfers CoA to various bile acids including cholic acid. 3b-hydroxy-5-cholenoic acid is structurally similar to cholic acid (PMID: 17401).

***SLC28A1*** (5,6-dihydrouridine, NU, rs55990066): *SLC28A1* encodes a pyrimidine-specific sodium-nucleoside cotransporter (hCNT1) which can transport uridine. Dihydrouridine is a frequent modification of uridine (PMID: 9124315).

***SORD*** (ribitol, CA, rs55901542): *SORD* encodes sorbitol dehydrogenase which oxidizes multiple polyols including ribitol (PMID: 3365415).

***SPHK2*** (sphingomyelin (d18:1/20:1, d18:2/20:0), LI, rs61751862): SPHK2, a sphingosine kinase, produces sphingosine-1-P(S1P), which can be incorporated into the observed metabolite. S1P also represents the initial step in sphingomyelin catabolism (PMID: 10802064).

***ST3GAL5*** (lactosyl-N-behenoyl-sphingosine (d18:1/22:0), LI, rs7603766): *ST3GAL5* encodes lactosylceramide alpha-2,3-sialyltransferase which transfers an sialic acid moiety to lactosyl-n-behenoyl-sphingosine to produce the ganglioside GM3 (PMID: 1999428).

***TH*** (dopamine 3-O-sulfate, AA, rs200180914): TH, tyrosine hydroxylase, converts tyrosine to L-DOPA, which can then be converted to dopamine 3-O-sulfate (PMID: 14216443).

***TMLHE*** (N6,N6,N6-trimethyllysine, AA, rs547447): *TMLHE* encodes an epsilon trimethyllysine hydroxylase which adds a hydroxyl group to epsilon-n-trimethyl-lysine in the first step of carnitine biosynthesis (PMID: 6772170).

***XPNPEP2*** (prolylproline, PE, rs4830164): *XPNPEP2* encodes a protease which specifically cleaves at a proline residue. Prolylproline is a plausible substrate or product (PMID: 15361070).

The putative causal gene is in bold. The associated metabolite, its biochemical class, and the index genetic variant are in parentheses after the gene symbol. The resource publication from which the supporting evidence was identified can be accessed through the PMID at the end of supportive evidence.

## References

1. Jakkula, E. et al. The genome-wide patterns of variation expose significant substructure in a founder population. Am J Hum Genet. 83, 787–94 (2008).

2. Locke, A.E. et al. Exome sequencing of Finnish isolates enhances rare-variant association power. Nature. 572, 323–8 (2019).

3. Laakso, M. et al. The Metabolic Syndrome in Men study: a resource for studies of metabolic and cardiovascular diseases. J Lipid Res. 58, 481–93 (2017).

4. Wishart, D.S. Metabolomics for investigating physiological and pathophysiological processes. Physiol Rev. 99, 1819–75 (2019).

5. Gieger, C. et al. Genetics meets metabolomics: a genome-wide association study of metabolite profiles in human serum. PLoS Genet. 4, e1000282 (2008).

6. Illig, T. et al. A genome-wide perspective of genetic variation in human metabolism. Nat Genet. 42, 137–41 (2010).

7. Suhre, K. et al. Human metabolic individuality in biomedical and pharmaceutical research. Nature. 477, 54–60 (2011).

8. Kettunen, J. et al. Genome-wide association study identifies multiple loci influencing human serum metabolite levels. Nat Genet. 44, 269–76 (2012).

9. Rhee, E.P. et al. A genome-wide association study of the human metabolome in a community-based cohort. Cell Metab. 18, 130–43 (2013).

10. Shin, S.Y. et al. An atlas of genetic influences on human blood metabolites. Nat Genet. 46, 543–50 (2014).

11. Kastenmuller, G., Raffler, J., Gieger, C. & Suhre, K. Genetics of human metabolism: an update. Hum Mol Genet. 24, r93–r101 (2015).

12. Rhee, E.P. et al. An exome array study of the plasma metabolome. Nat Commun. 7, 12360 (2016).

13. Feofanova, E.V. et al. A genome-wide association study discovers 46 loci of the human metabolome in the Hispanic community health study/study of Latinos. Am J Hum Genet. 107, 849–63 (2020).

14. Schlosser, P. et al. Genetic studies of urinary metabolites illuminate mechanisms of detoxification and excretion in humans. Nat Genet. 52, 167–76 (2020).

15. Lotta, L.A. et al. A cross-platform approach identifies genetic regulators of human metabolism and health. Nat Genet. 53, 54–64 (2021).

16. Long, T. et al. Whole-genome sequencing identifies common-to-rare variants associated with human blood metabolites. Nat Genet. 49, 568–78 (2017).

17. Yousri, N.A. et al. Whole-exome sequencing identifies common and rare variant metabolic QTLs in a Middle Eastern population. Nat Commun. 9, 333 (2018).

18. Moore, S.C. et al. Human metabolic correlates of body mass index. Metabolomics. 10, 259–69 (2014).

19. Mehler, A.H. & Knox, W.E. The conversion of tryptophan to kynurenine in liver. II. The enzymatic hydrolysis of formylkynurenine. J Biol Chem. 187, 431–8 (1950).

20. Wen, X., Lee, Y., Luca, F. & Pique-Regi, R. Efficient integrative multi-SNP association analysis via deterministic approximation of posteriors. Am J Hum Genet. 98, 1114–29 (2016).

21. Maller, J.B. et al. Bayesian refinement of association signals for 14 loci in 3 common diseases. Nat Genet. 44, 1294–301 (2012).

22. Bürzle, M. et al. The sodium-dependent ascorbic acid transporter family SLC23. Mol Aspects Med. 34, 436–54 (2013).

23. Ginguay, A., Cynober, L., Curis, E. & Nicolis, I. Ornithine aminotransferase, an important glutamate-metabolizing enzyme at the crossroads of multiple metabolic pathways. Biology (Basel). 6, 18 (2017).

24. Mitchell, G.A. et al. An initiator codon mutation in ornithine-delta-aminotransferase causing gyrate atrophy of the choroid and retina. J Clin Invest. 81, 630–3 (1988).

25. Arshinoff, S.A. et al. Amino-acid metabolism and liver ultrastructure in hyperornithinemia with gyrate atrophy of the choroid and retina. Metabolism. 28, 979–88 (1979).

26. Yang, S.Y. et al. Mental retardation linked to mutations in the HSD17B10 gene interfering with neurosteroid and isoleucine metabolism. Proc Natl Acad Sci U S A. 106, 14820–4 (2009).

27. Chatfield, K.C. et al. Mitochondrial energy failure in HSD10 disease is due to defective mtDNA transcript processing. Mitochondrion. 21, 1–10 (2015).

28. He, X.Y., Isaacs, C. & Yang, S.Y. Roles of mitochondrial 17β-hydroxysteroid dehydrogenase type 10 in Alzheimer’s disease. J Alzheimers Dis. 62, 665–73 (2018).

29. Costa, A.C., Joaquim, H.P.G., Forlenza, O.V., Gattaz, W.F. & Talib, L.L. Three plasma metabolites in elderly patients differentiate mild cognitive impairment and Alzheimer’s disease: a pilot study. Eur Arch Psychiatry Clin Neurosci. 270, 483–8 (2020).

30. Kudo, M. et al. The alpha-and beta-subunits of the human UDP-N-acetylglucosamine:lysosomal enzyme N-acetylglucosamine-1-phosphotransferase are encoded by a single cDNA. J Biol Chem. 280, 36141–9 (2005).

31. Zhang, L., Sheng, S. & Qin, C. The role of HDAC6 in Alzheimer’s disease. J Alzheimers Dis. 33, 283–95 (2013).

32. Hunsberger, H.C. et al. Divergence in the metabolome between natural aging and Alzheimer’s disease. Sci Rep. 10, 12171 (2020).

33. Kehlen, A. et al. N-terminal pyroglutamate formation in CX3CL1 is essential for its full biologic activity. Biosci Rep. 37(2017).

34. Ripke, S. et al. Genome-wide association analysis identifies 13 new risk loci for schizophrenia. Nat Genet. 45, 1150–9 (2013).

35. Jimenez-Sanchez, M. et al. siRNA screen identifies QPCT as a druggable target for Huntington’s disease. Nat Chem Biol. 11, 347–54 (2015).

36. Yasukochi, Y. et al. Identification of CDC42BPG as a novel susceptibility locus for hyperuricemia in a Japanese population. Mol Genet Genomics. 293, 371–9 (2018).

37. Wen, X., Pique-Regi, R. & Luca, F. Integrating molecular QTL data into genome-wide genetic association analysis: probabilistic assessment of enrichment and colocalization. PLoS Genet. 13, e1006646 (2017).

38. Hukku, A. et al. Probabilistic colocalization of genetic variants from complex and molecular traits: promise and limitations. Am J Hum Genet. 108, 25–35 (2021).

39. Goldschmidt, M.L. et al. Increased frequency of double and triple heterozygous gene variants in children with intrahepatic cholestasis. Hepatol Res. 46, 306–11 (2016).

40. Vujkovic, M. et al. A genome-wide association study for nonalcoholic fatty liver disease identifies novel genetic loci and trait-relevant candidate genes in the Million Veteran Program. Preprint at medRxiv https://doi.org/10.1101/2020.12.26.20248491 (2021).

41. Ward, L.D. et al. GWAS of serum ALT and AST reveals an association of SLC30A10 Thr95Ile with hypermanganesemia symptoms. Preprint at bioRxiv https://doi.org/10.1101/2020.05.19.104570 (2021).

42. Lammert, F. et al. Gallstones. Nat Rev Dis Primers. 2, 16024 (2016).

43. Kern, F., Jr. Effects of dietary cholesterol on cholesterol and bile acid homeostasis in patients with cholesterol gallstones. J Clin Invest. 93, 1186–94 (1994).

44. Käkelä, P. et al. Serum plant sterols associate with gallstone disease independent of weight loss and non-alcoholic fatty liver disease. Obes Surg. 27, 1284–91 (2017).

45. Kundaje, A. et al. Integrative analysis of 111 reference human epigenomes. Nature. 518, 317–30 (2015).

46. The GTEx Consortium atlas of genetic regulatory effects across human tissues. Science. 369, 1318–30 (2020).

47. Buch, S. et al. A genome-wide association scan identifies the hepatic cholesterol transporter ABCG8 as a susceptibility factor for human gallstone disease. Nat Genet. 39, 995–9 (2007).

48. Wang, D.Q., Lammert, F., Cohen, D.E., Paigen, B. & Carey, M.C. Cholic acid aids absorption, biliary secretion, and phase transitions of cholesterol in murine cholelithogenesis. Am J Physiol. 276, g751–60 (1999).

49. Wang, H.H., Liu, M., Portincasa, P. & Wang, D.Q. Recent advances in the critical role of the sterol efflux transporters ABCG5/G8 in health and disease. Adv Exp Med Biol. 1276, 105–36 (2020).

50. Williams, K., Segard, A. & Graf, G.A. Sitosterolemia: twenty years of discovery of the function of ABCG5ABCG8. Int J Mol Sci. 22, 2641 (2021).

51. Weinshilboum, R.M., Thoa, N.B., Johnson, D.G., Kopin, I.J. & Axelrod, J. Proportional release of norepinephrine and dopamine--hydroxylase from sympathetic nerves. Science. 174, 1349–51 (1971).

52. Matsuo, M., Tasaki, R., Kodama, H. & Hamasaki, Y. Screening for Menkes disease using the urine HVA/VMA ratio. J Inherit Metab Dis. 28, 89–93 (2005).

53. Ehret, G.B. et al. The genetics of blood pressure regulation and its target organs from association studies in 342,415 individuals. Nat Genet. 48, 1171–84 (2016).

54. Giri, A. et al. Trans-ethnic association study of blood pressure determinants in over 750,000 individuals. Nat Genet. 51, 51–62 (2019).

55. Kestilä, M., Ikonen, E. & Lehesjoki, A.E. Finnish disease heritage. Duodecim. 126, 2311–20 (2010).

56. Krawczyk, M. et al. Phytosterol and cholesterol precursor levels indicate increased cholesterol excretion and biosynthesis in gallstone disease. Hepatology. 55, 1507–17 (2012).

57. Malina, D.M. et al. Additive effects of plant sterols supplementation in addition to different lipid-lowering regimens. J Clin Lipidol. 9, 542–52 (2015).

58. Paltoo, D.N. et al. Data use under the NIH GWAS data sharing policy and future directions. Nat Genet. 46, 934–8 (2014).

59. Gagliano Taliun, S.A. et al. Exploring and visualizing large-scale genetic associations by using PheWeb. Nat Genet. 52, 550–2 (2020).

60. Wishart, D.S. et al. HMDB 4.0: the human metabolome database for 2018. Nucleic Acids Res. 46, d608–d17 (2018).

61. Evans, A.M. et al. High resolution mass spectrometry improves data quantity and quality as compared to unit mass resolution mass spectrometry in high-throughput profiling metabolomics. Metabolomics. 4, 2 (2014).

62. Teslovich, T.M. et al. Identification of seven novel loci associated with amino acid levels using single-variant and gene-based tests in 8545 Finnish men from the METSIM study. Hum Mol Genet. 27, 1664–74 (2018).

63. Ganel, L. et al. Mitochondrial genome copy number measured by DNA sequencing in human blood is strongly associated with metabolic traits via cell-type composition differences. Hum Genomics. 15, 34 (2021).

64. Li, H. A statistical framework for SNP calling, mutation discovery, association mapping and population genetical parameter estimation from sequencing data. Bioinformatics. 27, 2987–93 (2011).

65. Jun, G. et al. Detecting and estimating contamination of human DNA samples in sequencing and array-based genotype data. Am J Hum Genet. 91, 839–48 (2012).

66. Fuchsberger, C. et al. The genetic architecture of type 2 diabetes. Nature. 536, 41–7 (2016).

67. Browning, S.R. & Browning, B.L. Rapid and accurate haplotype phasing and missing-data inference for whole-genome association studies by use of localized haplotype clustering. Am J Hum Genet. 81, 1084–97 (2007).

68. Manichaikul, A. et al. Robust relationship inference in genome-wide association studies. Bioinformatics. 26, 2867–73 (2010).

69. Das, S. et al. Next-generation genotype imputation service and methods. Nat Genet. 48, 1284–7 (2016).

70. McLaren, W. et al. The Ensembl variant effect predictor. Genome Biol. 17, 122 (2016).

71. Flannick, J. et al. Exome sequencing of 20,791 cases of type 2 diabetes and 24,440 controls. Nature. 570, 71–6 (2019).

72. Liu, X., Li, C., Mou, C., Dong, Y. & Tu, Y. dbNSFP v4: a comprehensive database of transcript-specific functional predictions and annotations for human nonsynonymous and splice-site SNVs. Genome Med. 12, 103 (2020).

73. Adzhubei, I.A. et al. A method and server for predicting damaging missense mutations. Nat Methods. 7, 248–9 (2010).

74. Vaser, R., Adusumalli, S., Leng, S.N., Sikic, M. & Ng, P.C. SIFT missense predictions for genomes. Nat Protoc. 11, 1–9 (2016).

75. Schwarz, J.M., Cooper, D.N., Schuelke, M. & Seelow, D. MutationTaster2: mutation prediction for the deep-sequencing age. Nat Methods. 11, 361–2 (2014).

76. Chun, S. & Fay, J.C. Identification of deleterious mutations within three human genomes. Genome Res. 19, 1553–61 (2009).

77. Troyanskaya, O. et al. Missing value estimation methods for DNA microarrays. Bioinformatics. 17, 520–5 (2001).

78. Buniello, A. et al. The NHGRI-EBI GWAS Catalog of published genome-wide association studies, targeted arrays and summary statistics 2019. Nucleic Acids Res. 47, d1005–d12 (2019).

79. Maglott, D., Ostell, J., Pruitt, K.D. & Tatusova, T. Entrez gene: gene-centered information at NCBI. Nucleic Acids Res. 39, d52–7 (2011).

80. Ashburner, M. et al. Gene ontology: tool for the unification of biology. The Gene Ontology Consortium. Nat Genet. 25, 25–9 (2000).

81. Ogata, H. et al. KEGG: Kyoto encyclopedia of genes and genomes. Nucleic Acids Res. 27, 29–34 (1999).

82. Benner, C. et al. FINEMAP: efficient variable selection using summary data from genome-wide association studies. Bioinformatics. 32, 1493–501 (2016).

83. Wen, X. Molecular QTL discovery incorporating genomic annotations using Bayesian false discovery rate control. Ann Appl Stat. 10, 1619–38 (2016).

84. Burgess, S., Butterworth, A. & Thompson, S.G. Mendelian randomization analysis with multiple genetic variants using summarized data. Genet Epidemiol. 37, 658–65 (2013).

85. Bowden, J., Davey Smith, G., Haycock, P.C. & Burgess, S. Consistent estimation in Mendelian randomization with some invalid instruments using a weighted median estimator. Genet Epidemiol. 40, 304–14 (2016).

86. Verbanck, M., Chen, C.Y., Neale, B. & Do, R. Detection of widespread horizontal pleiotropy in causal relationships inferred from Mendelian randomization between complex traits and diseases. Nat Genet. 50, 693–8 (2018).

87. Bowden, J., Davey Smith, G. & Burgess, S. Mendelian randomization with invalid instruments: effect estimation and bias detection through Egger regression. Int J Epidemiol. 44, 512–25 (2015).

