## Supplementary File for "Genome-wide association study of 1,391 plasma metabolites in 6,136 Finnish men identifies 303 novel signals and provides biological insights into human diseases"

### Supplementary Material

#### Supplementary Figures

Supplementary Figure 1 ------------------------------------------------------------------ 2

Supplementary Figure 2 ------------------------------------------------------------------ 3

Supplementary Figure 3 ------------------------------------------------------------------ 4

Supplementary Figure 4 ------------------------------------------------------------------ 5

Supplementary Figure 5 ------------------------------------------------------------------ 6

Supplementary Figure 6 ------------------------------------------------------------------ 7

Supplementary Figure 7 ------------------------------------------------------------------ 8

Supplementary Figure 8 ------------------------------------------------------------------ 9

Supplementary Figure 9 ------------------------------------------------------------------ 10

Supplementary Figure 10 ------------------------------------------------------------------ 11

Supplementary Figure 11 ------------------------------------------------------------------ 12

Supplementary Figure 12 ------------------------------------------------------------------ 13

Supplementary Figure 13 ------------------------------------------------------------------ 14

#### Supplementary Note

FinnGen Contributors ------------------------------------------------------------------ 15

Bibliography for Supplementary Table 11 ------------------------------------------------------------------ 27

### Supplementary Figures

**Supplementary Figure 1**: Flow chart of the METSIM metabolomics study. MAC: minor allele count; SPIP and VPIP: signal and variant posterior inclusion probability in DAP-g Bayesian fine mapping; RCP: regional colocalization posterior probability in FastENLOC; PTV: protein-truncating variant; VMA: vanillylmandelate.

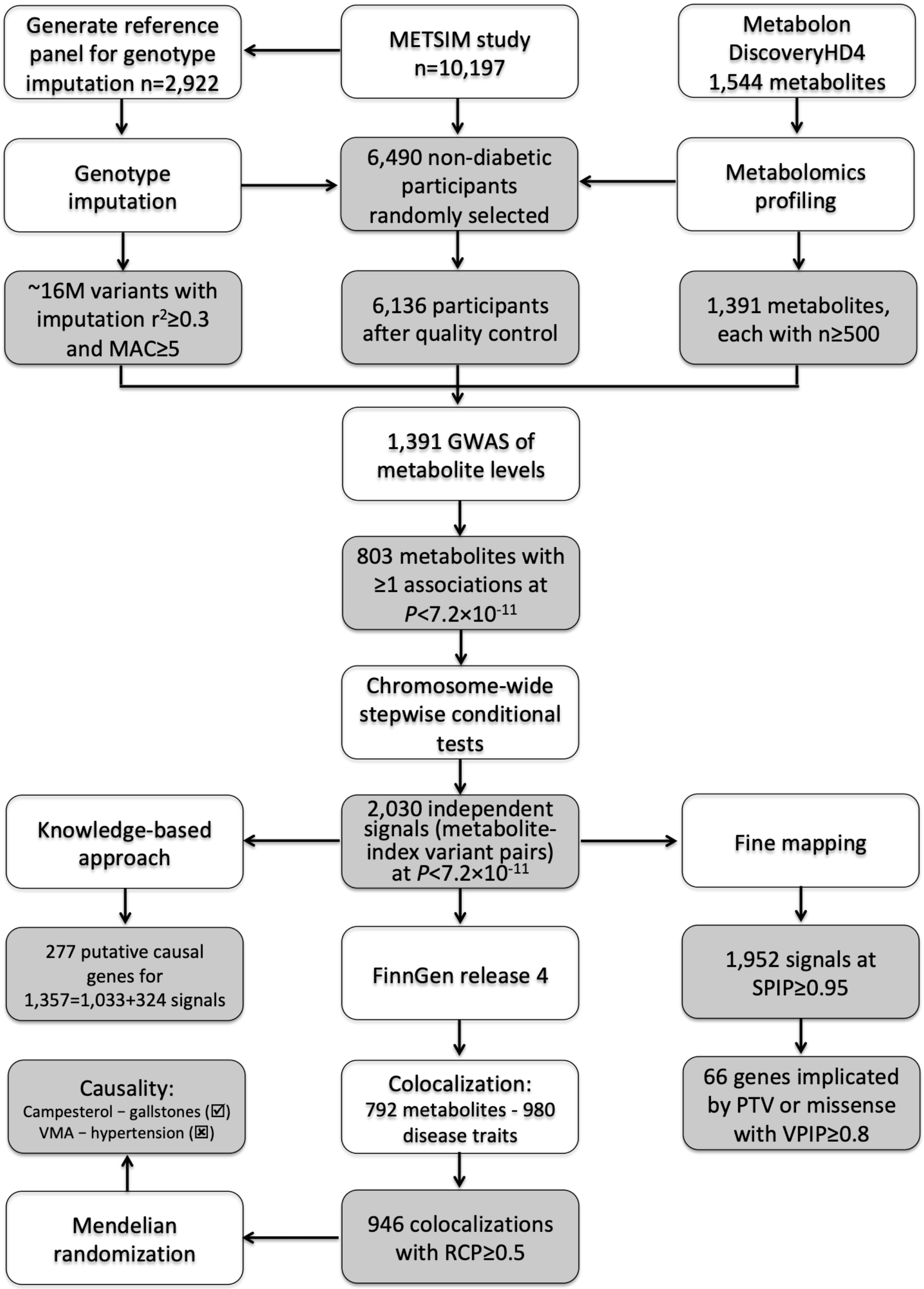

**Supplementary Figure 2**: Distribution of genomic control inflation factors for the 1,391 metabolite GWAS (median=1.00).

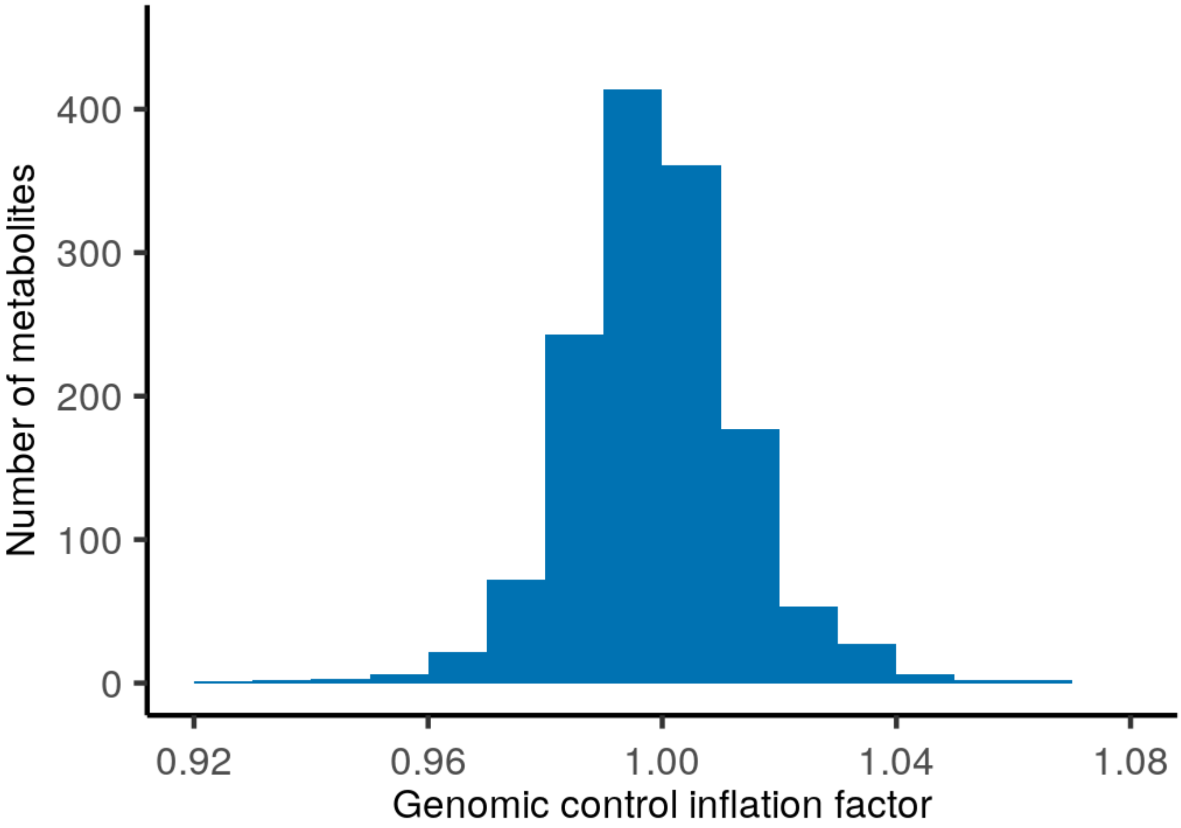

**Supplementary Figure 3**: Comparison of significant single-variant association results ($P<7.2\times10^{-11}$) with or without adjustment for BMI: a) $-\log_{10}P$ and b) effect size estimate. For each comparison, the Pearson correlation coefficient r=0.999. Each dot represents a genetic variant-metabolite pair.

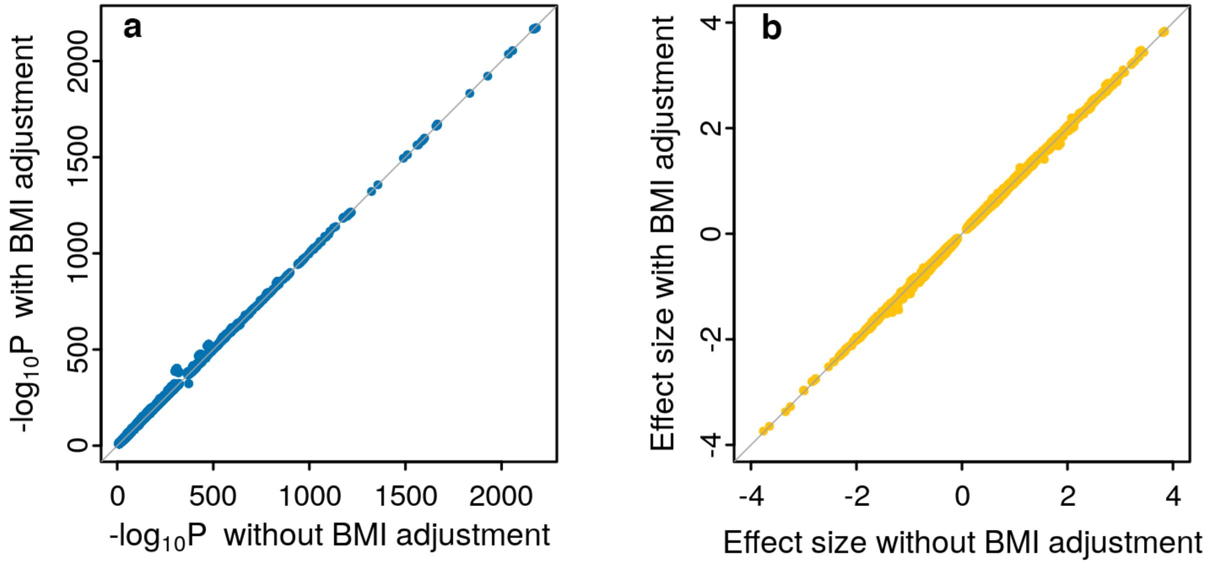

**Supplementary Figure 4**: Absolute value of the effect size estimate is inversely correlated with minor allele frequency (MAF) for the 2,030 conditional association signals (1,143 index variants; $P<7.2\times10^{-11}$). Each dot represents an association signal. Novel signals are colored in blue. Effect size was estimated for inverse normalized metabolite level residuals after regression on covariates. Four novel association signals at rare variants and one at a common variant are labeled.

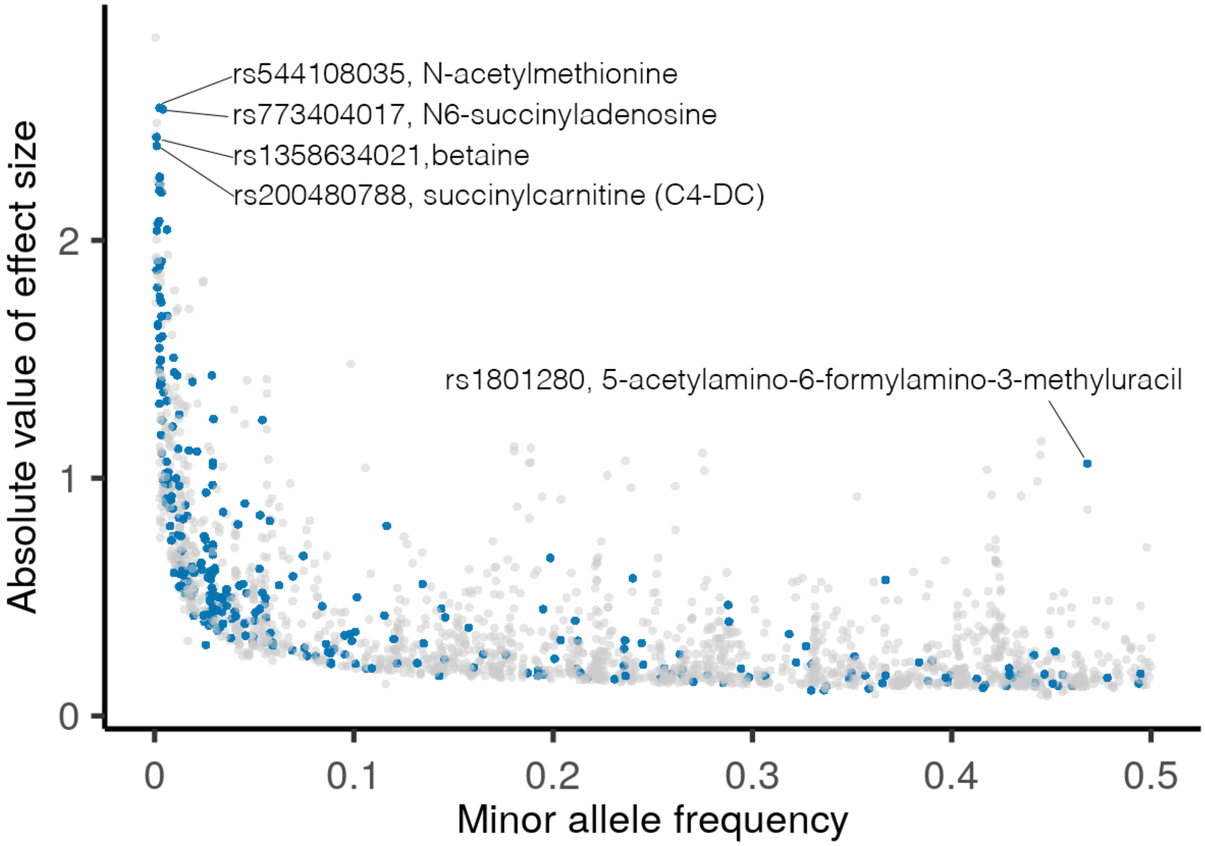

**Supplementary Figure 5**: Relationship between the number of metabolites and the number of significant ($P$<7.2x10$^{-11}$) conditional association signals for the ten metabolite biochemical classes. The proportion of metabolites and association signals are plotted on the log scale.

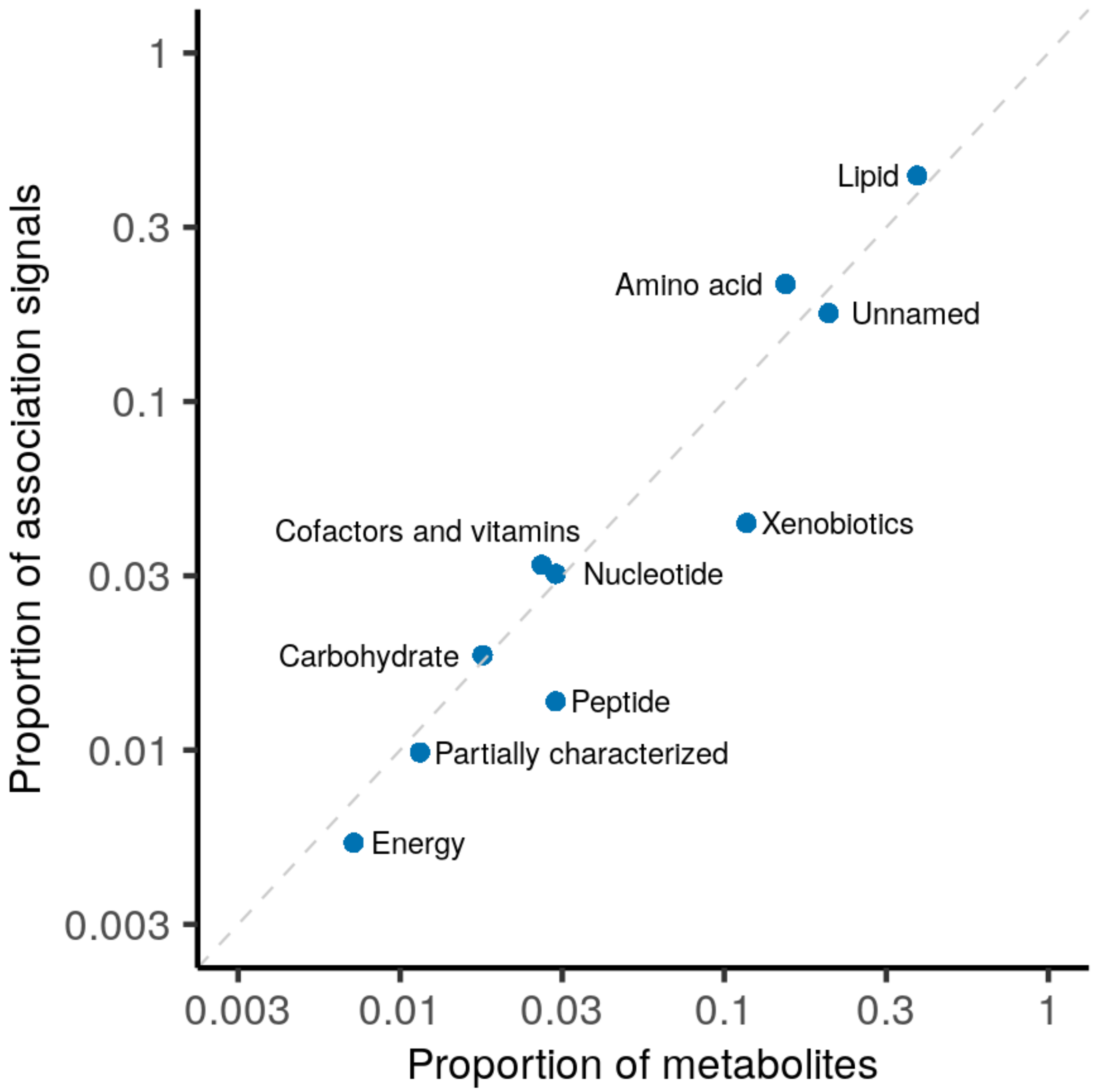

**Supplementary Figure 6**: Number of metabolites ($P<7.2x10^{-11}$) associated with each of the 1,143 index variants.

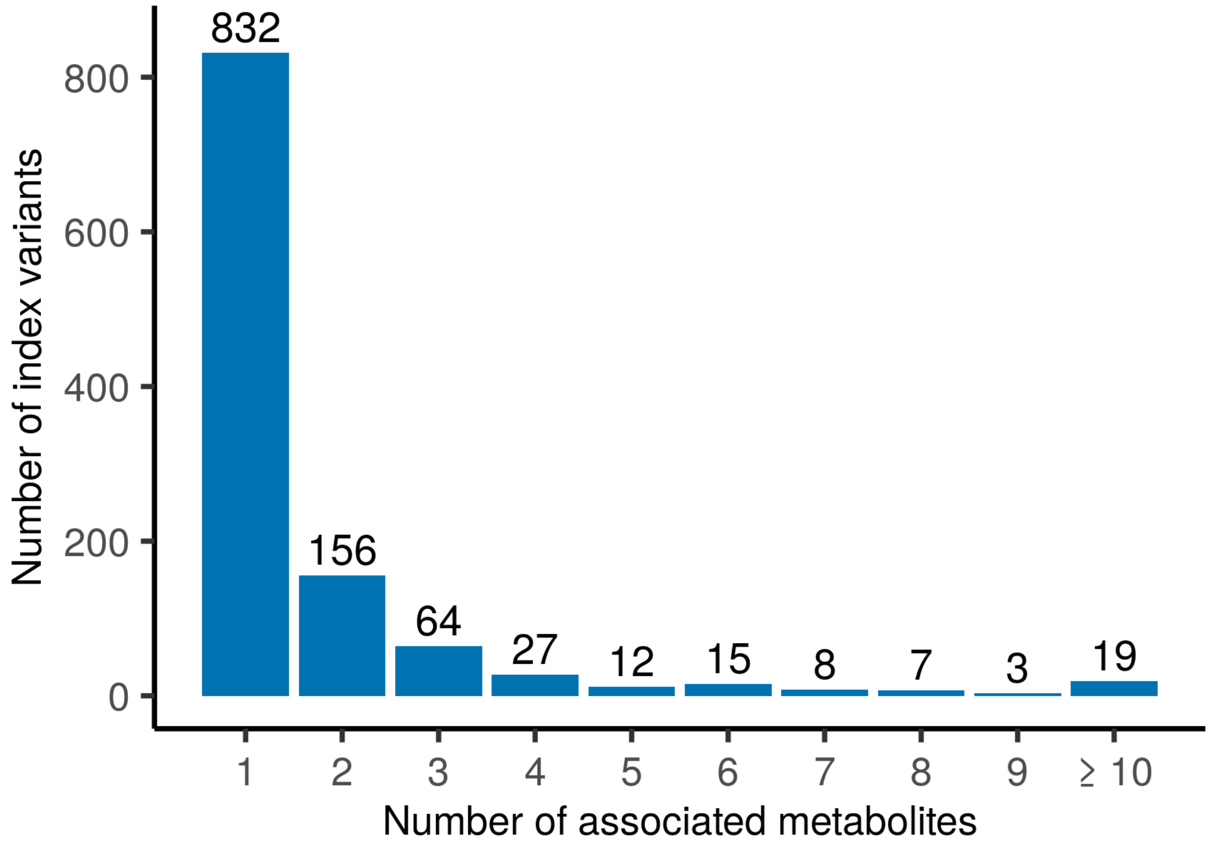

**Supplementary Figure 7**: Distribution of metabolite phenotypic variance explained by the index variant for each of the 2,030 significant association signals. The distribution is plotted on the log scale.

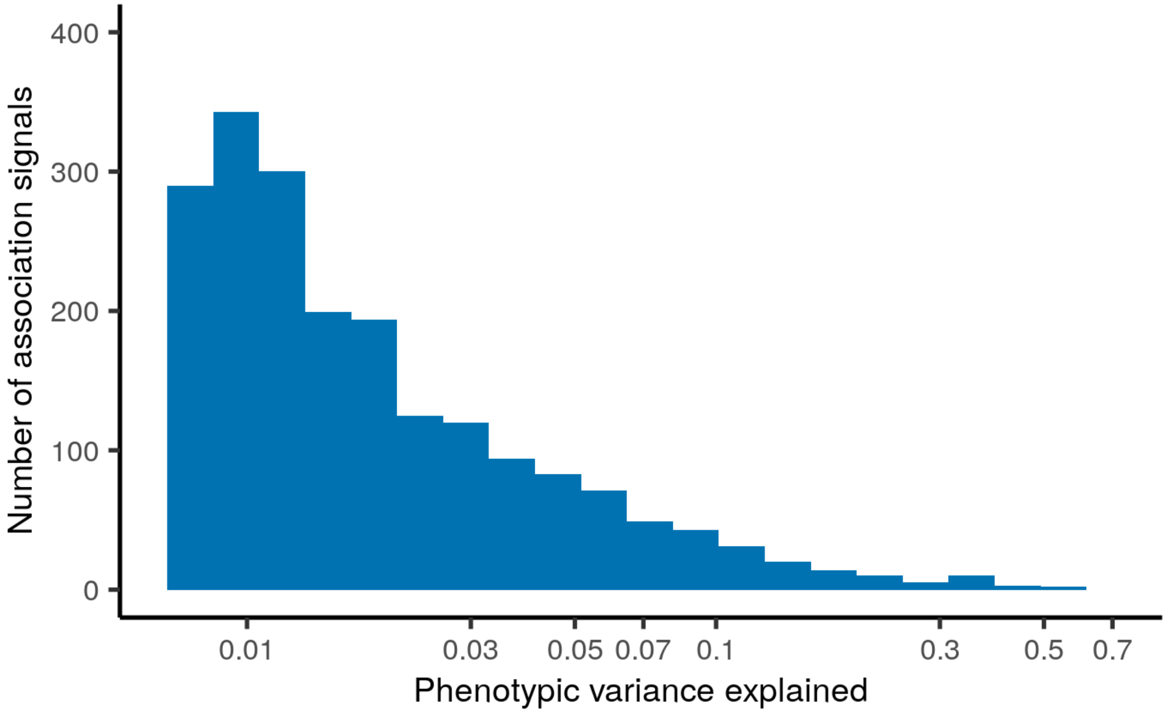

**Supplementary Figure 8**: Significant genetic associations ($P<7.2\times10^{-11}$) between *SLC23A3* missense variant p.Asn336Lys (rs192756070) and 19 metabolites. a) Phenotypic correlations between the 19 metabolites; and b) Effect size estimates and standard errors for the 19 metabolite association signals sorted by biochemical class. Within each class, metabolites are sorted by their association effect sizes.

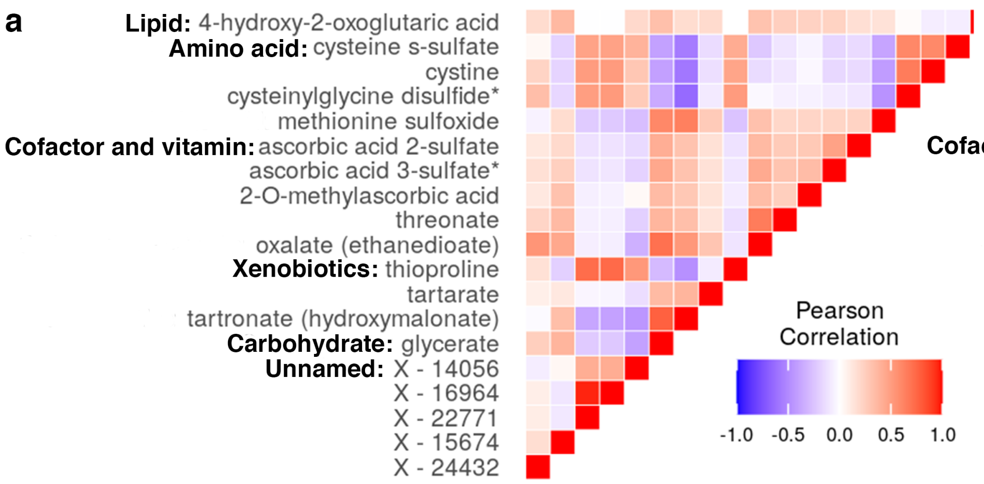


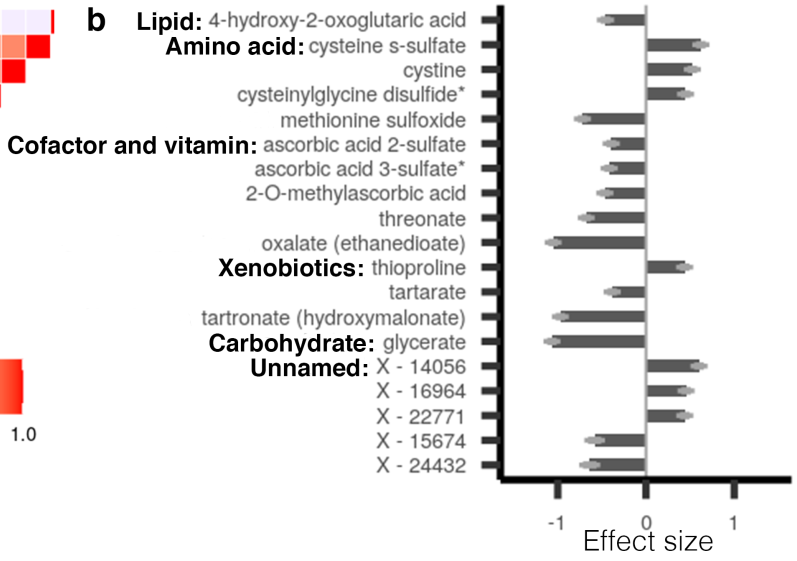

**Supplementary Figure 9:** Number of conditional association signals ($P<7.2x10^{-11}$) associated with each of the 215 putative causal genes.

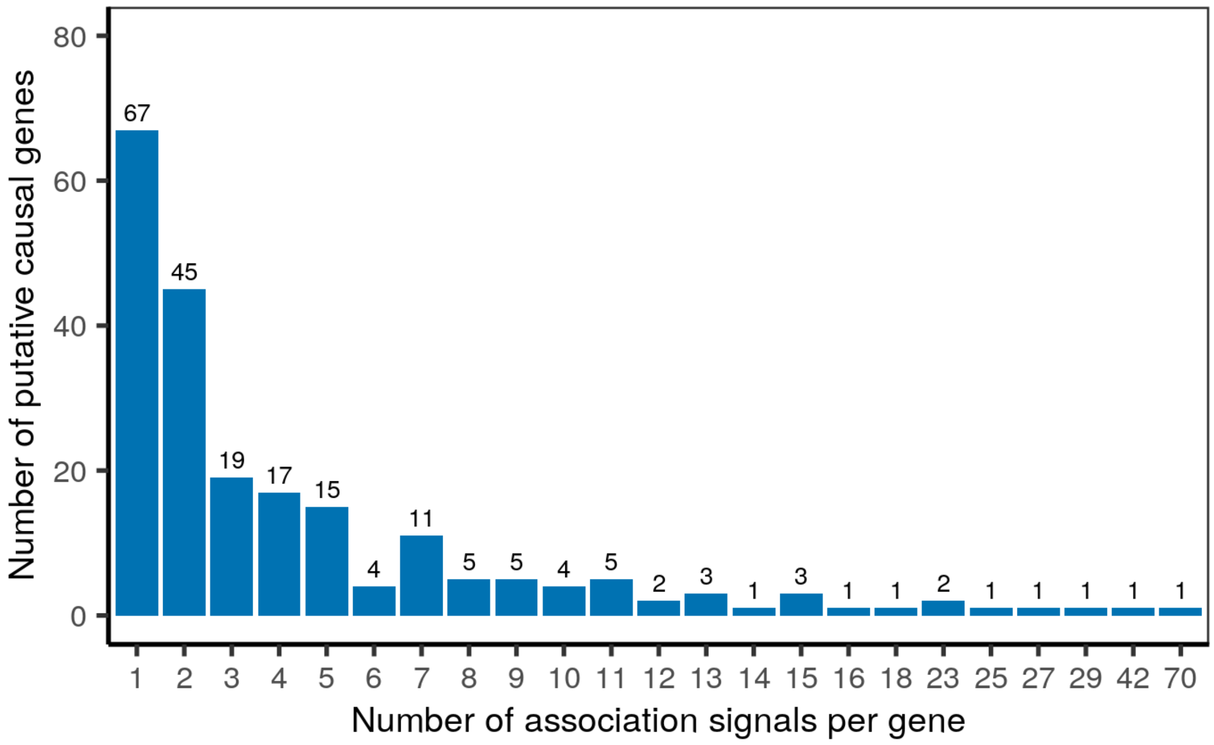

**Supplementary Figure 10**: Integrating metabolite GWAS into FinnGen disease genetic association results through colocalization analysis increases the posterior probabilities of a) 64% of the disease association signals (SPIP); and b) 90% of the putative causal variants (VPIP). SPIP is the estimated probability that there is an association signal in the region. Here, VPIP is the largest posterior probability that one of the genetic variants is causal. Each point denotes in a) an association signal and in b) a genetic variant.

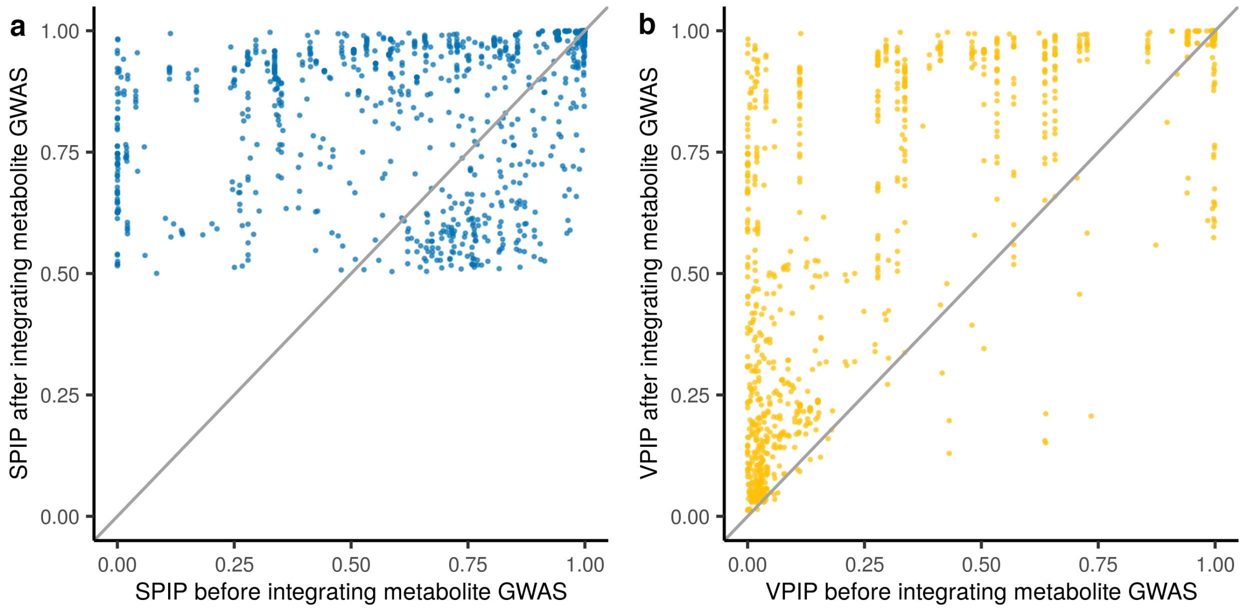

**Supplementary Figure 11**: Potential causal pathway from plasma campesterol to the risk of gallstones at the *ABCG5/ABCG8* region and the potential distinct effects of the *DBH* gene on plasma vanillylmandelate (VMA) and the risk of hypertension.

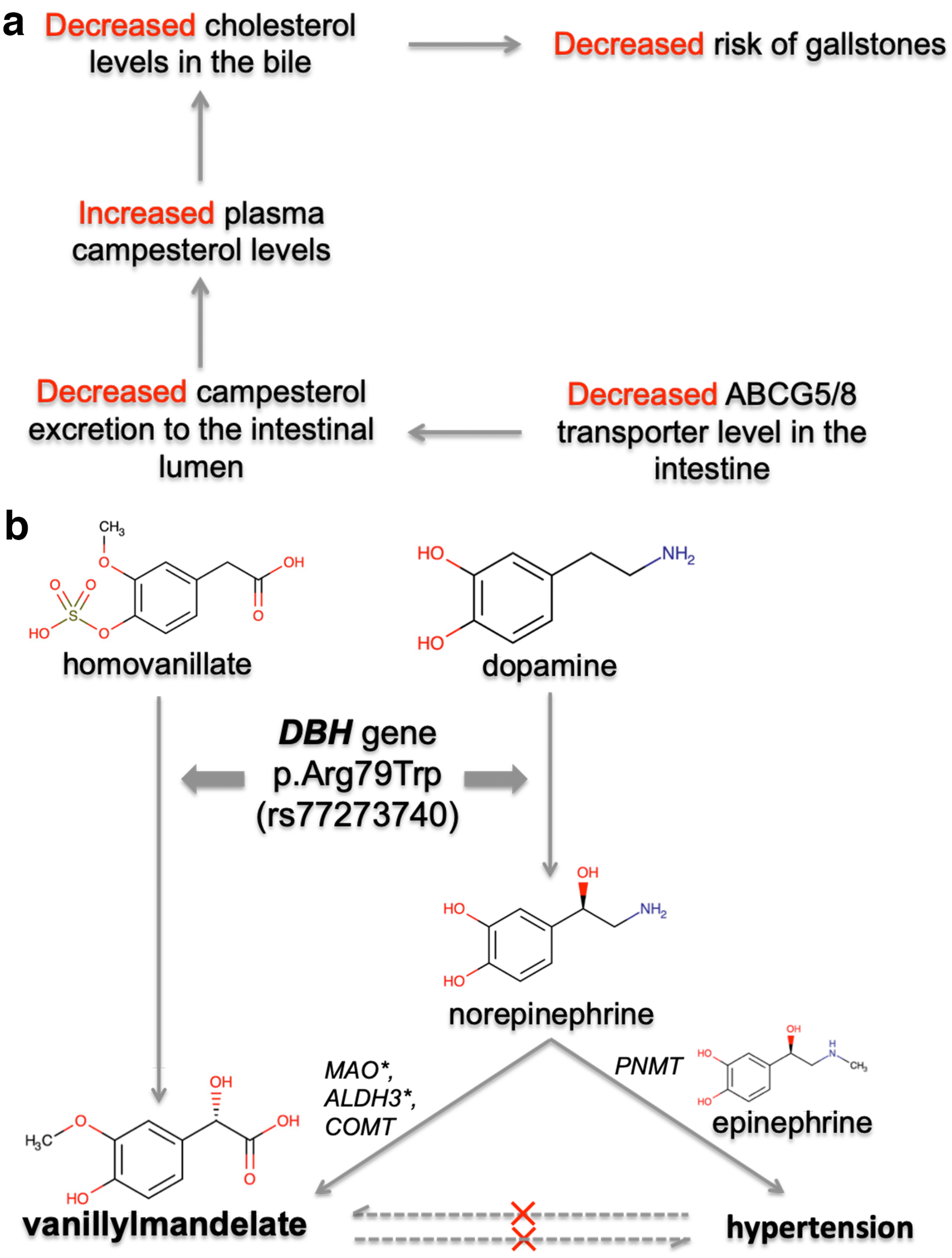

**Supplementary Figure 12**: Dopamine beta hydroxylase (*DBH*) influence on vanillylmandelate and hypertension: distinct pathways. a) Stacked regional association plots for vanillylmandelate and hypertension in the *DBH* region of chromosome 9. Genetic variants are colored by their linkage disequilibrium (LD) in METSIM to the index variant rs77273740; b) Comparison of effect sizes of ten instrumental variables without significant heterogeneity (*P*>0.05) in the genome used in the Mendelian randomization analysis between vanillylmandelate and hypertension. The slope of the blue dashed line depicts the estimated effect of vanillylmandelate on hypertension in Mendelian randomization analysis; and c) Effects on hypertension of ten instrumental variables used in Mendelian randomization analysis. OR: odds ratio.

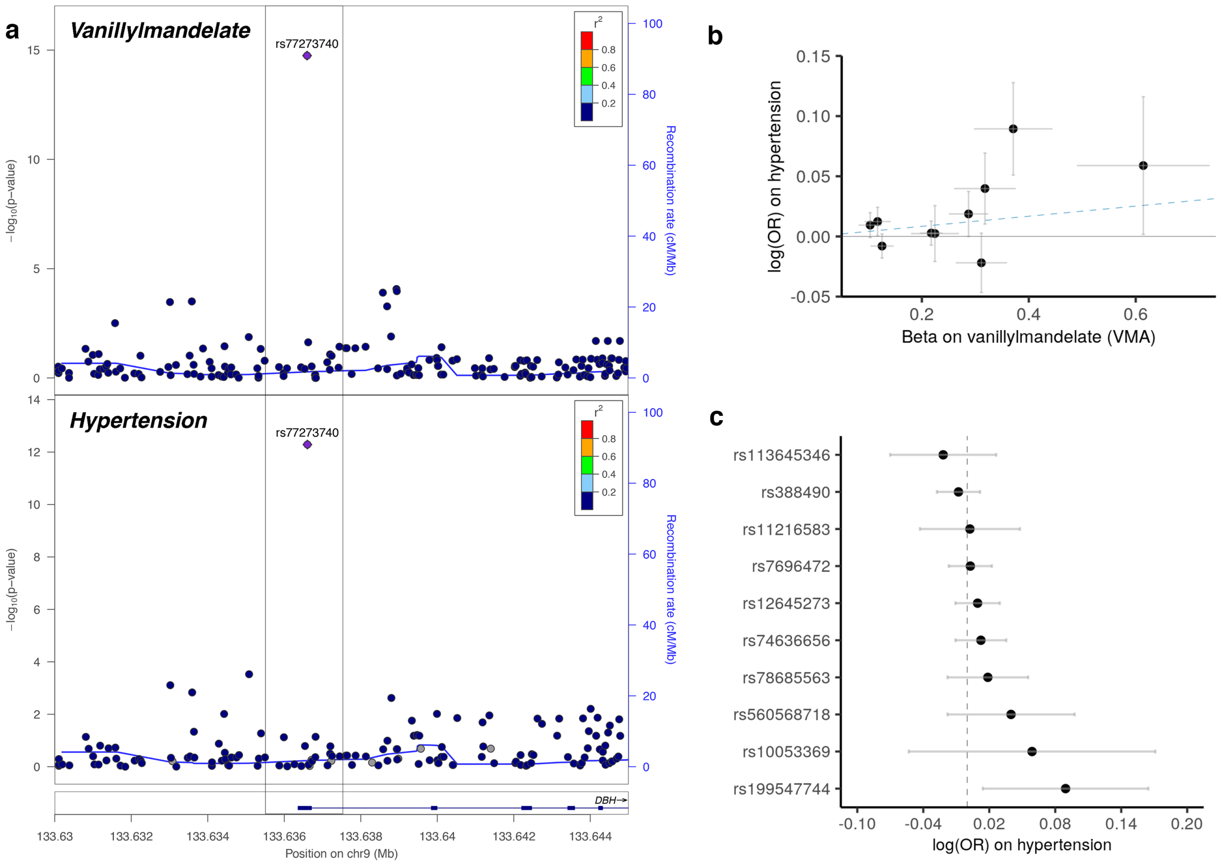

**Supplementary Figure 13**: Proportion of a) METSIM participants with missing data for each of the 1,391 metabolites; and b) metabolites with missing data among the 6,136 METSIM participants in the analysis dataset.

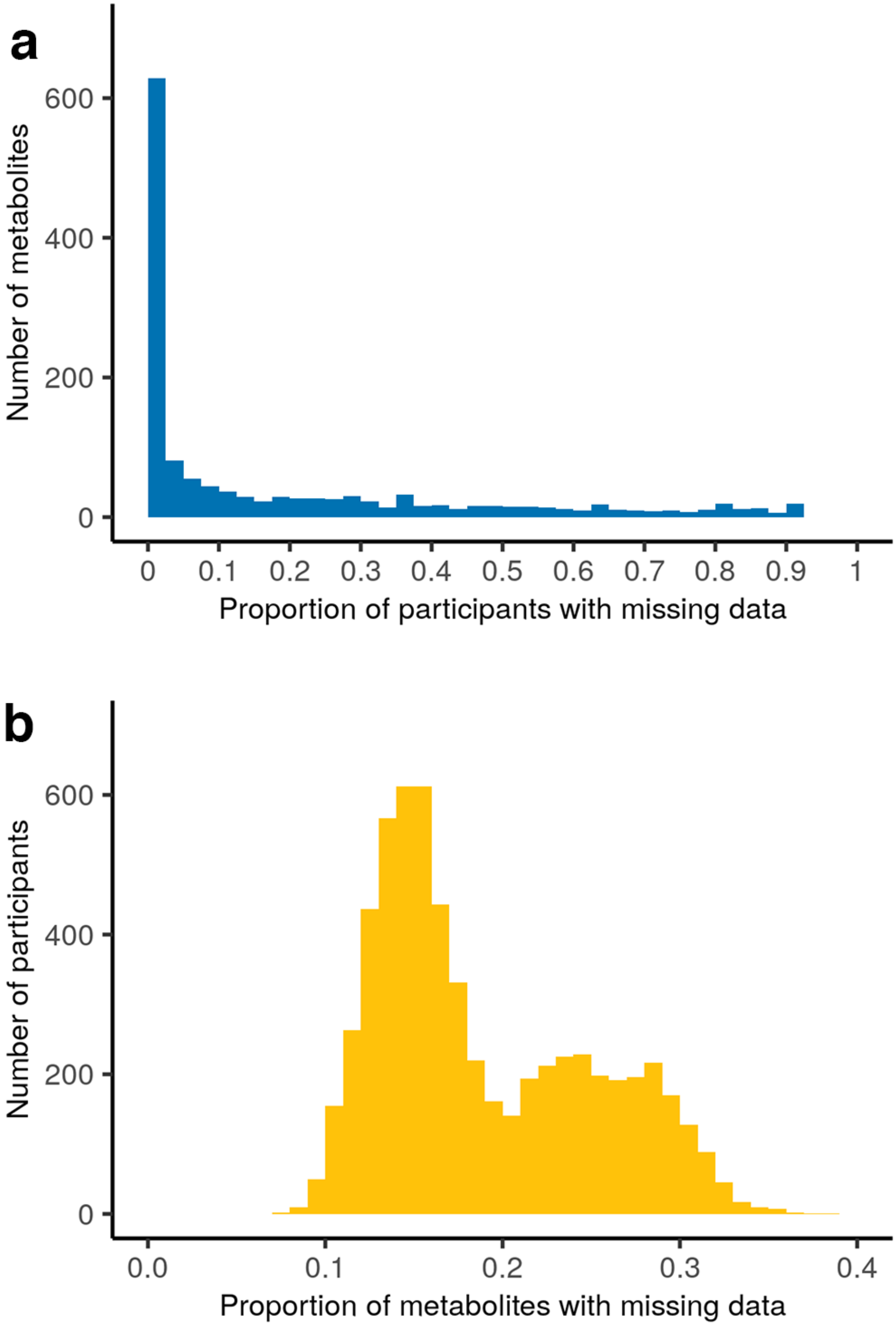

### Supplementary Note

#### FinnGen Contributors

##### Steering Committee

Aarno Palotie	Institute for Molecular Medicine Finland, HiLIFE, University of Helsinki, Finland
Mark Daly	Institute for Molecular Medicine Finland, HiLIFE, University of Helsinki, Finland

##### Pharmaceutical companies

Bridget Riley-Gills	Abbvie, Chicago, IL, United States
Howard Jacob	Abbvie, Chicago, IL, United States
Dirk Paul	Astra Zeneca, Cambridge, United Kingdom
Heiko Runz	Biogen, Cambridge, MA, United States
Sally John	Biogen, Cambridge, MA, United States
Robert Plenge	Celgene, Summit, NJ, United States/Bristol Myers Squibb, New York, NY, United States
Joseph Maranville	Celgene, Summit, NJ, United States/Bristol Myers Squibb, New York, NY, United States
Mark McCarthy	Genentech, San Francisco, CA, United States
Julie Hunkapiller	Genentech, San Francisco, CA, United States
Meg Ehm	GlaxoSmithKline, Brentford, United Kingdom
Kirsi Auro	GlaxoSmithKline, Brentford, United Kingdom
Simonne Longerich	Merck, Kenilworth, NJ, United States
Caroline Fox	Merck, Kenilworth, NJ, United States
Anders Mälarstig	Pfizer, New York, NY, United States
Katherine Klinger	Sanofi, Paris, France
Deepak Raipal	Sanofi, Paris, France
Eric Green	Maze Therapeutics, San Francisco, CA, United States
Robert Graham	Maze Therapeutics, San Francisco, CA, United States
Robert Yang	Janssen Biotech, Beerse, Belgium
Richard Siegel	Novartis, Basel, Switzerland

##### University of Helsinki & Biobanks

Tomi Mäkelä	HiLIFE, University of Helsinki, Finland, Finland
Jaakko Kaprio	Institute for Molecular Medicine Finland, HiLIFE, Helsinki, Finland, Finland
Petri Virolainen	Auria Biobank/University of Turku/Hospital District of Southwest Finland, Turku, Finland
Antti Hakanen	Auria Biobank/University of Turku/Hospital District of Southwest Finland, Turku, Finland
Terhi Kilpi	THL Biobank / The National Institute of Health and Welfare Helsinki, Finland
Markus Perola	THL Biobank / The National Institute of Health and Welfare Helsinki, Finland
Jukka Partanen	Finnish Red Cross Blood Service / Finnish Hematology Registry and Clinical Biobank, Helsinki, Finland
Anne Pitkäranta	Helsinki Biobank / Helsinki University and Hospital District of Helsinki and Uusimaa, Helsinki
Juhani Junttila	Northern Finland Biobank Borealis / University of Oulu / Northern Ostrobothnia Hospital District, Oulu, Finland
Raisa Serpi	Northern Finland Biobank Borealis / University of Oulu / Northern Ostrobothnia Hospital District, Oulu, Finland
Tarja Laitinen	Finnish Clinical Biobank Tampere / University of Tampere / Pirkanmaa Hospital District, Tampere, Finland
Veli-Matti Kosma	Biobank of Eastern Finland / University of Eastern Finland / Northern Savo Hospital District, Kuopio, Finland

Urho Kujala Central Finland Biobank / University of Jyväskylä / Central Finland Health Care District, Jyväskylä, Finland
Marco Hautalahti FINBB - Finnish biobank cooperative

**Other Experts/ Non-Voting Members**
Outi Tuovila Business Finland, Helsinki, Finland
Raimo Pakkanen Business Finland, Helsinki, Finland

#### Scientific Committee

**Pharmaceutical companies**

| | |
|---|---|
| Jeffrey Waring | Abbvie, Chicago, IL, United States |
| Bridget Riley-Gillis | Abbvie, Chicago, IL, United States |
| Ioanna Tachmazidou | Astra Zeneca, Cambridge, United Kingdom |
| Chia-Yen Chen | Biogen, Cambridge, MA, United States |
| Heiko Runz | Biogen, Cambridge, MA, United States |
| Shameek Biswas | Celgene, Summit, NJ, United States/Bristol Myers Squibb, New York, NY, United States |
| Sarah Pendergrass | Genentech, San Francisco, CA, United States |
| Julie Hunkapiller | Genentech, San Francisco, CA, United States |
| Meg Ehm | GlaxoSmithKline, Brentford, United Kingdom |
| David Pulford | GlaxoSmithKline, Brentford, United Kingdom |
| Neha Raghavan | Merck, Kenilworth, NJ, United States |
| Adriana Huertas-Vazquez | Merck, Kenilworth, NJ, United States |
| Anders Mälarstig | Pfizer, New York, NY, United States |
| Xinli Hu | Pfizer, New York, NY, United States |
| Katherine Klinger | Sanofi, Paris, France |
| Matthias Gossel | Sanofi, Paris, France |
| Robert Graham | Maze Therapeutics, San Francisco, CA, United States |
| Eric Green | Maze Therapeutics, San Francisco, CA, United States |
| Sahar Mozaffari | Maze Therapeutics, San Francisco, CA, United States |
| Dawn Waterworth | Janssen Biotech, Beerse, Belgium |
| Nicole Renaud | Novartis, Basel, Switzerland |
| Ma´en Obeidat | Novartis, Basel, Switzerland |

**University of Helsinki & Biobanks**

| | |
|---|---|
| Samuli Ripatti | Institute for Molecular Medicine Finland, HiLIFE, Helsinki, Finland |
| Johanna Schleutker | Auria Biobank / Univ. of Turku / Hospital District of Southwest Finland, Turku, Finland |
| Markus Perola | THL Biobank / The National Institute of Health and Welfare Helsinki, Finland |
| Mikko Arvas | Finnish Red Cross Blood Service / Finnish Hematology Registry and Clinical Biobank, Helsinki, Finland |
| Olli Carpén | Helsinki Biobank / Helsinki University and Hospital District of Helsinki and Uusimaa, Helsinki |
| Reetta Hinttala | Northern Finland Biobank Borealis / University of Oulu / Northern Ostrobothnia Hospital District, Oulu, Finland |
| Johannes Kettunen | Northern Finland Biobank Borealis / University of Oulu / Northern Ostrobothnia Hospital District, Oulu, Finland |
| Arto Mannermaa | Biobank of Eastern Finland / University of Eastern Finland / Northern Savo Hospital District, Kuopio, Finland |
| TBC | Finnish Clinical Biobank Tampere / University of Tampere / Pirkanmaa Hospital District, Tampere, Finland |
| Jari Laukkanen | Central Finland Biobank / University of Jyväskylä / Central Finland Health Care District, Jyväskylä, Finland |
| Urho Kujala | Central Finland Biobank / University of Jyväskylä / Central Finland Health Care District, Jyväskylä, Finland |

| | |
|---|---|
| Johanna Mäkelä | FINBB - Finnish biobank cooperative |

#### Clinical Groups

##### Neurology Group

| | |
|---|---|
| Reetta Kälviäinen | Northern Savo Hospital District, Kuopio, Finland |
| Valtteri Julkunen | Northern Savo Hospital District, Kuopio, Finland |
| Hilkka Soininen | Northern Savo Hospital District, Kuopio, Finland |
| Anne Remes | Northern Ostrobothnia Hospital District, Oulu, Finland |
| Mikko Hiltunen | Northern Savo Hospital District, Kuopio, Finland |
| Jukka Peltola | Pirkanmaa Hospital District, Tampere, Finland |
| Pentti Tienari | Hospital District of Helsinki and Uusimaa, Helsinki, Finland |
| Juha Rinne | Hospital District of Southwest Finland, Turku, Finland |
| Roosa Kallionpää | Hospital District of Southwest Finland, Turku, Finland |
| Ali Abbasi | Abbvie, Chicago, IL, United States |
| Adam Ziemann | Abbvie, Chicago, IL, United States |
| Jeffrey Waring | Abbvie, Chicago, IL, United States |
| Sahar Esmaeeli | Abbvie, Chicago, IL, United States |
| Nizar Smaoui | Abbvie, Chicago, IL, United States |
| Anne Lehtonen | Abbvie, Chicago, IL, United States |
| Susan Eaton | Biogen, Cambridge, MA, United States |
| Heiko Runz | Biogen, Cambridge, MA, United States |
| Sanni Lahdenperä | Biogen, Cambridge, MA, United States |
| Janet van Adelsberg | Celgene, Summit, NJ, United States/ Bristol Myers Squibb, New York, NY, United States |
| Shameek Biswas | Celgene, Summit, NJ, United States/ Bristol Myers Squibb, New York, NY, United States |
| Julie Hunkapiller | Genentech, San Francisco, CA, United States |
| Natalie Bowers | Genentech, San Francisco, CA, United States |
| Edmond Teng | Genentech, San Francisco, CA, United States |
| Sarah Pendergrass | Genentech, San Francisco, CA, United States |
| Onuralp Soylemez | Merck, Kenilworth, NJ, United States |
| Kari Linden | Pfizer, New York, NY, United States |
| Fanli Xu | GlaxoSmithKline, Brentford, United Kingdom |
| David Pulford | GlaxoSmithKline, Brentford, United Kingdom |
| Kirsi Auro | GlaxoSmithKline, Brentford, United Kingdom |
| Laura Addis | GlaxoSmithKline, Brentford, United Kingdom |
| John Eicher | GlaxoSmithKline, Brentford, United Kingdom |
| Minna Raivio | Hospital District of Helsinki and Uusimaa, Helsinki, Finland |
| Sarah Pendergrass | Genentech, San Francisco, CA, United States |
| Beryl Cummings | Maze Therapeutics, San Francisco, CA, United States |
| Juulia Partanen | Institute for Molecular Medicine Finland, HiLIFE, University of Helsinki, Finland |

##### Gastroenterology Group

| | |
|---|---|
| Martti Färkkilä | Hospital District of Helsinki and Uusimaa, Helsinki, Finland |
| Jukka Koskela | Hospital District of Helsinki and Uusimaa, Helsinki, Finland |
| Sampsa Pikkarainen | Hospital District of Helsinki and Uusimaa, Helsinki, Finland |
| Airi Jussila | Pirkanmaa Hospital District, Tampere, Finland |
| Katri Kaukinen | Pirkanmaa Hospital District, Tampere, Finland |
| Timo Blomster | Northern Ostrobothnia Hospital District, Oulu, Finland |
| Mikko Kiviniemi | Northern Savo Hospital District, Kuopio, Finland |
| Markku Voutilainen | Hospital District of Southwest Finland, Turku, Finland |
| Ali Abbasi | Abbvie, Chicago, IL, United States |
| Graham Heap | Abbvie, Chicago, IL, United States |
| Jeffrey Waring | Abbvie, Chicago, IL, United States |
| Nizar Smaoui | Abbvie, Chicago, IL, United States |
| Fedik Rahimov | Abbvie, Chicago, IL, United States |

Anne Lehtonen		Abbvie, Chicago, IL, United States
Keith Usiskin		Celgene, Summit, NJ, United States/ Bristol Myers Squibb, New York, NY, United States
Tim Lu			Genentech, San Francisco, CA, United States
Natalie Bowers		Genentech, San Francisco, CA, United States
Danny Oh		Genentech, San Francisco, CA, United States
Sarah Pendergrass	Genentech, San Francisco, CA, United States
Kirsi Kalpala		Pfizer, New York, NY, United States
Melissa Miller		Pfizer, New York, NY, United States
Xinli Hu			Pfizer, New York, NY, United States
Linda McCarthy		GlaxoSmithKline, Brentford, United Kingdom
Onuralp Soylemez	Merck, Kenilworth, NJ, United States
Mark Daly		Institute for Molecular Medicine Finland, HiLIFE, University of Helsinki, Finland

**Rheumatology Group**
Kari Eklund		Hospital District of Helsinki and Uusimaa, Helsinki, Finland
Antti Palomäki		Hospital District of Southwest Finland, Turku, Finland
Pia Isomäki		Pirkanmaa Hospital District, Tampere, Finland
Laura Pirilä		Hospital District of Southwest Finland, Turku, Finland
Oili Kaipiainen-Seppänen	Northern Savo Hospital District, Kuopio, Finland
Johanna Huhtakangas	Northern Ostrobothnia Hospital District, Oulu, Finland
Ali Abbasi		Abbvie, Chicago, IL, United States
Jeffrey Waring		Abbvie, Chicago, IL, United States
Fedik Rahimov		Abbvie, Chicago, IL, United States
Apinya Lertratanakul	Abbvie, Chicago, IL, United States
Nizar Smaoui		Abbvie, Chicago, IL, United States
Anne Lehtonen		Abbvie, Chicago, IL, United States
David Close		Astra Zeneca, Cambridge, United Kingdom
Marla Hochfeld		Celgene, Summit, NJ, United States/ Bristol Myers Squibb, New York, NY, United States
Natalie Bowers		Genentech, San Francisco, CA, United States
Sarah Pendergrass	Genentech, San Francisco, CA, United States
Onuralp Soylemez	Merck, Kenilworth, NJ, United States
Kirsi Kalpala		Pfizer, New York, NY, United States
Nan Bing		Pfizer, New York, NY, United States
Xinli Hu			Pfizer, New York, NY, United States
Jorge Esparza Gordillo	GlaxoSmithKline, Brentford, United Kingdom
Kirsi Auro		GlaxoSmithKline, Brentford, United Kingdom
Dawn Waterworth	Janssen Biotech, Beerse, Belgium
Nina Mars		Institute for Molecular Medicine Finland, HiLIFE, Helsinki, Finland

**Pulmonology Group**
Tarja Laitinen		Pirkanmaa Hospital District, Tampere, Finland
Margit Pelkonen		Northern Savo Hospital District, Kuopio, Finland
Paula Kauppi		Hospital District of Helsinki and Uusimaa, Helsinki, Finland
Hannu Kankaanranta	Pirkanmaa Hospital District, Tampere, Finland
Terttu Harju		Northern Ostrobothnia Hospital District, Oulu, Finland
Riitta Lahesmaa		Hospital District of Southwest Finland, Turku, Finland
Nizar Smaoui		Abbvie, Chicago, IL, United States
Alex Mackay		Astra Zeneca, Cambridge, United Kingdom
Glenda Lassi		Astra Zeneca, Cambridge, United Kingdom
Susan Eaton		Biogen, Cambridge, MA, United States
Steven Greenberg	Celgene, Summit, NJ, United States/ Bristol Myers Squibb, New York, NY, United States
Hubert Chen		Genentech, San Francisco, CA, United States
Sarah Pendergrass	Genentech, San Francisco, CA, United States

Natalie Bowers Genentech, San Francisco, CA, United States
Joanna Betts GlaxoSmithKline, Brentford, United Kingdom
Soumitra Ghosh GlaxoSmithKline, Brentford, United Kingdom
Kirsi Auro GlaxoSmithKline, Brentford, United Kingdom
Rajashree Mishra GlaxoSmithKline, Brentford, United Kingdom
Sina Rüeger Institute for Molecular Medicine Finland, HiLIFE, University of Helsinki, Finland

**Cardiometabolic Diseases Group**

Teemu Niiranen The National Institute of Health and Welfare Helsinki, Finland
Felix Vaura The National Institute of Health and Welfare Helsinki, Finland
Veikko Salomaa The National Institute of Health and Welfare Helsinki, Finland
Markus Juonala Hospital District of Southwest Finland, Turku, Finland
Kaj Metsärinne Hospital District of Southwest Finland, Turku, Finland
Mika Kähönen Pirkanmaa Hospital District, Tampere, Finland
Juhani Junttila Northern Ostrobothnia Hospital District, Oulu, Finland
Markku Laakso Northern Savo Hospital District, Kuopio, Finland
Jussi Pihlajamäki Northern Savo Hospital District, Kuopio, Finland
Daniel Gordin Hospital District of Helsinki and Uusimaa, Helsinki, Finland
Juha Sinisalo Hospital District of Helsinki and Uusimaa, Helsinki, Finland
Marja-Riitta Taskinen Hospital District of Helsinki and Uusimaa, Helsinki, Finland
Tiinamaija Tuomi Hospital District of Helsinki and Uusimaa, Helsinki, Finland
Jari Laukkanen Central Finland Health Care District, Jyväskylä, Finland
Benjamin Challis Astra Zeneca, Cambridge, United Kingdom
Dirk Paul Astra Zeneca, Cambridge, United Kingdom
Julie Hunkapiller Genentech, San Francisco, CA, United States
Natalie Bowers Genentech, San Francisco, CA, United States
Sarah Pendergrass Genentech, San Francisco, CA, United States
Onuralp Soylemez Merck, Kenilworth, NJ, United States
Jaakko Parkkinen Pfizer, New York, NY, United States
Melissa Miller Pfizer, New York, NY, United States
Russell Miller Pfizer, New York, NY, United States
Audrey Chu GlaxoSmithKline, Brentford, United Kingdom
Kirsi Auro GlaxoSmithKline, Brentford, United Kingdom
Keith Usiskin Celgene, Summit, NJ, United States/ Bristol Myers Squibb, New York, NY, United States
Amanda Elliott Institute for Molecular Medicine Finland, HiLIFE, University of Helsinki, Finland / Broad Institute, Cambridge, MA, United States
Joel Rämö Institute for Molecular Medicine Finland, HiLIFE, University of Helsinki, Finland
Samuli Ripatti Institute for Molecular Medicine Finland, HiLIFE, University of Helsinki, Finland
Mary Pat Reeve Institute for Molecular Medicine Finland, HiLIFE, University of Helsinki, Finland
Sanni Ruotsalainen Institute for Molecular Medicine Finland, HiLIFE, University of Helsinki, Finland

**Oncology Group**

Tuomo Meretoja Hospital District of Helsinki and Uusimaa, Helsinki, Finland
Heikki Joensuu Hospital District of Helsinki and Uusimaa, Helsinki, Finland
Olli Carpén Hospital District of Helsinki and Uusimaa, Helsinki, Finland
Lauri Aaltonen Hospital District of Helsinki and Uusimaa, Helsinki, Finland
Johanna Mattson Hospital District of Helsinki and Uusimaa, Helsinki, Finland
Annika Auranen Pirkanmaa Hospital District , Tampere, Finland
Peeter Karihtala Northern Ostrobothnia Hospital District, Oulu, Finland
Saila Kauppila Northern Ostrobothnia Hospital District, Oulu, Finland
Päivi Auvinen Northern Savo Hospital District, Kuopio, Finland
Klaus Elenius Hospital District of Southwest Finland, Turku, Finland
Johanna Schleutker Hospital District of Southwest Finland, Turku, Finland
Relja Popovic Abbvie, Chicago, IL, United States
Jeffrey Waring Abbvie, Chicago, IL, United States

| | |
|---|---|
| Bridget Riley-Gillis | Abbvie, Chicago, IL, United States |
| Anne Lehtonen | Abbvie, Chicago, IL, United States |
| Jennifer Schutzman | Genentech, San Francisco, CA, United States |
| Julie Hunkapiller | Genentech, San Francisco, CA, United States |
| Natalie Bowers | Genentech, San Francisco, CA, United States |
| Sarah Pendergrass | Genentech, San Francisco, CA, United States |
| Andrey Loboda | Merck, Kenilworth, NJ, United States |
| Aparna Chhibber | Merck, Kenilworth, NJ, United States |
| Heli Lehtonen | Pfizer, New York, NY, United States |
| Stefan McDonough | Pfizer, New York, NY, United States |
| Marika Crohns | Sanofi, Paris, France |
| Sauli Vuoti | Sanofi, Paris, France |
| Diptee Kulkarni | GlaxoSmithKline, Brentford, United Kingdom |
| Kirsi Auro | GlaxoSmithKline, Brentford, United Kingdom |
| Esa Pitkänen | Institute for Molecular Medicine Finland, HiLIFE, University of Helsinki, Finland |
| Nina Mars | Institute for Molecular Medicine Finland, HiLIFE, University of Helsinki, Finland |
| Mark Daly | Institute for Molecular Medicine Finland, HiLIFE, University of Helsinki, Finland |

**Opthalmology Group**

| | |
|---|---|
| Kai Kaarniranta | Northern Savo Hospital District, Kuopio, Finland |
| Joni A Turunen | Hospital District of Helsinki and Uusimaa, Helsinki, Finland |
| Terhi Ollila | Hospital District of Helsinki and Uusimaa, Helsinki, Finland |
| Sanna Seitsonen | Hospital District of Helsinki and Uusimaa, Helsinki, Finland |
| Hannu Uusitalo | Pirkanmaa Hospital District, Tampere, Finland |
| Vesa Aaltonen | Hospital District of Southwest Finland, Turku, Finland |
| Hannele Uusitalo-Järvinen | Pirkanmaa Hospital District, Tampere, Finland |
| Marja Luodonpää | Northern Ostrobothnia Hospital District, Oulu, Finland |
| Nina Hautala | Northern Ostrobothnia Hospital District, Oulu, Finland |
| Mengzhen Liu | Abbvie, Chicago, IL, United States |
| Heiko Runz | Biogen, Cambridge, MA, United States |
| Stephanie Loomis | Biogen, Cambridge, MA, United States |
| Erich Strauss | Genentech, San Francisco, CA, United States |
| Natalie Bowers | Genentech, San Francisco, CA, United States |
| Hao Chen | Genentech, San Francisco, CA, United States |
| Sarah Pendergrass | Genentech, San Francisco, CA, United States |
| Anna Podgornaia | Merck, Kenilworth, NJ, United States |
| Juha Karjalainen | Institute for Molecular Medicine Finland, HiLIFE, University of Helsinki, Finland / Broad Institute, Cambridge, MA, United States |
| Esa Pitkänen | Institute for Molecular Medicine Finland, HiLIFE, University of Helsinki, Finland |

**Dermatology Group**

| | |
|---|---|
| Kaisa Tasanen | Northern Ostrobothnia Hospital District, Oulu, Finland |
| Laura Huilaja | Northern Ostrobothnia Hospital District, Oulu, Finland |
| Katariina Hannula-Jouppi | Hospital District of Helsinki and Uusimaa, Helsinki, Finland |
| Teea Salmi | Pirkanmaa Hospital District, Tampere, Finland |
| Sirkku Peltonen | Hospital District of Southwest Finland, Turku, Finland |
| Leena Koulu | Hospital District of Southwest Finland, Turku, Finland |
| Kirsi Kalpala | Pfizer, New York, NY, United States |
| Ying Wu | Pfizer, New York, NY, United States |
| David Choy | Genentech, San Francisco, CA, United States |
| Sarah Pendergrass | Genentech, San Francisco, CA, United States |
| Nizar Smaoui | Abbvie, Chicago, IL, United States |
| Fedik Rahimov | Abbvie, Chicago, IL, United States |
| Anne Lehtonen | Abbvie, Chicago, IL, United States |
| Dawn Waterworth | Janssen Biotech, Beerse, Belgium |

**Odontology Group**

| | |
|---|---|
| Pirkko Pussinen | Hospital District of Helsinki and Uusimaa, Helsinki, Finland |
| Aino Salminen | Hospital District of Helsinki and Uusimaa, Helsinki, Finland |
| Tuula Salo | Hospital District of Helsinki and Uusimaa, Helsinki, Finland |
| David Rice | Hospital District of Helsinki and Uusimaa, Helsinki, Finland |
| Pekka Nieminen | Hospital District of Helsinki and Uusimaa, Helsinki, Finland |
| Ulla Palotie | Hospital District of Helsinki and Uusimaa, Helsinki, Finland |
| Juha Sinisalo | Hospital District of Helsinki and Uusimaa, Helsinki, Finland |
| Maria Siponen | Northern Savo Hospital District, Kuopio, Finland |
| Liisa Suominen | Northern Savo Hospital District, Kuopio, Finland |
| Päivi Mäntylä | Northern Savo Hospital District, Kuopio, Finland |
| Ulvi Gursoy | Hospital District of Southwest Finland, Turku, Finland |
| Vuokko Anttonen | Northern Ostrobothnia Hospital District, Oulu, Finland |
| Kirsi Sipilä | Northern Ostrobothnia Hospital District, Oulu, Finland |
| Sarah Pendergrass | Genentech, San Francisco, CA, United States |

**Women's Health and Reproduction Group**

| | |
|---|---|
| Hannele Laivuori | Institute for Molecular Medicine Finland, HiLIFE, University of Helsinki, Finland |
| Venla Kurra | Pirkanmaa Hospital District, Tampere, Finland |
| Oskari Heikinheimo | Hospital District of Helsinki and Uusimaa, Helsinki, Finland |
| Ilkka Kalliala | Hospital District of Helsinki and Uusimaa, Helsinki, Finland |
| Laura Kotaniemi-Talonen | Pirkanmaa Hospital District, Tampere, Finland |
| Kari Nieminen | Pirkanmaa Hospital District, Tampere, Finland |
| Päivi Polo | Hospital District of Southwest Finland, Turku, Finland |
| Kaarin Mäkikallio | Hospital District of Southwest Finland, Turku, Finland |
| Eeva Ekholm | Hospital District of Southwest Finland, Turku, Finland |
| Marja Vääräsmäki | Northern Ostrobothnia Hospital District, Oulu, Finland |
| Outi Uimari | Northern Ostrobothnia Hospital District, Oulu, Finland |
| Laure Morin-Papunen | Northern Ostrobothnia Hospital District, Oulu, Finland |
| Marjo Tuppurainen | Northern Savo Hospital District, Kuopio, Finland |
| Katja Kivinen | Institute for Molecular Medicine Finland, HiLIFE, University of Helsinki, Finland |
| Elisabeth Widen | Institute for Molecular Medicine Finland, HiLIFE, University of Helsinki, Finland |
| Taru Tukiainen | Institute for Molecular Medicine Finland, HiLIFE, University of Helsinki, Finland |
| Mary Pat Reeve | Institute for Molecular Medicine Finland, HiLIFE, University of Helsinki, Finland |
| Mark Daly | Institute for Molecular Medicine Finland, HiLIFE, University of Helsinki, Finland |
| Liu Aoxing | Institute for Molecular Medicine Finland, HiLIFE, University of Helsinki, Finland |
| Eija Laakkonen | University of Jyväskylä, Jyväskylä, Finland |
| Niko Välimäki | University of Helsinki, Helsinki, Finland |
| Lauri Aaltonen | Hospital District of Helsinki and Uusimaa, Helsinki, Finland |
| Johannes Kettunen | Northern Ostrobothnia Hospital District, Oulu, Finland |
| Mikko Arvas | Finnish Red Cross Blood Service, Helsinki, Finland |
| Jeffrey Waring | Abbvie, Chicago, IL, United States |
| Bridget Riley-Gillis | Abbvie, Chicago, IL, United States |
| Mengzhen Liu | Abbvie, Chicago, IL, United States |
| Janet Kumar | GlaxoSmithKline, Brentford, United Kingdom |
| Kirsi Auro | GlaxoSmithKline, Brentford, United Kingdom |
| Andrea Ganna | Institute for Molecular Medicine Finland, HiLIFE, University of Helsinki, Finland |
| Sarah Pendergrass | Genentech, San Francisco, CA, United States |

**FinnGen Analysis working group**

| | |
|---|---|
| Justin Wade Davis | Abbvie, Chicago, IL, United States |
| Bridget Riley-Gillis | Abbvie, Chicago, IL, United States |
| Reza Hammond | Abbvie, Chicago, IL, United States |
| Fedik Rahimov | Abbvie, Chicago, IL, United States |
| Sahar Esmaeeli | Abbvie, Chicago, IL, United States |
| Mengzhen Liu | Abbvie, Chicago, IL, United States |

Slavé Petrovski Astra Zeneca, Cambridge, United Kingdom
Eleonor Wigmore Astra Zeneca, Cambridge, United Kingdom
Adele Mitchell Biogen, Cambridge, MA, United States
Benjamin Sun Biogen, Cambridge, MA, United States
Ellen Tsai Biogen, Cambridge, MA, United States
Denis Baird Biogen, Cambridge, MA, United States
Paola Bronson Biogen, Cambridge, MA, United States
Ruoyu Tian Biogen, Cambridge, MA, United States
Stephanie Loomis Biogen, Cambridge, MA, United States
Yunfeng Huang Biogen, Cambridge, MA, United States
Joseph Maranville Celgene, Summit, NJ, United States/ Bristol Myers Squibb, New York, NY, United States
Shameek Biswas Celgene, Summit, NJ, United States/ Bristol Myers Squibb, New York, NY, United States
Elmutaz Mohammed Celgene, Summit, NJ, United States/ Bristol Myers Squibb, New York, NY, United States
Samir Wadhawan Celgene, Summit, NJ, United States/ Bristol Myers Squibb, New York, NY, United States
Erika Kvikstad Celgene, Summit, NJ, United States/ Bristol Myers Squibb, New York, NY, United States
Minal Caliskan Celgene, Summit, NJ, United States/ Bristol Myers Squibb, New York, NY, United States
Diana Chang Genentech, San Francisco, CA, United States
Julie Hunkapiller Genentech, San Francisco, CA, United States
Tushar Bhangale Genentech, San Francisco, CA, United States
Natalie Bowers Genentech, San Francisco, CA, United States
Sarah Pendergrass Genentech, San Francisco, CA, United States
Kirill Shkura Merck, Kenilworth, NJ, United States
Victor Neduva Merck, Kenilworth, NJ, United States
Xing Chen Pfizer, New York, NY, United States
Åsa Hedman Pfizer, New York, NY, United States
Karen S King GlaxoSmithKline, Brentford, United Kingdom
Padhraig Gormley GlaxoSmithKline, Brentford, United Kingdom
Jimmy Liu GlaxoSmithKline, Brentford, United Kingdom
Clarence Wang Sanofi, Paris, France
Ethan Xu Sanofi, Paris, France
Franck Auge Sanofi, Paris, France
Clement Chatelain Sanofi, Paris, France
Deepak Rajpal Sanofi, Paris, France
Dongyu Liu Sanofi, Paris, France
Katherine Call Sanofi, Paris, France
Tai-He Xia Sanofi, Paris, France
Beryl Cummings Maze Therapeutics, San Francisco, CA, United States
Matt Brauer Maze Therapeutics, San Francisco, CA, United States
Huilei Xu Novartis, Basel, Switzerland
Amy Cole Novartis, Basel, Switzerland
Jonathan Chung Novartis, Basel, Switzerland
Jaison Jacob Novartis, Basel, Switzerland
Katrina de Lange Novartis, Basel, Switzerland
Jonas Zierer Novartis, Basel, Switzerland
Mitja Kurki Institute for Molecular Medicine Finland, HiLIFE, University of Helsinki, Finland / Broad Institute, Cambridge, MA, United States
Samuli Ripatti Institute for Molecular Medicine Finland, HiLIFE, University of Helsinki, Finland
Mark Daly Institute for Molecular Medicine Finland, HiLIFE, University of Helsinki, Finland
Juha Karjalainen Institute for Molecular Medicine Finland, HiLIFE, University of Helsinki, Finland / Broad Institute, Cambridge, MA, United States

Aki Havulinna Institute for Molecular Medicine Finland, HiLIFE, University of Helsinki, Finland
Juha Mehtonen Institute for Molecular Medicine Finland, HiLIFE, University of Helsinki, Finland
Priit Palta Institute for Molecular Medicine Finland, HiLIFE, University of Helsinki, Finland
Shabbeer Hassan Institute for Molecular Medicine Finland, HiLIFE, University of Helsinki, Finland
Pietro Della Briotta Parolo Institute for Molecular Medicine Finland, HiLIFE, University of Helsinki, Finland
Wei Zhou Broad Institute, Cambridge, MA, United States
Mutaamba Maasha Broad Institute, Cambridge, MA, United States
Shabbeer Hassan Institute for Molecular Medicine Finland, HiLIFE, University of Helsinki, Finland
Susanna Lemmelä Institute for Molecular Medicine Finland, HiLIFE, University of Helsinki, Finland
Manuel Rivas University of Stanford, Stanford, CA, United States
Aarno Palotie Institute for Molecular Medicine Finland, HiLIFE, University of Helsinki, Finland
Arto Lehisto Institute for Molecular Medicine Finland, HiLIFE, University of Helsinki, Finland
Andrea Ganna Institute for Molecular Medicine Finland, HiLIFE, University of Helsinki, Finland
Vincent Llorens Institute for Molecular Medicine Finland, HiLIFE, University of Helsinki, Finland
Hannele Laivuori Institute for Molecular Medicine Finland, HiLIFE, University of Helsinki, Finland
Mari E Niemi Institute for Molecular Medicine Finland, HiLIFE, University of Helsinki, Finland
Taru Tukiainen Institute for Molecular Medicine Finland, HiLIFE, University of Helsinki, Finland
Mary Pat Reeve Institute for Molecular Medicine Finland, HiLIFE, University of Helsinki, Finland
Henrike Heyne Institute for Molecular Medicine Finland, HiLIFE, University of Helsinki, Finland
Nina Mars Institute for Molecular Medicine Finland, HiLIFE, University of Helsinki, Finland
Kimmo Palin University of Helsinki, Helsinki, Finland
Javier Garcia-Tabuenca University of Tampere, Tampere, Finland
Harri Siirtola University of Tampere, Tampere, Finland
Tuomo Kiiskinen Institute for Molecular Medicine Finland, HiLIFE, University of Helsinki, Finland
Jiwoo Lee Institute for Molecular Medicine Finland, HiLIFE, University of Helsinki, Finland / Broad Institute, Cambridge, MA, United States
Kristin Tsuo Institute for Molecular Medicine Finland, HiLIFE, University of Helsinki, Finland / Broad Institute, Cambridge, MA, United States
Amanda Elliott Institute for Molecular Medicine Finland, HiLIFE, University of Helsinki, Finland / Broad Institute, Cambridge, MA, United States
Kati Kristiansson THL Biobank / The National Institute of Health and Welfare Helsinki, Finland
Mikko Arvas Finnish Red Cross Blood Service / Finnish Hematology Registry and Clinical Biobank, Helsinki, Finland
Kati Hyvärinen Finnish Red Cross Blood Service, Helsinki, Finland
Jarmo Ritari Finnish Red Cross Blood Service, Helsinki, Finland
Miika Koskinen Helsinki Biobank / Helsinki University and Hospital District of Helsinki and Uusimaa, Helsinki
Olli Carpén Helsinki Biobank / Helsinki University and Hospital District of Helsinki and Uusimaa, Helsinki
Johannes Kettunen Northern Finland Biobank Borealis / University of Oulu / Northern Ostrobothnia Hospital District, Oulu, Finland
Katri Pylkäs University of Oulu, Oulu, Finland
Eeva Sliz University of Oulu, Oulu, Finland
Minna Karjalainen University of Oulu, Oulu, Finland
Tuomo Mantere Northern Finland Biobank Borealis / University of Oulu / Northern Ostrobothnia Hospital District, Oulu, Finland
Eeva Kangasniemi Finnish Clinical Biobank Tampere / University of Tampere / Pirkanmaa Hospital District, Tampere, Finland
Sami Heikkinen University of Eastern Finland, Kuopio, Finland
Arto Mannermaa Biobank of Eastern Finland / University of Eastern Finland / Northern Savo Hospital District, Kuopio, Finland
Eija Laakkonen University of Jyväskylä, Jyväskylä, Finland
Samuel Heron University of Turku, Turku, Finland
Dhanaprakash Jambulingam University of Turku, Turku, Finland
Venkat Subramaniam Rathinakannan University of Turku, Turku, Finland

Nina Pitkänen Auria Biobank / University of Turku / Hospital District of Southwest Finland, Turku, Finland

#### Biobank directors

Lila Kallio Auria Biobank / University of Turku / Hospital District of Southwest Finland, Turku, Finland
Sirpa Soini THL Biobank / The National Institute of Health and Welfare Helsinki, Finland
Jukka Partanen Finnish Red Cross Blood Service / Finnish Hematology Registry and Clinical Biobank, Helsinki, Finland
Eero Punkka Helsinki Biobank / Helsinki University and Hospital District of Helsinki and Uusimaa, Helsinki
Raisa Serpi Northern Finland Biobank Borealis / University of Oulu / Northern Ostrobothnia Hospital District, Oulu, Finland
TBC Finnish Clinical Biobank Tampere / University of Tampere / Pirkanmaa Hospital District, Tampere, Finland
Veli-Matti Kosma Biobank of Eastern Finland / University of Eastern Finland / Northern Savo Hospital District, Kuopio, Finland
Teijo Kuopio Central Finland Biobank / University of Jyväskylä / Central Finland Health Care District, Jyväskylä, Finland

#### FinnGen Teams

##### Administration

Anu Jalanko Institute for Molecular Medicine Finland, HiLIFE, University of Helsinki, Finland
Huei-Yi Shen Institute for Molecular Medicine Finland, HiLIFE, University of Helsinki, Finland
Risto Kajanne Institute for Molecular Medicine Finland, HiLIFE, University of Helsinki, Finland
Mervi Aavikko Institute for Molecular Medicine Finland, HiLIFE, University of Helsinki, Finland

##### Analysis

Mitja Kurki Institute for Molecular Medicine Finland, HiLIFE, University of Helsinki, Finland / Broad Institute, Cambridge, MA, United States
Juha Karjalainen Institute for Molecular Medicine Finland, HiLIFE, University of Helsinki, Finland / Broad Institute, Cambridge, MA, United States
Pietro Della Briotta Parolo Institute for Molecular Medicine Finland, HiLIFE, University of Helsinki, Finland
Arto Lehisto Institute for Molecular Medicine Finland, HiLIFE, University of Helsinki, Finland
Juha Mehtonen Institute for Molecular Medicine Finland, HiLIFE, University of Helsinki, Finland
Wei Zhou Broad Institute, Cambridge, MA, United States
Masahiro Kanai Broad Institute, Cambridge, MA, United States
Mutaamba Maasha Broad Institute, Cambridge, MA, United States

##### Clinical Endpoint Development

Hannele Laivuori Institute for Molecular Medicine Finland, HiLIFE, University of Helsinki, Finland
Aki Havulinna Institute for Molecular Medicine Finland, HiLIFE, University of Helsinki, Finland
Susanna Lemmelä Institute for Molecular Medicine Finland, HiLIFE, University of Helsinki, Finland
Tuomo Kiiskinen Institute for Molecular Medicine Finland, HiLIFE, University of Helsinki, Finland
L. Elisa Lahtela Institute for Molecular Medicine Finland, HiLIFE, University of Helsinki, Finland

##### Communication

Mari Kaunisto Institute for Molecular Medicine Finland, HiLIFE, University of Helsinki, Finland

##### E-Science

Elina Kilpeläinen Institute for Molecular Medicine Finland, HiLIFE, University of Helsinki, Finland
Timo P. Sipilä Institute for Molecular Medicine Finland, HiLIFE, University of Helsinki, Finland
Georg Brein Institute for Molecular Medicine Finland, HiLIFE, University of Helsinki, Finland

Oluwaseun Alexander Dada Institute for Molecular Medicine Finland, HiLIFE, University of Helsinki, Finland
Awaisa Ghazal Institute for Molecular Medicine Finland, HiLIFE, University of Helsinki, Finland
Anastasia Shcherban Institute for Molecular Medicine Finland, HiLIFE, University of Helsinki, Finland

**Genotyping**
Kati Donner Institute for Molecular Medicine Finland, HiLIFE, University of Helsinki, Finland
Timo P. Sipilä Institute for Molecular Medicine Finland, HiLIFE, University of Helsinki, Finland

**Sample Collection Coordination**
Anu Loukola Helsinki Biobank / Helsinki University and Hospital District of Helsinki and Uusimaa, Helsinki

**Sample Logistics**
Päivi Laiho THL Biobank / The National Institute of Health and Welfare Helsinki, Finland
Tuuli Sistonen THL Biobank / The National Institute of Health and Welfare Helsinki, Finland
Essi Kaiharju THL Biobank / The National Institute of Health and Welfare Helsinki, Finland
Markku Laukkanen THL Biobank / The National Institute of Health and Welfare Helsinki, Finland
Elina Järvensivu THL Biobank / The National Institute of Health and Welfare Helsinki, Finland
Sini Lähteenmäki THL Biobank / The National Institute of Health and Welfare Helsinki, Finland
Lotta Männikkö THL Biobank / The National Institute of Health and Welfare Helsinki, Finland
Regis Wong THL Biobank / The National Institute of Health and Welfare Helsinki, Finland

**Registry Data Operations**
Hannele Mattsson THL Biobank / The National Institute of Health and Welfare Helsinki, Finland
Kati Kristiansson THL Biobank / The National Institute of Health and Welfare Helsinki, Finland
Susanna Lemmelä Institute for Molecular Medicine Finland, HiLIFE, University of Helsinki, Finland
Sami Koskelainen THL Biobank / The National Institute of Health and Welfare Helsinki, Finland
Tero Hiekkalinna THL Biobank / The National Institute of Health and Welfare Helsinki, Finland
Teemu Paajanen THL Biobank / The National Institute of Health and Welfare Helsinki, Finland

**Sequencing Informatics**
Priit Palta Institute for Molecular Medicine Finland, HiLIFE, University of Helsinki, Finland
Kalle Pärn Institute for Molecular Medicine Finland, HiLIFE, University of Helsinki, Finland
Mart Kals Institute for Molecular Medicine Finland, HiLIFE, University of Helsinki, Finland
Shuang Luo Institute for Molecular Medicine Finland, HiLIFE, University of Helsinki, Finland
Vishal Sinha Institute for Molecular Medicine Finland, HiLIFE, University of Helsinki, Finland

**Trajectory**
Tarja Laitinen Pirkanmaa Hospital District, Tampere, Finland
Mary Pat Reeve Institute for Molecular Medicine Finland, HiLIFE, University of Helsinki, Finland
Harri Siirtola University of Tampere, Tampere, Finland
Javier Gracia-Tabuenca University of Tampere, Tampere, Finland
Mika Helminen University of Tampere, Tampere, Finland
Tiina Luukkaala University of Tampere, Tampere, Finland
Iida Vähätalo University of Tampere, Tampere, Finland

**Data protection officer**
Tero Jyrhämä Institute for Molecular Medicine Finland, HiLIFE, University of Helsinki, Finland

**FINBB - Finnish biobank cooperative**
Marco Hautalahti
Johanna Mäkelä
Laura Mustaniemi

Mirkka Koivusalo
Sarah Smith
Tom Southerington

**Bibliography for Supplementary Table 11.** Each reference starts with the PMID before a colon. The references are sorted by their PMIDs.


1. 2307: Sakakibara, S., Yamaguchi, K., Hosokawa, Y., Kohashi, N. & Ueda, I. Purification and some properties of rat liver cysteine oxidase (cysteine dioxygenase). *Biochim Biophys Acta* **422,** 273–279 (1976).
2. 8453: Enoch, H. G., Catalá, A. & Strittmatter, P. Mechanism of rat liver microsomal stearyl-CoA desaturase. Studies of the substrate specificity, enzyme-substrate interactions, and the function of lipid. *J Biol Chem* **251,** 5095–5103 (1976).
3. 17401: Vessey, D. A. & Zakim, D. Characterization of microsomal choloyl-coenzyme A synthetase. *Biochem J* **163,** 357–362 (1977).
4. 33715: Fluharty, A. L., Stevens, R. L., Goldstein, E. B. & Kihara, H. The activity of arylsulfatase A and B on tyrosine O-sulfates. *Biochim Biophys Acta* **566,** 321–326 (1979).
5. 218932: Spector, T., Jones, T. E. & Miller, R. L. Reaction mechanism and specificity of human GMP reductase. Substrates, inhibitors, activators, and inactivators. *J Biol Chem* **254,** 2308–2315 (1979).
6. 226968: Brown, M. S. & Goldstein, J. L. Receptor-mediated endocytosis: insights from the lipoprotein receptor system. *Proc Natl Acad Sci U S A* **76,** 3330–3337 (1979).
7. 240826: Spector, T. Studies with GMP synthetase from Ehrlich ascites cells. Purification, properties, and interactions with nucleotide analogs. *J Biol Chem* **250,** 7372–7376 (1975).
8. 873932: Lindblad, B., Lindstedt, G., Lindstedt, S. & Rundgren, M. Purification and some properties of human 4-hydroxyphenylpyruvate dioxygenase (I). *J Biol Chem* **252,** 5073–5084 (1977).
9. 1281977: Rip, J. W., Coulter-Mackie, M. B., Rupar, C. A. & Gordon, B. A. Purification and structure of human liver aspartylglucosaminidase. *Biochem J* **288 ( Pt 3),** 1005–1010 (1992).
10. 1302044: Gu, L., Gonzalez, F. J., Kalow, W. & Tang, B. K. Biotransformation of caffeine, paraxanthine, theobromine and theophylline by cDNA-expressed human CYP1A2 and CYP2E1. *Pharmacogenetics* **2,** 73–77 (1992).
11. 1550553: Carpenter, K., Pollitt, R. J. & Middleton, B. Human liver long-chain 3-hydroxyacyl-coenzyme A dehydrogenase is a multifunctional membrane-bound beta-oxidation enzyme of mitochondria. *Biochem Biophys Res Commun* **183,** 443–448 (1992).
12. 1730632: Izai, K., Uchida, Y., Orii, T., Yamamoto, S. & Hashimoto, T. Novel fatty acid beta-oxidation enzymes in rat liver mitochondria. I. Purification and properties of very-long-chain acyl-coenzyme A dehydrogenase. *J Biol Chem* **267,** 1027–1033 (1992).
13. 1865762: Gavino, G. R. & Gavino, V. C. Rat liver outer mitochondrial carnitine palmitoyltransferase activity towards long-chain polyunsaturated fatty acids and their CoA esters. *Lipids* **26,** 266–270 (1991).
14. 1973834: Rettenmeier, R., Natt, E., Zentgraf, H. & Scherer, G. Isolation and characterization of the human tyrosine aminotransferase gene. *Nucleic Acids Res* **18,** 3853–3861 (1990).
15. 1999428: Melkerson-Watson, L. J. & Sweeley, C. C. Purification to apparent homogeneity by immunoaffinity chromatography and partial characterization of the GM3 ganglioside-forming enzyme, CMP-sialic acid:lactosylceramide alpha 2,3-sialyltransferase (SAT-1), from rat liver Golgi. *J Biol Chem* **266,** 4448–4457 (1991).
16. 2157703: Höglund, L. & Reichard, P. Cytoplasmic 5'(3')-nucleotidase from human placenta. *J Biol Chem* **265,** 6589–6595 (1990).
17. 2211729: Small, W. C. & Jones, M. E. Pyrroline 5-carboxylate dehydrogenase of the mitochondrial matrix of rat liver. Purification, physical and kinetic characteristics. *J Biol Chem* **265,** 18668–18672 (1990).
18. 2340092: Chang, C. Y., Wu, D. A., Mohandas, T. K. & Chung, B. C. Structure, sequence, chromosomal location, and evolution of the human ferredoxin gene family. *DNA Cell Biol* **9,** 205–212 (1990).
19. 2351134: Bloisi, W. *et al.* Purification and properties of carnitine acetyltransferase from human liver. *Eur J Biochem* **189,** 539–546 (1990).
20. 2444024: Rask, L., Anundi, H., Fohlman, J. & Peterson, P. A. The complete amino acid sequence of human serum retinol-binding protein. *Ups J Med Sci* **92,** 115–146 (1987).

21. 2463916: Schoedon, G., Redweik, U. & Curtius, H. C. Purification of GTP cyclohydrolase I from human liver and production of specific monoclonal antibodies. *Eur J Biochem* **178,** 627–634 (1989).
22. 2917560: Van Schaftingen, E. A protein from rat liver confers to glucokinase the property of being antagonistically regulated by fructose 6-phosphate and fructose 1-phosphate. *Eur J Biochem* **179,** 179–184 (1989).
23. 2993784: Andersson, S., Boström, H., Danielsson, H. & Wikvall, K. Purification from rabbit and rat liver of cytochromes P-450 involved in bile acid biosynthesis. *Methods Enzymol* **111,** 364–377 (1985).
24. 3081514: Lenich, A. C. & Goodman, S. I. The purification and characterization of glutaryl-coenzyme A dehydrogenase from porcine and human liver. *J Biol Chem* **261,** 4090–4096 (1986).
25. 3094014: Flink, I. L., Bailey, T. J., Gustafson, T. A., Markham, B. E. & Morkin, E. Complete amino acid sequence of human thyroxine-binding globulin deduced from cloned DNA: close homology to the serine antiproteases. *Proc Natl Acad Sci U S A* **83,** 7708–7712 (1986).
26. 3299377: Hammond, G. L. *et al.* Primary structure of human corticosteroid binding globulin, deduced from hepatic and pulmonary cDNAs, exhibits homology with serine protease inhibitors. *Proc Natl Acad Sci U S A* **84,** 5153–5157 (1987).
27. 3346674: Blakely, R. D., Robinson, M. B., Thompson, R. C. & Coyle, J. T. Hydrolysis of the brain dipeptide N-acetyl-L-aspartyl-L-glutamate: subcellular and regional distribution, ontogeny, and the effect of lesions on N-acetylated-alpha-linked acidic dipeptidase activity. *J Neurochem* **50,** 1200–1209 (1988).
28. 3365415: Maret, W. & Auld, D. S. Purification and characterization of human liver sorbitol dehydrogenase. *Biochemistry* **27,** 1622–1628 (1988).
29. 3473473: Fischer, W. H. & Spiess, J. Identification of a mammalian glutaminyl cyclase converting glutaminyl into pyroglutamyl peptides. *Proc Natl Acad Sci U S A* **84,** 3628–3632 (1987).
30. 3521732: Chung, D. W., Fujikawa, K., McMullen, B. A. & Davie, E. W. Human plasma prekallikrein, a zymogen to a serine protease that contains four tandem repeats. *Biochemistry* **25,** 2410–2417 (1986).
31. 3532105: Weiss, M. J. *et al.* Isolation and characterization of a cDNA encoding a human liver/bone/kidney-type alkaline phosphatase. *Proc Natl Acad Sci U S A* **83,** 7182–7186 (1986).
32. 3540966: Haraguchi, Y. *et al.* Molecular cloning and nucleotide sequence of cDNA for human liver arginase. *Proc Natl Acad Sci U S A* **84,** 412–415 (1987).
33. 3558558: van den Berg, G. A., Kingma, A. W., Elzinga, H. & Muskiet, F. A. Determination of N-(3-acetamidopropyl)pyrrolidin-2-one, a metabolite of spermidine, in urine by isotope dilution mass fragmentography. *J Chromatogr* **383,** 251–258 (1986).
34. 3597357: Finocchiaro, G., Ito, M. & Tanaka, K. Purification and properties of short chain acyl-CoA, medium chain acyl-CoA, and isovaleryl-CoA dehydrogenases from human liver. *J Biol Chem* **262,** 7982–7989 (1987).
35. 3800397: Gross, M. D., Eggen, M. A., Simon, A. M. & Van Pilsum, J. F. The purification and characterization of human kidney L-arginine:glycine amidinotransferase. *Arch Biochem Biophys* **251,** 747–755 (1986).
36. 4150152: Kaufman, S. The phenylalanine hydroxylating system from mammalian liver. *Adv Enzymol Relat Areas Mol Biol* **35,** 245–319 (1971).
37. 4373031: Weinhold, P. A. & Rethy, V. B. The separation, purification, and characterization of ethanolamine kinase and choline kinase from rat liver. *Biochemistry* **13,** 5135–5141 (1974).
38. 4389188: Okamoto, H. & Hayaishi, O. Solubilization and partial purification of kynurenine hydroxylase from mitochondrial outer membrane and its electron donors. *Arch Biochem Biophys* **131,** 603–608 (1969).
39. 4402936: Bosron, W. F. & Prairie, R. L. Triphosphopyridine nucleotide-linked aldehyde reductase. I. Purification and properties of the enzyme from pig kidney cortex. *J Biol Chem* **247,** 4480–4485 (1972).
40. 4585091: Kikuchi, G. The glycine cleavage system: composition, reaction mechanism, and physiological significance. *Mol Cell Biochem* **1,** 169–187 (1973).
41. 4607556: Kanda, Y., Goodman, D. S., Canfield, R. E. & Morgan, F. J. The amino acid sequence of human plasma prealbumin. *J Biol Chem* **249,** 6796–6805 (1974).
42. 4621658: Hiles, R. A. & Henderson, L. M. The partial purification and properties of hydroxylysine kinase from rat liver. *J Biol Chem* **247,** 646–651 (1972).

43. 4623131: Shrawder, E. & Martinez-Carrion, M. Evidence of phenylalanine transaminase activity in the isoenzymes of aspartate transaminase. *J Biol Chem* **247,** 2486–2492 (1972).
44. 4981785: Jost, J. P. & Bock, R. M. Enzymatic hydrolysis of N-substituted aminoacyl transfer ribonucleic acid in yeast. *J Biol Chem* **244,** 5866–5873 (1969).
45. 4990629: Sanada, Y., Suemori, I. & Katunuma, N. Properties of ornithine aminotransferase from rat liver, kidney and small intestine. *Biochim Biophys Acta* **220,** 42–50 (1970).
46. 5044040: Krenitsky, T. A., Neil, S. M., Elion, G. B. & Hitchings, G. H. A comparison of the specificities of xanthine oxidase and aldehyde oxidase. *Arch Biochem Biophys* **150,** 585–599 (1972).
47. 5432795: Lowy, B. & Dorfman, B. Z. Adenylosuccinase activity in human and rabbit erythrocyte lysates. *J Biol Chem* **245,** 3043–3046 (1970).
48. 5773299: Kakimoto, Y., Taniguchi, K. & Sano, I. D-beta-aminoisobutyrate:pyruvate aminotransferase in mammalian liver and excretion of beta-aminoisobutyrate by man. *J Biol Chem* **244,** 335–340 (1969).
49. 6122685: Kozak, E. M. & Tate, S. S. Glutathione-degrading enzymes of microvillus membranes. *J Biol Chem* **257,** 6322–6327 (1982).
50. 6130452: Butler, J. D. & Spielberg, S. P. Accumulation of cystine from glutathione-cysteine mixed disulfide in cystinotic fibroblasts; blockade by an inhibitor of gamma-glutamyl transpeptidase. *Life Sci* **31,** 2563–2570 (1982).
51. 6194510: Bock, H. G., Su, T. S., O'Brien, W. E. & Beaudet, A. L. Sequence for human argininosuccinate synthetase cDNA. *Nucleic Acids Res* **11,** 6505–6512 (1983).
52. 6432877: Yamamoto, S., Yoshimoto, T., Furukawa, M., Horie, T. & Watanabe-Kohno, S. Arachidonate 5-lipoxygenase and its new inhibitors. *J Allergy Clin Immunol* **74,** 349–352 (1984).
53. 6457829: Reitman, M. L. & Kornfeld, S. Lysosomal enzyme targeting. N-Acetylglucosaminylphosphotransferase selectively phosphorylates native lysosomal enzymes. *J Biol Chem* **256,** 11977–11980 (1981).
54. 6772170: Sachan, D. S. & Hoppel, C. L. Carnitine biosynthesis. Hydroxylation of N6-trimethyl-lysine to 3-hydroxy-N6-trimethyl-lysine. *Biochem J* **188,** 529–534 (1980).
55. 6893554: McClard, R. W., Black, M. J., Livingstone, L. R. & Jones, M. E. Isolation and initial characterization of the single polypeptide that synthesizes uridine 5'-monophosphate from orotate in Ehrlich ascites carcinoma. Purification by tandem affinity chromatography of uridine-5'-monophosphate synthase. *Biochemistry* **19,** 4699–4706 (1980).
56. 7284378: Desgranges, C., Razaka, G., Rabaud, M. & Bricaud, H. Catabolism of thymidine in human blood platelets: purification and properties of thymidine phosphorylase. *Biochim Biophys Acta* **654,** 211–218 (1981).
57. 7360517: Goodman, S. I., McCabe, E. R., Fennessey, P. V. & Mace, J. W. Multiple acyl-CoA dehydrogenase deficiency (glutaric aciduria type II) with transient hypersarcosinemia and sarcosinuria; possible inherited deficiency of an electron transfer flavoprotein. *Pediatr Res* **14,** 12–17 (1980).
58. 7417375: Kuusi, T., Saarinen, P. & Nikkilä, E. A. Evidence for the role of hepatic endothelial lipase in the metabolism of plasma high density lipoprotein2 in man. *Atherosclerosis* **36,** 589–593 (1980).
59. 7488099: Watanabe, S. & Uchida, T. Cloning and expression of human uridine phosphorylase. *Biochem Biophys Res Commun* **216,** 265–272 (1995).
60. 7564244: Wanders, R. J. & Mooyer, P. D-2-hydroxyglutaric acidaemia: identification of a new enzyme, D-2-hydroxyglutarate dehydrogenase, localized in mitochondria. *J Inherit Metab Dis* **18,** 194–196 (1995).
61. 7608105: Kimoto, M., Whitley, G. S., Tsuji, H. & Ogawa, T. Detection of NG,NG-dimethylarginine dimethylaminohydrolase in human tissues using a monoclonal antibody. *J Biochem* **117,** 237–238 (1995).
62. 7639524: Luo, M. J., Mao, L. F. & Schulz, H. Short-chain 3-hydroxy-2-methylacyl-CoA dehydrogenase from rat liver: purification and characterization of a novel enzyme of isoleucine metabolism. *Arch Biochem Biophys* **321,** 214–220 (1995).
63. 7690140: Markovich, D., Forgo, J., Stange, G., Biber, J. & Murer, H. Expression cloning of rat renal Na+/SO4(2-) cotransport. *Proc Natl Acad Sci U S A* **90,** 8073–8077 (1993).

64. 7702556: Krupenko, S. A., Wagner, C. & Cook, R. J. Recombinant 10-formyltetrahydrofolate dehydrogenase catalyses both dehydrogenase and hydrolase reactions utilizing the synthetic substrate 10-formyl-5,8-dideazafolate. *Biochem J* **306 ( Pt 3),** 651–655 (1995).
65. 7704034: Goldstein, J. A. & de Morais, S. M. Biochemistry and molecular biology of the human CYP2C subfamily. *Pharmacogenetics* **4,** 285–299 (1994).
66. 7734398: Levy, M. A. *et al.* Cloning, expression and functional characterization of type 1 and type 2 steroid 5 alpha-reductases from Cynomolgus monkey: comparisons with human and rat isoenzymes. *J Steroid Biochem Mol Biol* **52,** 307–319 (1995).
67. 7773568: Dawson, P. A. & Oelkers, P. Bile acid transporters. *Curr Opin Lipidol* **6,** 109–114 (1995).
68. 7860756: Shneider, B. L. *et al.* Cloning and molecular characterization of the ontogeny of a rat ileal sodium-dependent bile acid transporter. *J Clin Invest* **95,** 745–754 (1995).
69. 7896285: Hofmann, K., Düker, M., Fink, T., Lichter, P. & Stoffel, W. Human neutral amino acid transporter ASCT1: structure of the gene (SLC1A4) and localization to chromosome 2p13-p15. *Genomics* **24,** 20–26 (1994).
70. 7957888: Hooper, N. M. Families of zinc metalloproteases. *FEBS Lett* **354,** 1–6 (1994).
71. 8083224: Yokota, H. *et al.* cDNA cloning and chromosome mapping of human dihydropyrimidine dehydrogenase, an enzyme associated with 5-fluorouracil toxicity and congenital thymine uraciluria. *J Biol Chem* **269,** 23192–23196 (1994).
72. 8093002: Veronese, M. E., Burgess, W., Zhu, X. & McManus, M. E. Functional characterization of two human sulphotransferase cDNAs that encode monoamine- and phenol-sulphating forms of phenol sulphotransferase: substrate kinetics, thermal-stability and inhibitor-sensitivity studies. *Biochem J* **302 ( Pt 2),** 497–502 (1994).
73. 8095225: Eugster, H. P., Probst, M., Würgler, F. E. & Sengstag, C. Caffeine, estradiol, and progesterone interact with human CYP1A1 and CYP1A2. Evidence from cDNA-directed expression in Saccharomyces cerevisiae. *Drug Metab Dispos* **21,** 43–49 (1993).
74. 8117268: Aoyama, T. *et al.* Molecular cloning and functional expression of a human peroxisomal acyl-coenzyme A oxidase. *Biochem Biophys Res Commun* **198,** 1113–1118 (1994).
75. 8198549: Petronini, P. G., De Angelis, E., Borghetti, A. F. & Wheeler, K. P. Osmotically inducible uptake of betaine via amino acid transport system A in SV-3T3 cells. *Biochem J* **300 ( Pt 1),** 45–50 (1994).
76. 8241290: Jansen, G. A. & Wanders, R. J. L-2-hydroxyglutarate dehydrogenase: identification of a novel enzyme activity in rat and human liver. Implications for L-2-hydroxyglutaric acidemia. *Biochim Biophys Acta* **1225,** 53–56 (1993).
77. 8399210: Chen, F. *et al.* Characterization of a cloned human dihydrotestosterone/androstanediol UDP-glucuronosyltransferase and its comparison to other steroid isoforms. *Biochemistry* **32,** 10648–10657 (1993).
78. 8422236: Kühn, K., Bertling, W. M. & Emmrich, F. Cloning of a functional cDNA for human cytidine deaminase (CDD) and its use as a marker of monocyte/macrophage differentiation. *Biochem Biophys Res Commun* **190,** 1–7 (1993).
79. 8427875: Kontani, Y., Kaneko, M., Kikugawa, M., Fujimoto, S. & Tamaki, N. Identity of D-3-aminoisobutyrate-pyruvate aminotransferase with alanine-glyoxylate aminotransferase 2. *Biochim Biophys Acta* **1156,** 161–166 (1993).
80. 8486162: Kilponen, J. M. & Hiltunen, J. K. Beta-oxidation of unsaturated fatty acids in humans. Isoforms of delta 3, delta 2-enoyl-CoA isomerase. *FEBS Lett* **322,** 299–303 (1993).
81. 8575776: Durocher, F., Morissette, J., Dufort, I., Simard, J. & Luu-The, V. Genetic linkage mapping of the dehydroepiandrosterone sulfotransferase (STD) gene on the chromosome 19q13.3 region. *Genomics* **29,** 781–783 (1995).
82. 8640791: Jedlitschky, G. *et al.* Transport of glutathione, glucuronate, and sulfate conjugates by the MRP gene-encoded conjugate export pump. *Cancer Res* **56,** 988–994 (1996).
83. 8725270: Corydon, M. J. *et al.* Ethylmalonic aciduria is associated with an amino acid variant of short chain acyl-coenzyme A dehydrogenase. *Pediatr Res* **39,** 1059–1066 (1996).
84. 8798461: Garrow, T. A. Purification, kinetic properties, and cDNA cloning of mammalian betaine-homocysteine methyltransferase. *J Biol Chem* **271,** 22831–22838 (1996).
85. 8798464: Beaulieu, M., Lévesque, E., Hum, D. W. & Bélanger, A. Isolation and characterization of a novel cDNA encoding a human UDP-glucuronosyltransferase active on C19 steroids. *J Biol Chem* **271,** 22855–22862 (1996).

86. 8858963: Sastre, M., Regunathan, S., Galea, E. & Reis, D. J. Agmatinase activity in rat brain: a metabolic pathway for the degradation of agmatine. *J Neurochem* **67,** 1761–1765 (1996).
87. 9124315: Ritzel, M. W. *et al.* Molecular cloning and functional expression of cDNAs encoding a human Na+-nucleoside cotransporter (hCNT1). *Am J Physiol* **272,** C707-714 (1997).
88. 9188469: Cupillard, L., Koumanov, K., Mattéi, M. G., Lazdunski, M. & Lambeau, G. Cloning, chromosomal mapping, and expression of a novel human secretory phospholipase A2. *J Biol Chem* **272,** 15745–15752 (1997).
89. 9233839: Srivenugopal, K. S. & Ali-Osman, F. Activity and distribution of the cysteine prodrug activating enzyme, 5-oxo-L-prolinase, in human normal and tumor tissues. *Cancer Lett* **117,** 105–111 (1997).
90. 9252398: Alperin, E. S. & Shapiro, L. J. Characterization of point mutations in patients with X-linked ichthyosis. Effects on the structure and function of the steroid sulfatase protein. *J Biol Chem* **272,** 20756–20763 (1997).
91. 9452426: Auchus, R. J., Lee, T. C. & Miller, W. L. Cytochrome b5 augments the 17,20-lyase activity of human P450c17 without direct electron transfer. *J Biol Chem* **273,** 3158–3165 (1998).
92. 9516450: Prasad, P. D. *et al.* Cloning and functional expression of a cDNA encoding a mammalian sodium-dependent vitamin transporter mediating the uptake of pantothenate, biotin, and lipoate. *J Biol Chem* **273,** 7501–7506 (1998).
93. 9544928: Henriksen, C. M., Nielsen, J. & Villadsen, J. Cyclization of alpha-aminoadipic acid into the the delta-lactam 6-oxo-piperidine-2-carboxylic acid by Penicillium chrysogenum. *J Antibiot (Tokyo)* **51,** 99–106 (1998).
94. 9575212: Sugimoto, H., Odani, S. & Yamashita, S. Cloning and expression of cDNA encoding rat liver 60-kDa lysophospholipase containing an asparaginase-like region and ankyrin repeat. *J Biol Chem* **273,** 12536–12542 (1998).
95. 9647737: Dousset, B. *et al.* Purification from human plasma of a hexapeptide that potentiates the sulfation and mitogenic activities of insulin-like growth factors. *Biochem Biophys Res Commun* **247,** 587–591 (1998).
96. 9654057: Ventura, F. V. *et al.* Carnitine palmitoyltransferase II specificity towards beta-oxidation intermediates--evidence for a reverse carnitine cycle in mitochondria. *Eur J Biochem* **253,** 614–618 (1998).
97. 9924800: Schenk, G., Duggleby, R. G. & Nixon, P. F. Properties and functions of the thiamin diphosphate dependent enzyme transketolase. *Int J Biochem Cell Biol* **30,** 1297–1318 (1998).
98. 9925947: Blackburn, A. C., Woollatt, E., Sutherland, G. R. & Board, P. G. Characterization and chromosome location of the gene GSTZ1 encoding the human Zeta class glutathione transferase and maleylacetoacetate isomerase. *Cytogenet Cell Genet* **83,** 109–114 (1998).
99. 10102904: Moolenaar, S. H. *et al.* Defect in dimethylglycine dehydrogenase, a new inborn error of metabolism: NMR spectroscopy study. *Clin Chem* **45,** 459–464 (1999).
100. 10329726: Hirohashi, T., Suzuki, H. & Sugiyama, Y. Characterization of the transport properties of cloned rat multidrug resistance-associated protein 3 (MRP3). *J Biol Chem* **274,** 15181–15185 (1999).
101. 10419495: Geisbrecht, B. V., Zhang, D., Schulz, H. & Gould, S. J. Characterization of PECI, a novel monofunctional Delta(3), Delta(2)-enoyl-CoA isomerase of mammalian peroxisomes. *J Biol Chem* **274,** 21797–21803 (1999).
102. 10441143: Brix, L. A., Barnett, A. C., Duggleby, R. G., Leggett, B. & McManus, M. E. Analysis of the substrate specificity of human sulfotransferases SULT1A1 and SULT1A3: site-directed mutagenesis and kinetic studies. *Biochemistry* **38,** 10474–10479 (1999).
103. 10471399: Rajan, D. P. *et al.* Human placental sodium-dependent vitamin C transporter (SVCT2): molecular cloning and transport function. *Biochem Biophys Res Commun* **262,** 762–768 (1999).
104. 10556521: Daruwala, R., Song, J., Koh, W. S., Rumsey, S. C. & Levine, M. Cloning and functional characterization of the human sodium-dependent vitamin C transporters hSVCT1 and hSVCT2. *FEBS Lett* **460,** 480–484 (1999).
105. 10588648: Pfeiffer, R. *et al.* Luminal heterodimeric amino acid transporter defective in cystinuria. *Mol Biol Cell* **10,** 4135–4147 (1999).
106. 10605936: Nebert, D. W. *et al.* Role of the aromatic hydrocarbon receptor and [Ah] gene battery in the oxidative stress response, cell cycle control, and apoptosis. *Biochem Pharmacol* **59,** 65–85 (2000).

107. 10620514: Gómez-Fabre, P. M. *et al.* Molecular cloning, sequencing and expression studies of the human breast cancer cell glutaminase. *Biochem J* **345 Pt 2,** 365–375 (2000).
108. 10744128: Gross, M. *et al.* Distribution and concordance of N-acetyltransferase genotype and phenotype in an American population. *Cancer Epidemiol Biomarkers Prev* **8,** 683–692 (1999).
109. 10748143: Kojima, Y. *et al.* Molecular cloning of globotriaosylceramide/CD77 synthase, a glycosyltransferase that initiates the synthesis of globo series glycosphingolipids. *J Biol Chem* **275,** 15152–15156 (2000).
110. 10777549: Jones, J. M., Morrell, J. C. & Gould, S. J. Identification and characterization of HAOX1, HAOX2, and HAOX3, three human peroxisomal 2-hydroxy acid oxidases. *J Biol Chem* **275,** 12590–12597 (2000).
111. 10794676: Wang, H. *et al.* Structure, function, and genomic organization of human Na(+)-dependent high-affinity dicarboxylate transporter. *Am J Physiol Cell Physiol* **278,** C1019-1030 (2000).
112. 10802064: Nava, V. E. *et al.* Functional characterization of human sphingosine kinase-1. *FEBS Lett* **473,** 81–84 (2000).
113. 10829015: Zimmer, S. *et al.* A novel human tocopherol-associated protein: cloning, in vitro expression, and characterization. *J Biol Chem* **275,** 25672–25680 (2000).
114. 10843803: Chen, Y. M. *et al.* Genomic structure, expression, and chromosomal localization of the human glycine N-methyltransferase gene. *Genomics* **66,** 43–47 (2000).
115. 10859351: Mandala, S. M. *et al.* Molecular cloning and characterization of a lipid phosphohydrolase that degrades sphingosine-1- phosphate and induces cell death. *Proc Natl Acad Sci U S A* **97,** 7859–7864 (2000).
116. 10860550: Kawashima, H. *et al.* Human fatty acid omega-hydroxylase, CYP4A11: determination of complete genomic sequence and characterization of purified recombinant protein. *Arch Biochem Biophys* **378,** 333–339 (2000).
117. 10860662: Marquardt, A., Stöhr, H., White, K. & Weber, B. H. cDNA cloning, genomic structure, and chromosomal localization of three members of the human fatty acid desaturase family. *Genomics* **66,** 175–183 (2000).
118. 10903140: Bröer, A., Wagner, C. A., Lang, F. & Bröer, S. The heterodimeric amino acid transporter 4F2hc/y+LAT2 mediates arginine efflux in exchange with glutamine. *Biochem J* **349 Pt 3,** 787–795 (2000).
119. 11067870: Schwarz, M. *et al.* The bile acid synthetic gene 3beta-hydroxy-Delta(5)-C(27)-steroid oxidoreductase is mutated in progressive intrahepatic cholestasis. *J Clin Invest* **106,** 1175–1184 (2000).
120. 11102558: Zschocke, J. *et al.* Progressive infantile neurodegeneration caused by 2-methyl-3-hydroxybutyryl-CoA dehydrogenase deficiency: a novel inborn error of branched-chain fatty acid and isoleucine metabolism. *Pediatr Res* **48,** 852–855 (2000).
121. 11248200: Akita, H. *et al.* Characterization of bile acid transport mediated by multidrug resistance associated protein 2 and bile salt export pump. *Biochim Biophys Acta* **1511,** 7–16 (2001).
122. 11279164: Lowenson JD, Kim E, Young SG, & Clarke, S. Limited accumulation of damaged proteins in l-isoaspartyl (D-aspartyl) O-methyltransferase-deficient mice. *J Biol Chem* **276**: 20695-702 (2001).
123. 11302742: Ichida, K., Matsumura, T., Sakuma, R., Hosoya, T. & Nishino, T. Mutation of human molybdenum cofactor sulfurase gene is responsible for classical xanthinuria type II. *Biochem Biophys Res Commun* **282,** 1194–1200 (2001).
124. 11306093: Aoki, S., Ishikura, S., Asada, Y., Usami, N. & Hara, A. Identity of dimeric dihydrodiol dehydrogenase as NADP(+)-dependent D-xylose dehydrogenase in pig liver. *Chem Biol Interact* **130–132,** 775–784 (2001).
125. 11342103: Charbonneau, A. & The, V. L. Genomic organization of a human 5beta-reductase and its pseudogene and substrate selectivity of the expressed enzyme. *Biochim Biophys Acta* **1517,** 228–235 (2001).
126. 11389701: de Graaf, M. *et al.* Cloning and characterization of human liver cytosolic beta-glycosidase. *Biochem J* **356,** 907–910 (2001).
127. 11401432: Yan, W. *et al.* Cloning and characterization of a human beta,beta-carotene-15,15'-dioxygenase that is highly expressed in the retinal pigment epithelium. *Genomics* **72,** 193–202 (2001).

128. 11737202: Traving, C., Bruse, P., Wächter, A. & Schauer, R. The sialate-pyruvate lyase from pig kidney. Elucidation of the primary structure and expression of recombinant enzyme activity. *Eur J Biochem* **268,** 6473–6486 (2001).
129. 11827462: Kim, D. K. *et al.* The human T-type amino acid transporter-1: characterization, gene organization, and chromosomal location. *Genomics* **79,** 95–103 (2002).
130. 11937514: Ternes, P., Franke, S., Zähringer, U., Sperling, P. & Heinz, E. Identification and characterization of a sphingolipid delta 4-desaturase family. *J Biol Chem* **277,** 25512–25518 (2002).
131. 11997390: Sontag, T. J. & Parker, R. S. Cytochrome P450 omega-hydroxylase pathway of tocopherol catabolism. Novel mechanism of regulation of vitamin E status. *J Biol Chem* **277,** 25290–25296 (2002).
132. 12023972: Farrar, C., & Clarke, S. Altered levels of S-adenosylmethionine and S-adenosylhomocysteine in the brains of L-isoaspartyl (D-Aspartyl) O-methyltransferase-deficient mice. *J Biol Chem* **277**: 27856-63 (2002).
133. 12072962: Elmore, B. O., Bollinger, J. A. & Dooley, D. M. Human kidney diamine oxidase: heterologous expression, purification, and characterization. *J Biol Inorg Chem* **7,** 565–579 (2002).
134. 12089149: Enomoto, A. *et al.* Molecular identification of a novel carnitine transporter specific to human testis. Insights into the mechanism of carnitine recognition. *J Biol Chem* **277,** 36262–36271 (2002).
135. 12093894: Pullinger, C. R. *et al.* Human cholesterol 7alpha-hydroxylase (CYP7A1) deficiency has a hypercholesterolemic phenotype. *J Clin Invest* **110,** 109–117 (2002).
136. 12105227: Venkataraman, K. *et al.* Upstream of growth and differentiation factor 1 (uog1), a mammalian homolog of the yeast longevity assurance gene 1 (LAG1), regulates N-stearoyl-sphinganine (C18-(dihydro)ceramide) synthesis in a fumonisin B1-independent manner in mammalian cells. *J Biol Chem* **277,** 35642–35649 (2002).
137. 12359132: Nguyen, T. V. *et al.* Identification of isobutyryl-CoA dehydrogenase and its deficiency in humans. *Mol Genet Metab* **77,** 68–79 (2002).
138. 12371743: Leonard, A. E. *et al.* Identification and expression of mammalian long-chain PUFA elongation enzymes. *Lipids* **37,** 733–740 (2002).
139. 12569161: Jin, W., Millar, J. S., Broedl, U., Glick, J. M. & Rader, D. J. Inhibition of endothelial lipase causes increased HDL cholesterol levels in vivo. *J Clin Invest* **111,** 357–362 (2003).
140. 12606581: Zhang, Y. *et al.* HDAC-6 interacts with and deacetylates tubulin and microtubules in vivo. *EMBO J* **22,** 1168–1179 (2003).
141. 12660232: Wu, T., Yankovskaya, V. & McIntire, W. S. Cloning, sequencing, and heterologous expression of the murine peroxisomal flavoprotein, N1-acetylated polyamine oxidase. *J Biol Chem* **278,** 20514–20525 (2003).
142. 12694300: Rutkowski, B. *et al.* N-methyl-2-pyridone-5-carboxamide: a novel uremic toxin? *Kidney Int Suppl* S19-21 (2003). doi:10.1046/j.1523-1755.63.s84.36.x
143. 12730697: Abifadel, M. *et al.* Mutations in PCSK9 cause autosomal dominant hypercholesterolemia. *Nat Genet* **34,** 154–156 (2003).
144. 12912983: Riebeling, C., Allegood, J. C., Wang, E., Merrill, A. H. & Futerman, A. H. Two mammalian longevity assurance gene (LAG1) family members, trh1 and trh4, regulate dihydroceramide synthesis using different fatty acyl-CoA donors. *J Biol Chem* **278,** 43452–43459 (2003).
145. 12917409: Wang, N. *et al.* ATP-binding cassette transporter A7 (ABCA7) binds apolipoprotein A-I and mediates cellular phospholipid but not cholesterol efflux. *J Biol Chem* **278,** 42906–42912 (2003).
146. 13018112: Hall, D. A. Histidine alpha-deaminase and the production of urocanic acid in the mammal. *Biochem J* **51,** 499–504 (1952).
147. 13032082: Knox, W. E. The relation of liver kynureninase to tryptophan metabolism in pyridoxine deficiency. *Biochem J* **53,** 379–385 (1953).
148. 13457352: Rendina, G. & Coon, M. J. Enzymatic hydrolysis of the coenzyme a thiol esters of beta-hydroxypropionic and beta-hydroxyisobutyric acids. *J Biol Chem* **225,** 523–534 (1957).
149. 13538975: Caravaca, J. & Grisolia, S. Enzymatic decarbamylation of carbamyl beta-alanine and carbamyl beta-aminoisobutyric acid. *J Biol Chem* **231,** 357–365 (1958).

150. 13575440: Axelrod, J. & Tomchick, R. Enzymatic O-methylation of epinephrine and other catechols. *J Biol Chem* **233,** 702–705 (1958).
151. 13672973: Tabor, H. & Wyngarden, L. The enzymatic formation of formiminotetrahydrofolic acid, 5,10-methenyltetrahydrofolic acid, and 10-formyltetrahydrofolic acid in the metabolism of formiminoglutamic acid. *J Biol Chem* **234,** 1830–1846 (1959).
152. 13716069: Hoskins, D. D. & Mackenzie, C. G. Solubilization and electron transfer flavoprtein requirement of mitochondrial sarcosine dehydrogenase and dimethylglycine dehydrogenase. *J Biol Chem* **236,** 177–183 (1961).
153. 13752080: Kaziro, Y., Ochoa, S., Warner, R. C. & Chen, J. Y. Metabolism of propionic acid in animal tissues. VIII. Crystalline propionyl carboxylase. *J Biol Chem* **236,** 1917–1923 (1961).
154. 13758723: Lane, M. D., Halenz, D. R., Kosow, D. P. & Hegre, C. S. Further studies on mitochondrial propionyl carboxylase. *J Biol Chem* **235,** 3082–3086 (1960).
155. 14081922: Kizer, D. E., Cox, B., Lovig, C. A. & Francodeestrugo, S. ADENYLIC ACID DEAMINASE OF RAT LIVER. *J Biol Chem* **238,** 3048–3052 (1963).
156. 14216443: Nagatsu, T., Levitt, M. & Udenfriend, S. TYROSINE HYDROXYLASE. The initial step in norepinephrine biosynthesis. *J Biol Chem* **239,** 2910–2917 (1964).
157. 14253475: Friedman, S. & Kaufman, S. 3,4-dihydroxyphenylethylamine beta-hydroxylase: a copper protein. *J Biol Chem* **240,** PC552-554 (1965).
158. 14504269: Graf, G. A. *et al.* ABCG5 and ABCG8 are obligate heterodimers for protein trafficking and biliary cholesterol excretion. *J Biol Chem* **278,** 48275–48282 (2003).
159. 14569421: Burk, O. & Wojnowski, L. Cytochrome P450 3A and their regulation. *Naunyn Schmiedebergs Arch Pharmacol* **369,** 105–124 (2004).
160. 14656720: Pushkin, A. *et al.* Structural characterization, tissue distribution, and functional expression of murine aminoacylase III. *Am J Physiol Cell Physiol* **286,** C848-856 (2004).
161. 14674884: Agrimi, G. *et al.* Identification of the human mitochondrial S-adenosylmethionine transporter: bacterial expression, reconstitution, functional characterization and tissue distribution. *Biochem J* **379,** 183–190 (2004).
162. 14794728: Mehler, A. H. & Knox, W. E. The conversion of tryptophan to kynurenine in liver. II. The enzymatic hydrolysis of formylkynurenine. *J Biol Chem* **187,** 431–438 (1950).
163. 15026176: Penning, T. M., Jin, Y., Steckelbroeck, S., Lanisnik Rizner, T. & Lewis, M. Structure-function of human 3 alpha-hydroxysteroid dehydrogenases: genes and proteins. *Mol Cell Endocrinol* **215,** 63–72 (2004).
164. 15044460: Bröer, A. *et al.* Molecular cloning of mouse amino acid transport system B0, a neutral amino acid transporter related to Hartnup disorder. *J Biol Chem* **279,** 24467–24476 (2004).
165. 15283699: Coleman, C. S., Stanley, B. A., Jones, A. D. & Pegg, A. E. Spermidine/spermine-N1-acetyltransferase-2 (SSAT2) acetylates thialysine and is not involved in polyamine metabolism. *Biochem J* **384,** 139–148 (2004).
166. 15361070: Molinaro, G. *et al.* Human recombinant membrane-bound aminopeptidase P: production of a soluble form and characterization using novel, internally quenched fluorescent substrates. *Biochem J* **385,** 389–397 (2005).
167. 15561973: Burckhardt, B. C., Lorenz, J., Kobbe, C. & Burckhardt, G. Substrate specificity of the human renal sodium dicarboxylate cotransporter, hNaDC-3, under voltage-clamp conditions. *Am J Physiol Renal Physiol* **288,** F792-799 (2005).
168. 15606768: Han, Q., Li, J. & Li, J. pH dependence, substrate specificity and inhibition of human kynurenine aminotransferase I. *Eur J Biochem* **271,** 4804–4814 (2004).
169. 15632147: Takanaga, H., Mackenzie, B., Suzuki, Y. & Hediger, M. A. Identification of mammalian proline transporter SIT1 (SLC6A20) with characteristics of classical system imino. *J Biol Chem* **280,** 8974–8984 (2005).
170. 15655246: Tsuboi, K. *et al.* Molecular characterization of N-acylethanolamine-hydrolyzing acid amidase, a novel member of the choloylglycine hydrolase family with structural and functional similarity to acid ceramidase. *J Biol Chem* **280,** 11082–11092 (2005).
171. 15702409: Matsuo, M., Tasaki, R., Kodama, H. & Hamasaki, Y. Screening for Menkes disease using the urine HVA/VMA ratio. *J Inherit Metab Dis* **28,** 89–93 (2005).
172. 15804236: Böhmer, C. *et al.* Characterization of mouse amino acid transporter B0AT1 (slc6a19). *Biochem J* **389,** 745–751 (2005).

173. 15809331: Ishikura, S., Usami, N., Araki, M. & Hara, A. Structural and functional characterization of rabbit and human L-gulonate 3-dehydrogenase. *J Biochem* **137,** 303–314 (2005).
174. 15907797: Wang, J. *et al.* Characterization of HSCD5, a novel human stearoyl-CoA desaturase unique to primates. *Biochem Biophys Res Commun* **332,** 735–742 (2005).
175. 15968458: Amici, A., Ciccioli, K., Naponelli, V., Raffaelli, N. & Magni, G. Evidence for essential catalytic determinants for human erythrocyte pyrimidine 5'-nucleotidase. *Cell Mol Life Sci* **62,** 1613–1620 (2005).
176. 15980104: Kimura, M. *et al.* Cyp2a6 is a principal enzyme involved in hydroxylation of 1,7-dimethylxanthine, a main caffeine metabolite, in humans. *Drug Metab Dispos* **33,** 1361–1366 (2005).
177. 16317684: Ballatori, N. *et al.* OSTalpha-OSTbeta: a major basolateral bile acid and steroid transporter in human intestinal, renal, and biliary epithelia. *Hepatology* **42,** 1270–1279 (2005).
178. 16399346: Court, M.H. Isoform-selective probe substrates for in vitro studies of human UDP-glucuronosyltransferases. *Methods Enzymol* **400**,104-16 (2005).
179. 16491085: Mills, P. B. *et al.* Mutations in antiquitin in individuals with pyridoxine-dependent seizures. *Nat Med* **12,** 307–309 (2006).
180. 16589713: Ichihara, A. & Greenberg, D. M. Pathway of serine formation from carbohydrate in rat liver. *Proc Natl Acad Sci U S A* **41,** 605–609 (1955).
181. 16807375: Shin, M., Kim, I., Inoue, Y., Kimura, S. & Gonzalez, F. J. Regulation of mouse hepatic alpha-amino-beta-carboxymuconate-epsilon-semialdehyde decarboxylase, a key enzyme in the tryptophan-nicotinamide adenine dinucleotide pathway, by hepatocyte nuclear factor 4alpha and peroxisome proliferator-activated receptor alpha. *Mol Pharmacol* **70,** 1281–1290 (2006).
182. 16842543: Ha, E. *et al.* Melatonin stimulates glucose transport via insulin receptor substrate-1/phosphatidylinositol 3-kinase pathway in C2C12 murine skeletal muscle cells. *J Pineal Res* **41,** 67–72 (2006).
183. 16874462: Hu, C. A. *et al.* Overexpression of proline oxidase induces proline-dependent and mitochondria-mediated apoptosis. *Mol Cell Biochem* **295,** 85–92 (2007).
184. 16940157: Hunt, M. C., Rautanen, A., Westin, M. A. K., Svensson, L. T. & Alexson, S. E. H. Analysis of the mouse and human acyl-CoA thioesterase (ACOT) gene clusters shows that convergent, functional evolution results in a reduced number of human peroxisomal ACOTs. *FASEB J* **20,** 1855–1864 (2006).
185. 16992869: Plimmer, R. H. On the presence of lactase in the intestines of animals and on the adaptation of the intestine to lactose. *J Physiol* **35,** 20–31 (1906).
186. 17015445: Wei, B. Q., Mikkelsen, T. S., McKinney, M. K., Lander, E. S. & Cravatt, B. F. A second fatty acid amide hydrolase with variable distribution among placental mammals. *J Biol Chem* **281,** 36569–36578 (2006).
187. 17023427: Hornemann, T., Richard, S., Rütti, M. F., Wei, Y. & von Eckardstein, A. Cloning and initial characterization of a new subunit for mammalian serine-palmitoyltransferase. *J Biol Chem* **281,** 37275–37281 (2006).
188. 17393504: Shin, H. J. *et al.* Novel liver-specific organic anion transporter OAT7 that operates the exchange of sulfate conjugates for short chain fatty acid butyrate. *Hepatology* **45,** 1046–1055 (2007).
189. 17412732: Gempel, K. *et al.* The myopathic form of coenzyme Q10 deficiency is caused by mutations in the electron-transferring-flavoprotein dehydrogenase (ETFDH) gene. *Brain* **130,** 2037–2044 (2007).
190. 17707131: Yamashita, A. *et al.* Topology of acyltransferase motifs and substrate specificity and accessibility in 1-acyl-sn-glycero-3-phosphate acyltransferase 1. *Biochim Biophys Acta* **1771,** 1202–1215 (2007).
191. 17896864: Kaper, T. *et al.* Nanosensor detection of an immunoregulatory tryptophan influx/kynurenine efflux cycle. *PLoS Biol* **5,** e257 (2007).
192. 18088226: Dungan, K. M. 1,5-anhydroglucitol (GlycoMark) as a marker of short-term glycemic control and glycemic excursions. *Expert Rev Mol Diagn* **8,** 9–19 (2008).
193. 18094042: Lee, H.-C. *et al.* Caenorhabditis elegans mboa-7, a member of the MBOAT family, is required for selective incorporation of polyunsaturated fatty acids into phosphatidylinositol. *Mol Biol Cell* **19,** 1174–1184 (2008).

194. 18222180: Lindner, H. A., Täfler-Naumann, M. & Röhm, K.-H. N-acetylamino acid utilization by kidney aminoacylase-1. *Biochimie* **90,** 773–780 (2008).
195. 18230605: Szegedi, S. S., Castro, C. C., Koutmos, M. & Garrow, T. A. Betaine-homocysteine S-methyltransferase-2 is an S-methylmethionine-homocysteine methyltransferase. *J Biol Chem* **283,** 8939–8945 (2008).
196. 18327257: Vitart, V. *et al.* SLC2A9 is a newly identified urate transporter influencing serum urate concentration, urate excretion and gout. *Nat Genet* **40,** 437–442 (2008).
197. 18632736: Yonezawa, A., Masuda, S., Katsura, T. & Inui, K. Identification and functional characterization of a novel human and rat riboflavin transporter, RFT1. *Am J Physiol Cell Physiol* **295,** C632-641 (2008).
198. 18775983: Novick, R. M. & Elfarra, A. A. Purification and characterization of flavin-containing monooxygenase isoform 3 from rat kidney microsomes. *Drug Metab Dispos* **36,** 2468–2474 (2008).
199. 18902391: Kazenko, A. & Laskowski, M. On the specificity of chicken pancreas conjugase (gamma-glutamic acid carboxypeptidase). *J Biol Chem* **173,** 217–221 (1948).
200. 18941301: Kikuchi, G., Motokawa, Y., Yoshida, T. & Hiraga, K. Glycine cleavage system: reaction mechanism, physiological significance, and hyperglycinemia. *Proc Jpn Acad Ser B Phys Biol Sci* **84,** 246–263 (2008).
201. 18981171: Mackenzie, P. I. *et al.* Identification of UDP glycosyltransferase 3A1 as a UDP N-acetylglucosaminyltransferase. *J Biol Chem* **283,** 36205–36210 (2008).
202. 19339287: Eminoglu, F. T. *et al.* 3-Methylcrotonyl-CoA carboxylase deficiency: phenotypic variability in a family. *J Child Neurol* **24,** 478–481 (2009).
203. 19411760: Lu, G. *et al.* Protein phosphatase 2Cm is a critical regulator of branched-chain amino acid catabolism in mice and cultured cells. *J Clin Invest* **119,** 1678–1687 (2009).
204. 19453107: Zhao, H., Martin, B. M., Bisoffi, M. & Dunaway-Mariano, D. The Akt C-terminal modulator protein is an acyl-CoA thioesterase of the Hotdog-Fold family. *Biochemistry* **48,** 5507–5509 (2009).
205. 19506252: Woodward, O. M. *et al.* Identification of a urate transporter, ABCG2, with a common functional polymorphism causing gout. *Proc Natl Acad Sci U S A* **106,** 10338–10342 (2009).
206. 19595734: Krasnikov, B. F. *et al.* Identification of the putative tumor suppressor Nit2 as omega-amidase, an enzyme metabolically linked to glutamine and asparagine transamination. *Biochimie* **91,** 1072–1080 (2009).
207. 19679820: Fujita, M. *et al.* Hepatic uptake of gamma-butyrobetaine, a precursor of carnitine biosynthesis, in rats. *Am J Physiol Gastrointest Liver Physiol* **297,** G681-686 (2009).
208. 20285042: Kalckar, H. M. The enzymatic synthesis of purine ribosides. *J Biol Chem* **167,** 477–486 (1947).
209. 20335586: Magen, D. *et al.* A loss-of-function mutation in NaPi-IIa and renal Fanconi's syndrome. *N Engl J Med* **362,** 1102–1109 (2010).
210. 21515882: Wu, L.-C. *et al.* Purification, identification, and cloning of lysoplasmalogenase, the enzyme that catalyzes hydrolysis of the vinyl ether bond of lysoplasmalogen. *J Biol Chem* **286,** 24916–24930 (2011).
211. 21733844: Öberg, F. *et al.* Glycosylation increases the thermostability of human aquaporin 10 protein. *J Biol Chem* **286,** 31915–31923 (2011).
212. 21846720: Witkowski, A., Thweatt, J. & Smith, S. Mammalian ACSF3 protein is a malonyl-CoA synthetase that supplies the chain extender units for mitochondrial fatty acid synthesis. *J Biol Chem* **286,** 33729–33736 (2011).
213. 21886157: Suhre, K. *et al.* Human metabolic individuality in biomedical and pharmaceutical research. *Nature* **477,** 54–60 (2011).
214. 21908619: Collard, F., Vertommen, D., Constantinescu, S., Buts, L. & Van Schaftingen, E. Molecular identification of β-citrylglutamate hydrolase as glutamate carboxypeptidase 3. *J Biol Chem* **286,** 38220–38230 (2011).
215. 21926997: Long, J. Z. *et al.* Metabolomics annotates ABHD3 as a physiologic regulator of medium-chain phospholipids. *Nat Chem Biol* **7,** 763–765 (2011).
216. 22016388: Linster, C. L. *et al.* Ethylmalonyl-CoA decarboxylase, a new enzyme involved in metabolite proofreading. *J Biol Chem* **286,** 42992–43003 (2011).
217. 22063269: Mattijssen, F. & Kersten, S. Regulation of triglyceride metabolism by Angiopoietin-like proteins. *Biochim Biophys Acta* **1821,** 782–789 (2012).

218. 22241472: Veiga-da-Cunha, M., Hadi, F., Balligand, T., Stroobant, V. & Van Schaftingen, E. Molecular identification of hydroxylysine kinase and of ammoniophospholyases acting on 5-phosphohydroxy-L-lysine and phosphoethanolamine. *J Biol Chem* **287,** 7246–7255 (2012).
219. 22427816: Ehrlund, A. *et al.* Knockdown of SF-1 and RNF31 affects components of steroidogenesis, TGFβ, and Wnt/β-catenin signaling in adrenocortical carcinoma cells. *PLoS One* **7,** e32080 (2012).
220. 22586271: Zhuravleva, E. *et al.* Acyl coenzyme A thioesterase Them5/Acot15 is involved in cardiolipin remodeling and fatty liver development. *Mol Cell Biol* **32,** 2685–2697 (2012).
221. 22952014: Tamai, I. Pharmacological and pathophysiological roles of carnitine/organic cation transporters (OCTNs: SLC22A4, SLC22A5 and Slc22a21). *Biopharm Drug Dispos* **34**:29-44 (2013).
222. 23010440: Davids, M., Ndika, J. D. T., Salomons, G. S., Blom, H. J. & Teerlink, T. Promiscuous activity of arginine:glycine amidinotransferase is responsible for the synthesis of the novel cardiovascular risk factor homoarginine. *FEBS Lett* **586,** 3653–3657 (2012).
223. 23024808: De Ingeniis, J. *et al.* Functional specialization in proline biosynthesis of melanoma. *PLoS One* **7,** e45190 (2012).
224. 23038267: Liu, P. *et al.* Role of glutamate decarboxylase-like protein 1 (GADL1) in taurine biosynthesis. *J Biol Chem* **287,** 40898–40906 (2012).
225. 23141293: Danhauser, K. *et al.* DHTKD1 mutations cause 2-aminoadipic and 2-oxoadipic aciduria. *Am J Hum Genet* **91,** 1082–1087 (2012).
226. 23266187: Palmieri, F. The mitochondrial transporter family SLC25: identification, properties and physiopathology. *Mol Aspects Med* **34,** 465–484 (2013).
227. 23506882: Bürzle, M. *et al.* The sodium-dependent ascorbic acid transporter family SLC23. *Mol Aspects Med* **34,** 436–454 (2013).
228. 23798576: Karamitri, A., Renault, N., Clement, N., Guillaume, J.-L. & Jockers, R. Minireview: Toward the establishment of a link between melatonin and glucose homeostasis: association of melatonin MT2 receptor variants with type 2 diabetes. *Mol Endocrinol* **27,** 1217–1233 (2013).
229. 23863933: Simanshu, D. K. *et al.* Non-vesicular trafficking by a ceramide-1-phosphate transfer protein regulates eicosanoids. *Nature* **500,** 463–467 (2013).
230. 24121108: Yamada, S. & Arikawa, S. An ectotherm homologue of human predicted gene NAT16 encodes histidine N-acetyltransferase responsible for Nα-acetylhistidine synthesis. *Biochim Biophys Acta* **1840,** 434–442 (2014).
231. 24573086: Ghezzi, C. *et al.* Fingerprints of hSGLT5 sugar and cation selectivity. *Am J Physiol Cell Physiol* **306,** C864-870 (2014).
232. 24586186: Rueedi, R. *et al.* Genome-wide association study of metabolic traits reveals novel gene-metabolite-disease links. *PLoS Genet* **10,** e1004132 (2014).
233. 24652292: Porcelli, V., Fiermonte, G., Longo, A. & Palmieri, F. The human gene SLC25A29, of solute carrier family 25, encodes a mitochondrial transporter of basic amino acids. *J Biol Chem* **289,** 13374–13384 (2014).
234. 24697329: Wichelecki, D. J. *et al.* Enzymatic and structural characterization of rTSγ provides insights into the function of rTSβ. *Biochemistry* **53,** 2732–2738 (2014).
235. 24768817: Nomme, J., Su, Y. & Lavie, A. Elucidation of the specific function of the conserved threonine triad responsible for human L-asparaginase autocleavage and substrate hydrolysis. *J Mol Biol* **426,** 2471–2485 (2014).
236. 24847004: Houten, S. M. *et al.* Mitochondrial NADP(H) deficiency due to a mutation in NADK2 causes dienoyl-CoA reductase deficiency with hyperlysinemia. *Hum Mol Genet* **23,** 5009–5016 (2014).
237. 24891507: Veiga-da-Cunha, M., Chevalier, N., Stroobant, V., Vertommen, D. & Van Schaftingen, E. Metabolite proofreading in carnosine and homocarnosine synthesis: molecular identification of PM20D2 as β-alanyl-lysine dipeptidase. *J Biol Chem* **289,** 19726–19736 (2014).
238. 24927523: Mahdessian, H. *et al.* TM6SF2 is a regulator of liver fat metabolism influencing triglyceride secretion and hepatic lipid droplet content. *Proc Natl Acad Sci U S A* **111,** 8913–8918 (2014).
239. 25231977: Pinto, J. T. *et al.* Kynurenine aminotransferase III and glutamine transaminase L are identical enzymes that have cysteine S-conjugate β-lyase activity and can transaminate L-selenomethionine. *J Biol Chem* **289,** 30950–30961 (2014).

240. 25760036: Ellis, J. M., Bowman, C. E. & Wolfgang, M. J. Metabolic and tissue-specific regulation of acyl-CoA metabolism. *PLoS One* **10**, e0116587 (2015).
241. 25947375: Naito, T. *et al.* Phospholipid Flippase ATP10A Translocates Phosphatidylcholine and Is Involved in Plasma Membrane Dynamics. *J Biol Chem* **290,** 15004–15017 (2015).
242. 26014429: Heemskerk, M. M., van Harmelen, V. J. A., van Dijk, K. W. & van Klinken, J. B. Reanalysis of mGWAS results and in vitro validation show that lactate dehydrogenase interacts with branched-chain amino acid metabolism. *Eur J Hum Genet* **24,** 142–145 (2016).
243. 26234207: Mattjus, P. Specificity of the mammalian glycolipid transfer proteins. *Chem Phys Lipids* **194,** 72–78 (2016).
244. 26517927: Bauer, R. C., Yenilmez, B. O. & Rader, D. J. Tribbles-1: a novel regulator of hepatic lipid metabolism in humans. *Biochem Soc Trans* **43,** 1079–1084 (2015).
245. 27180734: Deol, R. & Josephy, P. D. Acetylation of aromatic cysteine conjugates by recombinant human N-acetyltransferase 8. *Xenobiotica* **47,** 202–207 (2017).
246. 27288731: Halwachs, S., Schäfer, I., Kneuer, C., Seibel, P. & Honscha, W. Assessment of ABCG2-mediated transport of pesticides across the rabbit placenta barrier using a novel MDCKII in vitro model. *Toxicol Appl Pharmacol* **305,** 66–74 (2016).
247. 28257906: Holmes, R. S. Comparative and evolutionary studies of mammalian arylsulfatase and sterylsulfatase genes and proteins encoded on the X-chromosome. *Comput Biol Chem* **68,** 71–77 (2017).
248. 28465440: Toomey, M. B. *et al.* High-density lipoprotein receptor SCARB1 is required for carotenoid coloration in birds. *Proc Natl Acad Sci U S A* **114,** 5219–5224 (2017).
249. 28516954: Hai, Y., Shinsky, S. A., Porter, N. J. & Christianson, D. W. Histone deacetylase 10 structure and molecular function as a polyamine deacetylase. *Nat Commun* **8,** 15368 (2017).
250. 28642719: Eriksson, O., Lalowski, M. & Lindholm, D. Commentary: LACTB is a tumour suppressor that modulates lipid metabolism and cell state. *Front Physiol* **8,** 396 (2017).
251. 28942964: Kim, H. I. *et al.* Fine Mapping and Functional Analysis Reveal a Role of SLC22A1 in Acylcarnitine Transport. *Am J Hum Genet* **101,** 489–502 (2017).
252. 29578721: Zhang, Q., Ford, L. A., Evans, A. M. & Toal, D. R. Identification of an Endogenous Organosulfur Metabolite by Interpretation of Mass Spectrometric Data. *Org Lett* **20,** 2100–2103 (2018).
253. 30913229: Pirim, D. *et al.* Apolipoprotein E-C1-C4-C2 gene cluster region and inter-individual variation in plasma lipoprotein levels: a comprehensive genetic association study in two ethnic groups. *PLoS One* **14,** e0214060 (2019).
254. 31764986: Laufs, U., Parhofer, K. G., Ginsberg, H. N. & Hegele, R. A. Clinical review on triglycerides. *Eur Heart J* **41,** 99-109c (2020).
255. 33415501: Di, L., Balesano, A., Jordan, S. & Shi, S. M. The Role of Alcohol Dehydrogenase in Drug Metabolism: Beyond Ethanol Oxidation. *AAPS J* **23,** 20 (2021).
